# The Interaction Effects of Sex, Age, APOE and Common Health Risk Factors on Human Brain Functions

**DOI:** 10.1101/2024.08.05.24311482

**Authors:** Tengfei Li, Jie Chen, Bingxin Zhao, Gwenn A. Garden, Kelly S. Giovanello, Guorong Wu, Hongtu Zhu

**Affiliations:** Biomedical Research Imaging Center, School of Medicine, University of North Carolina at Chapel Hill, Chapel Hill, NC, USA; Department of Radiology, School of Medicine, University of North Carolina at Chapel Hill, Chapel Hill, NC, USA; Department of Biostatistics, University of North Carolina at Chapel Hill, Chapel Hill, NC, USA; Department of Statistics and Data Science, the Wharton School, University of Pennsylvania, Philadelphia, PA, USA; Department of Neurology, School of Medicine, University of North Carolina at Chapel Hill, Chapel Hill, NC, USA; Department of Psychiatry, School of Medicine, University of North Carolina at Chapel Hill, Chapel Hill, NC, USA; Departments of Statistics and Computer Science, University of North Carolina at Chapel Hill, Chapel Hill, NC, USA; UNC Neuroscience Center, University of North Carolina at Chapel Hill, Chapel Hill, NC, USA; Carolina Insititute for Developmental Disabilities, Chapel Hill, NC, USA; Departments of Genetics, University of North Carolina at Chapel Hill, Chapel Hill, NC, USA

## Abstract

**Importance:** Nonlinear changes in brain function during aging are shaped by a complex interplay of factors, including sex, age, genetics, and modifiable health risk factors. However, the combined effects and underlying mechanisms of these factors on brain functional connectivity remain poorly understood.

**Objective:** To comprehensively investigate the combined associations of sex, age, *APOE* genotypes, and ten common modifiable health risk factors with brain functional connectivities during aging.

**Design, Setting, and Participants:** This analysis used data from 36,630 UK Biobank participants, aged 44–81, who were assessed for sex, age, *APOE* genotypes, 10 health risk factors, and brain functional connectivities through resting-state functional magnetic resonance imaging.

**Main Outcomes and Measures:** Brain functional connectivities were evaluated through within- and between-network functional connectivities and connectivity strength. Associations between risk factors and brain functional connectivities, including their interaction effects, were analyzed.

**Results:** Hypertension, BMI, and education were the top three influential factors. Sex-specific effects were also observed in interactions involving APOE4 gene, smoking, alcohol consumption, diabetes, BMI, and education. Notably, a negative sex-excessive alcohol interaction showed a stronger negative effect on functional connectivities in males, particularly between the dorsal attention network and the language network, while moderate alcohol consumption appeared to have protective effects. A significant negative interaction between sex and *APOE4* revealed a greater reduction in functional connectivity between the cingulo-opercular network and the posterior multimodal network in male *APOE4* carriers. Additional findings included a negative age-BMI interaction between the visual and dorsal attention networks, and a positive age-hypertension interaction between the frontoparietal and default mode networks.

**Conclusions and Relevance:** The findings highlight significant sex disparities in the associations between age, the *APOE*-ε4 gene, modifiable health risk factors, and brain functional connectivity, emphasizing the necessity of jointly considering these factors to gain a deeper understanding of the complex processes underlying brain aging.

## INTRODUCTION

Brain structural and functional aging exhibits substantial variability, influenced by factors such as sex, age, genetics, and modifiable health risk factors (MHRFs) including socioeconomic status (SES), lifestyle, and cardiovascular risk factors (CVRFs) ^1,2^. Our previous research examined the impact of MHRFs and their interactions on brain structure during aging ^3^. Building on this, it is essential to explore how these factors collectively affect brain functionality.

Age, sex, and Apolipoprotein E (*APOE*) genotypes are well-studied non-modifiable risk factors affecting brain function and cognition during aging^4^. Cognitive domains follow different trajectories, with vocabulary remaining stable while memory, reasoning, and processing speed decline ^5–7^. These cognitive changes can be partially explained by aging-related alterations in brain networks, including reduced connectivity within and between networks, decreased default mode network (DMN) connectivity, and network reorganization^8–16^. *APOE*-ε4 (*APOE4*), a key genetic risk factor for Alzheimer’s disease (AD), is associated with altered connectivity in memory and cognitive networks, accelerated age-related connectivity loss, and sometimes increased hyperconnectivity ^17–21^. The effects of *APOE*4 differ by sex ^22^ and age ^23^, with a stronger impact on attention in women and memory and executive functions in men ^24^. However, most studies emphasize the vulnerability of female *APOE*4 carriers, with limited data on functional atrophy in males ^3,22,25^. The joint influence of age, sex, *APOE*, and other MHRFs on brain function also remains unclear.

CVRFs, lifestyle factors, and SES are extensively studied in relation to brain and cardiovascular health. Key CVRFs identified by the Framingham Heart Study—such as hypertension, smoking, cholesterol, diabetes, obesity, and family history of heart disease—can impair brain health and contribute to aging and neurodegenerative diseases ^26–30^. The American Heart Association’s Life’s Essential 8 highlights crucial lifestyle measures like diet, physical activity, and sleep for maintaining cardiovascular and brain health ^31^. Recent studies show a strong link between cardiovascular and brain aging, with cardiovascular dysfunction from risk factors potentially impairing brain health ^32–34^. The 2020 Lancet Commission report suggests that modifying 12 major dementia risk factors could prevent or delay up to 40% of dementia cases ^35^.

This study aims to perform a comprehensive association analysis between brain functional connectivity (FC) and various risk factors, including *APOE* genotype, age, sex, and ten MHRFs (**Figure 1**). These MHRFs encompass six adverse factors—hypertension ^36–40^, diabetes ^41–43^, smoking ^44–46^, obesity ^47–49^, excessive alcohol consumption ^50,51^, and social deprivation ^52^—and four beneficial ones—education ^53–56^, physical activity ^57^, healthy diet ^58^ and sleep ^59^ that may affect brain FCs. Using resting-state functional magnetic resonance imaging (rsfMRI) data from 36,630 UK Biobank (UKB) subjects, this study seeks to uncover new insights into the dynamics of aging and preclinical dementia risks, particularly focusing on sex disparities.

**Figure 1.**
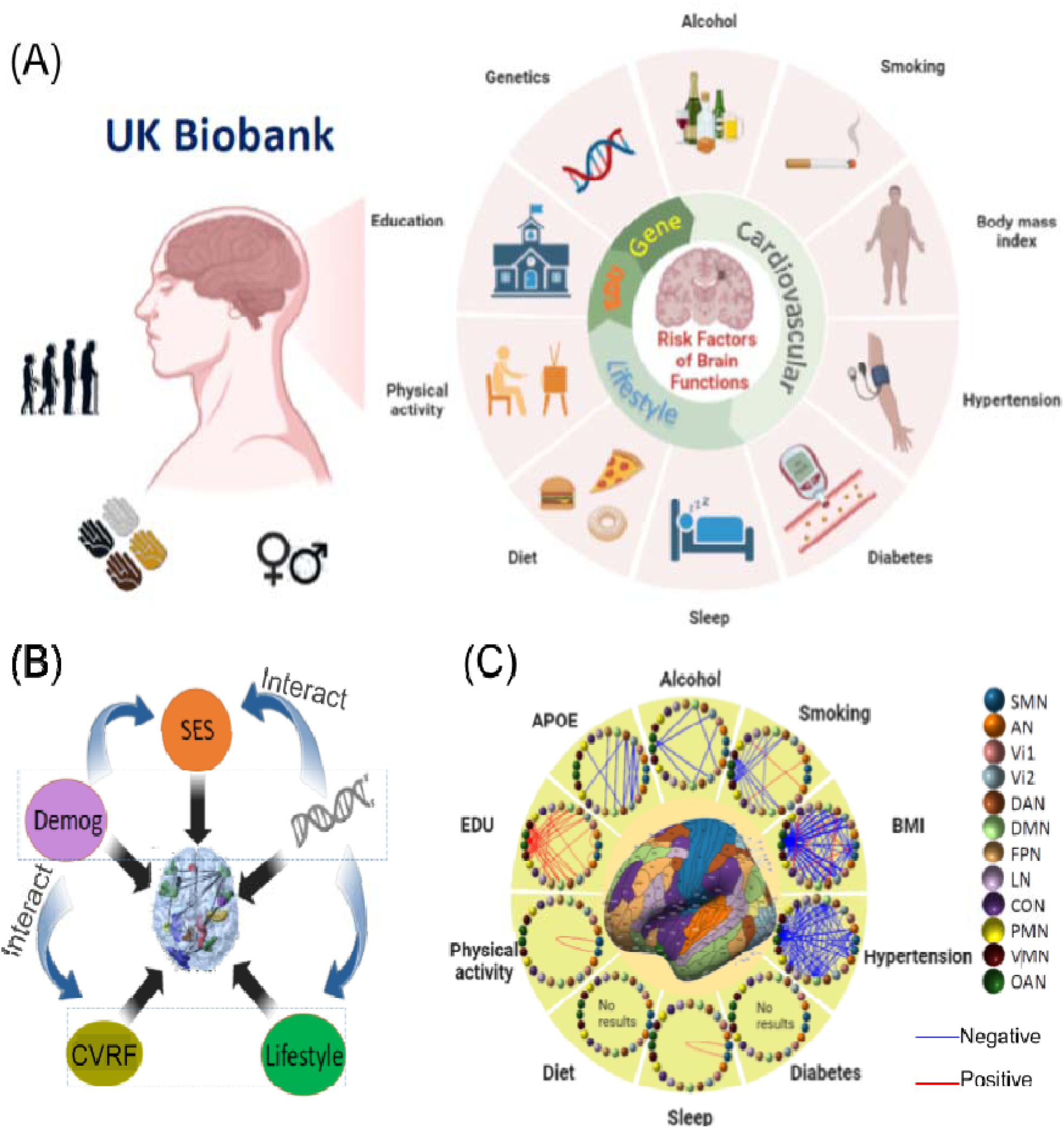
The study design. **(A)** Multimodal data in the UK Biobank. **(B)** Schematic diagram illustrating the analyzed associations between the *APOE* gene, demographics, modifiable health risk factors, and brain functional traits. **(C)** Identified brain network pairs based on the Ji-12 atlas, showing network edge strength (NES) measures associated with *APOE*, alcohol, smoking, BMI, physical activity, diet, sleep, education, diabates and hypertension (main effects only; no interactions considered). SMN, somatomotor network; AN, auditory network; Vi1, Vi2, visual networks 1 and 2; DAN, dorsal attention network; DMN, default mode network; FPN, frontoparietal network; LN, language network; CON, cingulo-opercular network; PMN/VMN, posterior/ventral multimodal network; OAN, orbito-affective network; SES, socioeconomic status (such as education); Demog: demographics (age and sex); EDU: education; *APOE*: apolipoprotein E; CVRF: cardiovascular risk factors; BMI, Body Mass Index.

## METHODS

### Study Population

The UKB dataset comprises around 500,000 participants, including over 40,000 who underwent MRI scans. We analyzed data from 39,354 subjects with both T1-weighted and rsfMRI scans, including 36,339 with British ancestry and 3,015 with non-British ancestry. The primary analyses focused on 36,630 unrelated individuals with complete brain imaging, genetic, and MHRF data. Key findings were based on 33,824 white British subjects, with subsequent validation in 2,806 non-British subjects.

### Imaging Data Processing

Imaging data acquisition and preprocessing followed protocols outlined in the UKB Brain Imaging Documentation (https://biobank.ctsu.ox.ac.uk/crystal/crystal/docs/brain_mri.pdf). RsfMRI data underwent motion correction, intensity normalization, and ICA-based artifact removal. High-quality T1- weighted images were preprocessed and co-registered rsfMRI data to MNI standard space. The HCP-MMP ^60^ atlas was employed to generate 64,620 functional connectivity (FC) traits per subject, which were then classified into twelve resting-state networks (Ji-12 network) ^61^, namely, the somatomotor (SMN), auditory (AN), visual1 (Vi1), visual2 (Vi2), dorsal attention (DAN), DMN, frontoparietal (FPN), language (LAN), cingulo-opercular (CON), posterior multimodal (PMN), ventral multimodal (VMN), and orbito-Affective (OAN) networks. For each rsfMRI scan, the mean time series from each of 360 ROIs were extracted and the correlation between each pair of regional time series was transformed from Pearson correlations to z-statistics using Fisher transformation. For each pair of the 12 networks we calculated the between- and within-network functional connectivity (NFC; or mean FC) and edge strength (NES; or mean absolute value of FC) measures. We also generated 100 × 100 FC matrices based on Schaefer 100 parcellation atlas and network-level traits for the Yeo-7 and Yeo-17 atlases ^62^. Detailed imaging protocol and processing steps were provided in **<u>Supplement 1</u>**.

### *APOE* Genotyping and covariates

*APOE* haplotypes were determined using SNPs rs7412 and rs429358, defining three alleles—*APOE*-ε2 (*APOE2*), *APOE*-ε3, and *APOE*4. Details on MHRFs and other covariates are provided in **<u>Supplement 1</u>**.

### Statistical Analysis

Statistical analyses were conducted using R version 4.1.0. *APOE*2 and *APOE4* were modeled using additive models. We analyzed between- and within-network NFC and NES measures, considering *APOE4* and *APOE2* counts, sex, age, and MHRFs as covariates of interest. To investigate interactions between age or *APOE* genotype and MHRFs on FC measures, we included two-way interactions (age or *APOE* with MHRFs) as well as age-*APOE*-MHRF interactions. To assess sex differences, two-way interactions between sex and age, *APOE* genotype, and MHRFs were incorporated, along with sex-related three-way interactions (sex-age-MHRF, sex-*APOE*-MHRF, and sex-age-*APOE*). Each main and interaction effect was tested separately, controlling for age, sex, all the other risk factors, head motion, brain position, volumetric scaling, study site, phase, and living with a partner. Age-squared and sex-age-squared interactions were also controlled, except when sex, age, or sex-age interaction were terms of interest. Detailed models are available in eTable 2, **<u>Supplement 2</u>**. Type II ANOVA F-tests were used to assess the main and interaction effects ^63^. Our preliminary findings, based on white British subjects, were evaluated using Bonferroni-corrected significance levels of *0.05/m* (details on the number of tests *m*, are provided in eTable 3, **<u>Supplement 2</u>**). Results were validated in non-British UKB populations by consistency of association directions. Post hoc analyses of specific ROIs within identified networks were conducted for each risk factor, with the same confounding covariates and Bonferroni correction methods (*m* being the number of ROI pairs in identified networks for each risk factor).

## RESULTS

### Participant Characteristics

Demographic information for the 36,630 UKB subjects is summarized in **Table 1**. Correlation plots for *APOE* gene and MHRFs (eFigure 1, **<u>Supplement 3</u>**) show weak correlations (*r < 0.3* except for moderate and excessive alcohol consumption). Population-mean NFC and NES matrices for the Ji-12, Yeo-7, and Yeo-17 atlases (eFigures 43-45, **<u>Supplement 3</u>**) reveals anti-correlations between both the DMN and VMN with the AN, CON, DAN, PMN, SMN, Vi1, and Vi2 networks, suggesting competitive or complementary functions critical for cognitive processing. Demographic differences between populations are detailed in eTables 4 and 5 (**<u>Supplement 2</u>**). With a Bonferroni-corrected significance level of 0.0038, the non-British cohort was, on average, two years younger, with fewer ever-smokers (-3.3%) and fewer partnered individuals (-6.9%), but higher proportions of advanced education (14.1%), higher SoDep index (17.1%), excessive alcohol consumption (10.8%), and diabetes (1.5%).

**Table 1.** Demographic information, *APOE* gene, and modifiable health-related risk factors for 36,630 UK Biobank subjects. No., sample size; SD, standard deviation; SoDep, social deprivation.

| Characteristic | Female (N=19395) | Male (N=17235) | Total (N=36630) |
| --- | --- | --- | --- |
| Age at imaging: mean (SD) [range] | 63.0 (7.39)<br>[45.0, 81.0] | 64.3 (7.65)<br>[44.0, 81.0] | 63.6 (7.54)<br>[44.0, 81.0] |
| Ethnicity---British: No. (%) | 17861 (92.1%) | 15963 (92.6%) | 33824 (92.3%) |
| Ethnicity---non-British: No. (%) | 1534 (7.9%) | 1272 (7.4%) | 2806 (7.7%) |
| *APOE4* = 0: No. (%) | 13868 (71.5%) | 12576 (73.0%) | 26444 (72.2%) |
| *APOE4* = 1: No. (%) | 5085 (26.2%) | 4295 (24.9%) | 9380 (25.6%) |
| *APOE4* = 2: No. (%) | 442 (2.3%) | 364 (2.1%) | 806 (2.2%) |
| *APOE2* = 0: No. (%) | 16378 (84.4%) | 14619 (84.8%) | 30997 (84.6%) |
| *APOE2* = 1: No. (%) | 2904 (15.0%) | 2518 (14.6%) | 5422 (14.8%) |
| *APOE2* = 2: No. (%) | 113 (0.6%) | 98 (0.6%) | 211 (0.6%) |
| Marriage---living with a partner: No. (%) | 13619 (70.2%) | 13913 (80.7%) | 27532 (75.2%) |
| Marriage---not with a partner: No. (%) | 1235 (6.4%) | 520 (3.0%) | 1755 (4.8%) |
| Education---college degree/above: No. (%) | 8626 (44.5%) | 8293 (48.1%) | 16919 (46.2%) |
| Education---no college/below: No. (%) | 9433 (48.6%) | 7737 (44.9%) | 17170 (46.9%) |
| SoDep Index---above the median: No. (%) | 8275 (42.7%) | 7092 (41.1%) | 15367 (42.0%) |
| SoDep Index---below the median: No. (%) | 11099 (57.2%) | 10130 (58.8%) | 21229 (58.0%) |
| Smoking status---ever smoked: No. (%) | 12459 (64.2%) | 9867 (57.2%) | 22326 (61.0%) |
| Smoking status---never smoked: No. (%) | 6936 (35.8%) | 7368 (42.8%) | 14304 (39.1%) |
| Regular physical activity---yes: No. (%) | 14199 (73.2%) | 13267 (77.0%) | 27466 (75.0%) |
| Regular physical activity---no: No. (%) | 5040 (26.0%) | 3893 (22.6%) | 8933 (24.4%) |
| Health diet---yes: No. (%) | 10517 (54.2%) | 7371 (42.8%) | 17888 (48.8%) |
| Health diet---no: No. (%) | 8870 (45.7%) | 9854 (57.2%) | 18724 (51.1%) |
| Alcohol consumption---excessive: No. (%) | 2742 (14.1%) | 1284 (7.5%) | 4026 (11.0%) |
| Alcohol consumption---moderate: No. (%) | 11115 (57.3%) | 11096 (64.4%) | 22211 (60.6%) |
| Alcohol consumption---not current: No. (%) | 5531 (28.5%) | 4848 (28.1%) | 10379 (28.3%) |
| Sleeping---between 6-8 hours: No. (%) | 8193 (42.2%) | 7759 (45.0%) | 15952 (43.5%) |
| Sleeping---outside 6-8 hours: No. (%) | 11145 (57.5%) | 9455 (54.9%) | 20600 (56.2%) |
| Hypertension---yes: No. (%) | 3060 (15.8%) | 4259 (24.7%) | 7319 (20.0%) |
| Hypertension---no: No. (%) | 16335 (84.2%) | 12976 (75.3%) | 29311 (80.0%) |
| Diabetes---yes: No. (%) | 573 (3.0%) | 975 (5.7%) | 1548 (4.2%) |
| Diabetes---no: No. (%) | 18822 (97.0%) | 16260 (94.3%) | 35082 (95.8%) |
| BMI: mean (SD) [range] | 26.0 (4.53)<br>[14.7,56.6] | 27.1 (3.73)<br>[16.7,56.0] | 26.5 (4.20)<br>[14.7,56.6] |

### Overall Findings

Findings for NFC and NES measures across the Ji-12, Yeo-7, and Yeo-17 atlases are summarized in eTable 6, **<u>Supplement 2</u>**, and depicted in eFigures 37-42, **<u>Supplement 3</u>**. Main effects are presented in eFigures 2-7, **<u>Supplement 3</u>**, while two-way and three-way interaction effects are shown in eFigures 8-19, **<u>Supplement 3</u>**. In the white British population, we identified 113, 54, and 123 associations between NES measures and MHRFs for the Ji-12, Yeo-7, and Yeo-17 atlases, respectively, with 89.4% or more validated in non-British populations. For NFC measures, 113, 68, and 185 associations were identified, with 80-83% validated. Effect sizes were consistent across ethnicities, demonstrating generally robust associations (eFigures 31-36, **<u>Supplement 3</u>**).

**Figure 2.**
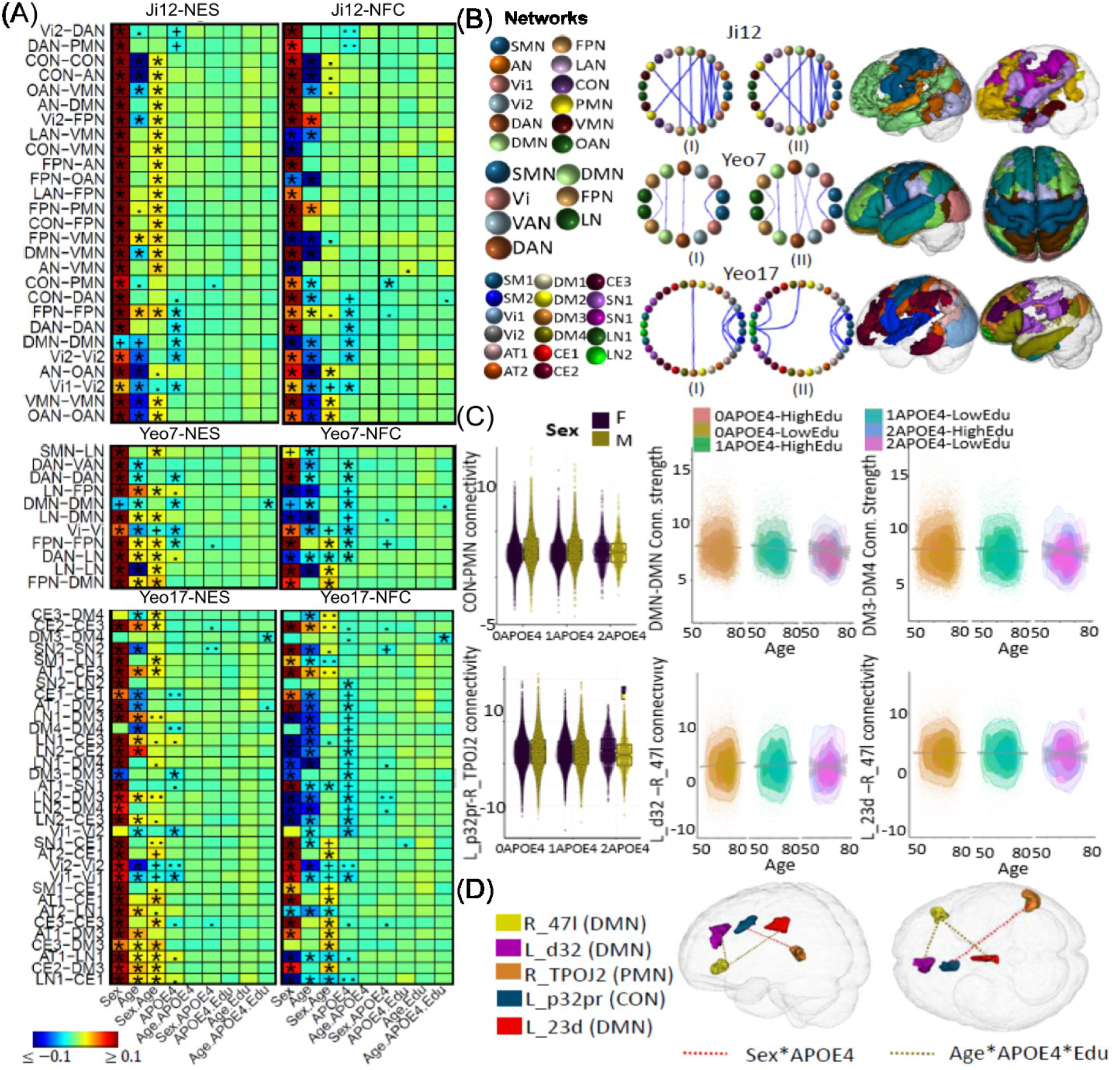
Selected associations of *APOE4* with brain functional network connectivity measures. (A) Heatmaps show association results from the white British population using three network atlases: Ji-12, Yeo-7, and Yeo-17, for network edge strength (NES; I) and functional connectivity (NFC; II). Significant results validated in the non-British population are marked with (*); significant but unvalidated results are indicated by (+). Non-significant results but with p-values *<1e-4* and *<1e-3* are denoted by (.) and (..), respectively. (B) Circular plots showing NES (I) and NFC (II) associations from the three atlases. Network spatial locations shown on the right, with colored spheres representing different networks and red and blue lines for positive and negative associations. (C) Boxplots and scatterplots illustrating *APOE-* ε*4* interaction effects on network and regional connectivity: Row 1 shows sex-*APOE* effect on the CON- PMN NFC (Ji-12) and age-*APOE*-education effects on within-DMN NES (Yeo-7) and DM3-DM4 NES (Yeo-17); Row 2 shows sex-*APOE* effects on connectivity between L_P32pr and R_TPOJ2, and age- *APOE*-education effects on connectivity between Ld32 and R47I, and between L_23d and R_47I. (D) Spatial locations of brain regions for sex-*APOE4* and age-*APOE4*-education interactions.

### Associations between Age, Sex and *APOE* and Brain Functions

Figure 2A shows associations between age, sex, *APOE*, and FC measures. Consistent with previous research ^8,64^, most within- and between-network NES measures decrease with age, except for increases within the FPN and between the FPN-DAN and FPN-VMN networks (eFigure 2, **<u>Supplement 3</u>**). These exceptions align with the posterior-anterior shift in aging (PASA), reflecting increased frontal activity as posterior occipital activity declines ^10,12,13^. Sex differences revealed that males generally exhibited higher NES measures across most networks, except for the DMN, where females had higher connectivity, consistent with prior findings ^65,66^ . Positive age-sex interactions showed that males experienced more pronounced increases in FPN and LN connectivity and larger reductions in visual networks with aging, while females showed stronger reductions in connectivity between the AN, DMN, VMN, and CON networks. No significant effects were observed for the *APOE2* variant, but *APOE4* carriers showed reduced connectivity, particularly within the Vi2, DAN, DMN, and FPN networks, consistent with earlier studies ^17^. A negative sex-*APOE4* interaction (Figure 2C) in the CON-PMN networks (ES: *-0.020*, *p = 5.2E-4*) suggested reduced CON-PMN connectivity in male *APOE4* carriers, offering new insights. Further ROI analysis revealed reduced CON-PMN connectivity in male *APOE4* carriers between L_p32pr and R_TPOJ2 (Figure 2D; ES: *-0.023*, *p = 7.02E-05*).

### Associations between ten MHRFs and Brain Functions

#### Hypertension

Negative effects of hypertension were widespread across networks, including AN, SMN, OAN, VMN, CON, LAN, and DMN and 26 between-network NES measures (Ji-12), supporting its role in accelerating aging ^37,38^. A positive age-hypertension interaction was found on NFC between the FPN and DMN (Figure 3C and eFigure 20, **<u>Supplement 3</u>**) networks (ES: *0.029*, *p = 4.9E-6*). Further post-hoc analyses revealed a positive age-hypertension interaction between L_p9-46v and R_9m (Figure 3D and eFigure 28, **<u>Supplement 3</u>**; ES: *0.029*, *p = 1.2E-5*), indicating reduced anti-correlation, or decreased functional segregation in participants with hypertension during aging ^39,40^.

**Figure 3.**
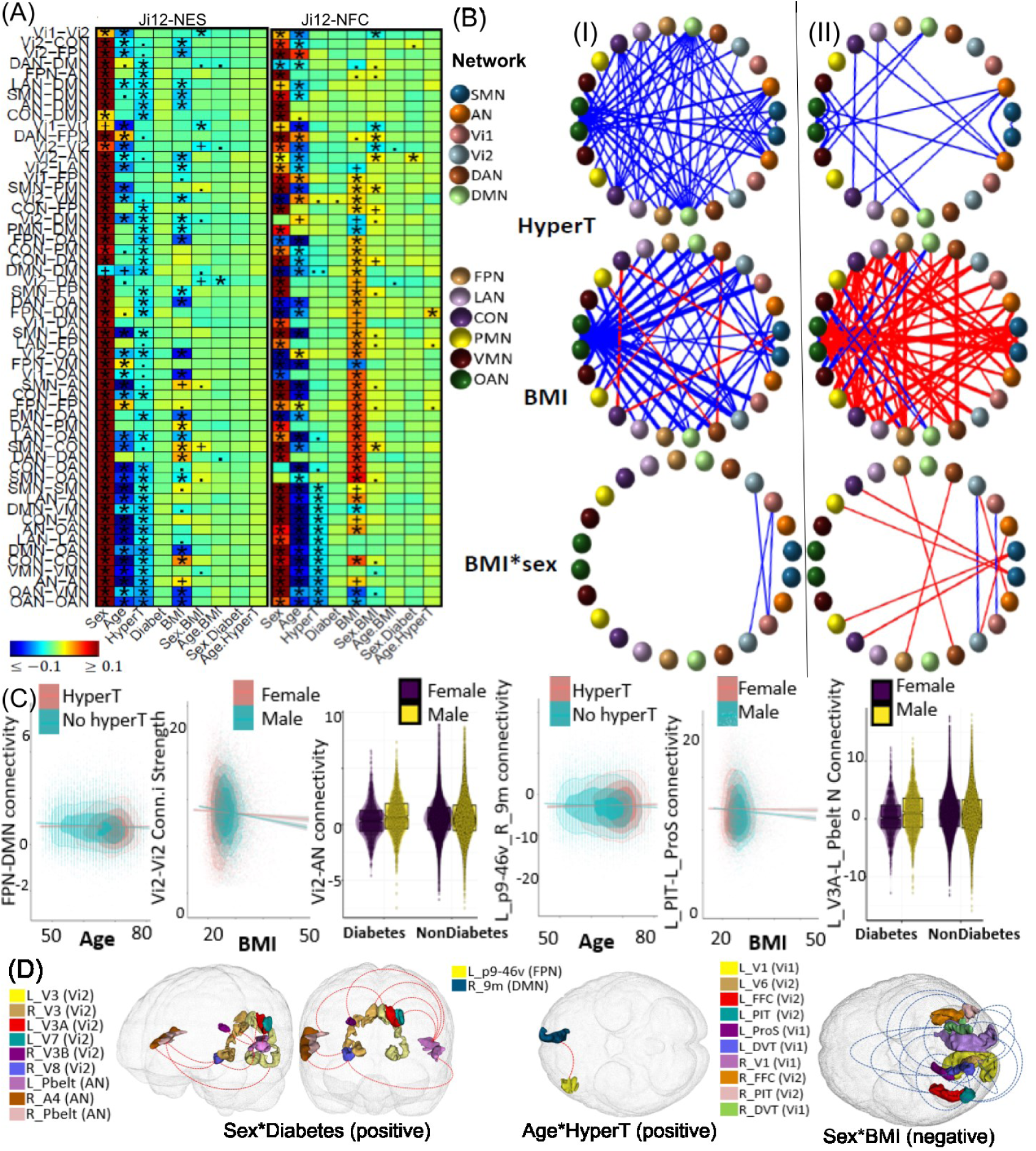
Selected associations of hypertension, BMI, and diabetes with brain functional network connectivity measures. (A) Heatmaps showing association results from the white British population using Ji-12, Yeo-7, and Yeo-17, for network edge strength (NES; I) and network functional connectivity (NFC; II). Significant results validated in the non-British population are marked with (*); significant but unvalidated results are indicated by (+). Non-significant results but with p-values *<1e-4* and *<1e-3* are denoted by (.) and (..), respectively. (B) Circular plots showing NES (I) and NFC (II) associations from the Ji-12 atlas. Colors spheres represent different networks with red and blue lines for positive and negative associations. (C) Scatter plots and boxplots illustrating interaction effects. Columns 1-3: age- hypertension effects on FPN-DMN NFC, sex-BMI effects on Vi2-Vi2 NES, and sex-diabetes effects on Vi2-AN NFC; Columns 4-6: age-hypertension effects on connectivity between L_p9-46v and R_9m, sex-BMI effects on connectivity between L_PIT and L_ProS, and sex-diabetes effects on connectivity between L_V3A and L_Pbelt. (D) Spatial locations of the identified brain regions for the sex-diabetes, age-hypertension, and sex-BMI interactions.

#### BMI

BMI exhibited complex effects on connectivity, with negative effects on NES within the OAN and 19 between-network pairs, and positive effects within the DAN and CON networks (Figures 3A and **3B**, and eFigures 21-23, **<u>Supplement 3</u>**). A negative age-BMI interaction on Vi2-DAN NES suggests that higher BMI accelerates age-related decline in FC (eFigure 49, **<u>Supplement 3</u>**); negative sex-BMI interactions were found on NES within Vi1 (ES: *-0.023*, *p = 1.2E-4*), and between Vi1 and Vi2 networks (ES: *-0.020*, *p = 6.4E-4*), indicating a more rapid decline in visual connectivity strength in males (Figures 3A and **3C**). Further post-hoc analyses revealed 19 ROI-level negative associations in the within-Vi1 and Vi-Vi2 networks (Figures 3C and **3D**, and eFigure 29, **<u>Supplement 3</u>**), 18 of which (94.7%) were validated in non-British populations.

#### Diabetes

Although no main effects of diabetes were observed, a positive sex-diabetes interaction was found on NFC between the Vi2 and AN networks (ES: *0.021*, *p = 5.7E-4*) (Figure 3C). Further post-hoc analyses revealed positive sex-diabetes interaction effects on FCs between L_PBelt and ROIs L_V3A, L_V7, R_V3, R_V8, and R_V3B, and between R_V8 and R_A4 (Figures 3C and **3D** and eFigure 30, **<u>Supplement 3</u>**), highlighting the greater negative impact of diabetes on visual-auditory connectivity in females ^41^.

#### Smoking

Consistent with previous literature ^44–46^, smoking was associated with widespread negative effects, including within FPN and VMN, between VMN and six networks, and between DAN and OAN (Figures 4A and **4B**). However, a positive effect was noted between the CON and AN networks. A negative sex-smoking interaction on NFC between the VMN and CON (ES: -0.020, p = 2.3E-4) suggested greater detrimental effects from smoking in males (Figures 4A and **4C**). ROI-level analyses confirmed these effects between PeEc and pOFC (Figures 4C and **4D**).

**Figure 4.**
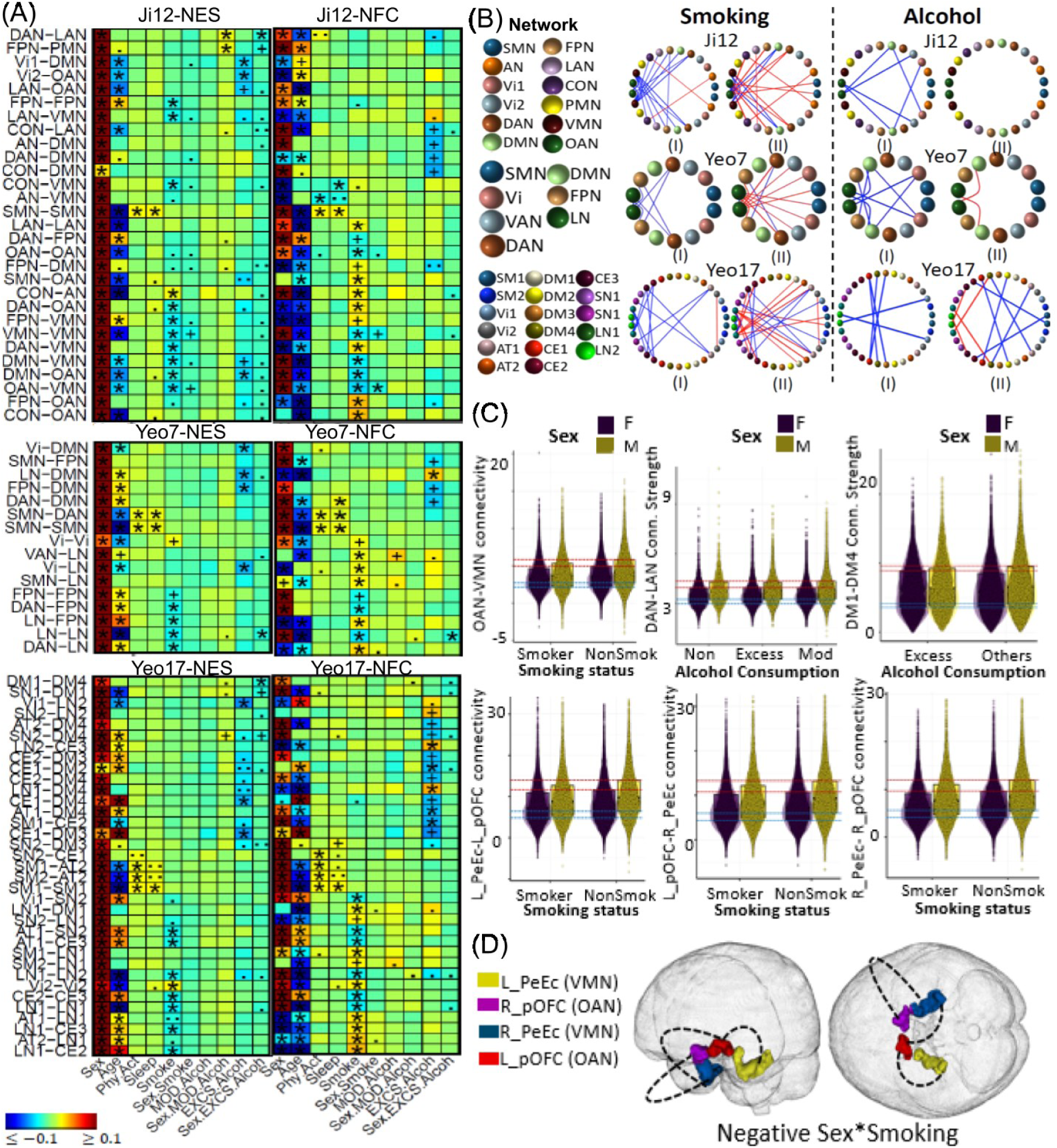
Selected associations of smoking, alcohol consumption, physical activity and sleep with brain functional network connectivity measures. (A) Heatmaps showing association results from the white British population using Ji-12, Yeo-7, and Yeo-17, for network edge strength (NES; I) and network functional connectivity (NFC; II). Significant results validated in the non-British population are marked with (*); significant but unvalidated results are indicated by (+). Non-significant results but with p-values *<1e-4* and *<1e-3* are denoted by (.) and (..), respectively. (B) Circular plots showing NES (I) and NFC (II) associations from the three atlases. Colors spheres represent different networks with red and blue lines for positive and negative associations. (C) Scatter plots and boxplots illustrating interaction effects. Row 1, columns 1-3: the sex-smoking interaction effect on the OAN-VMN between-network NFC, sex-excessive alcohol interaction effect on the DAN-LAN between-network NES, and sex-excessive alcohol interaction effect on the DM1-DM4 between-network NES (Yeo-17 atlas), respectively; row 2, columns 1-3: sex- smoking effects on brain functional connectivities between brain regions L_PeEc and L_pOFC, between L_pOFC and R_PeEc, and between R_9m and R_pOFC, respectively. (D) Spatial locations of the identified brain regions for sex-smoking interactions.

#### Alcohol

Excessive alcohol consumption negatively impacted NES between the DMN-OAN, DMN-Vi1, and OAN-Vi2 networks (Figures 4A and **4B**). A negative sex-excessive alcohol interaction showed a stronger effect in males, particularly between the DAN and LAN networks (ES: -0.021, p = 1.9E-4; Figures 4A, **4C**, and eFigure 25, **<u>Supplement 3</u>**). Conversely, a positive sex-moderate alcohol interaction was observed between the DAN-LAN and FPN-PMN networks, indicating different effects across alcohol consumption levels.

#### Physical activity, Sleep and Education

Positive effects of physical activity and sleep were observed within SMN networks (Figures 4A and **4B**), consistent with previous studies linking sensory/somatomotor network connectivity to sleep quality ^59^. Education was positively associated with NES within the OAN, VMN, and LAN networks, and across 13 network pairs (eFigure 50, **<u>Supplement 3</u>**) A negative sex-education interaction impacted NFC between CON and FPN (ES: *-0.021, p = 1.7E-4*), while a positive interaction enhanced NES between the PMN-DMN networks (ES: *0.023, p = 4.5E-5*), suggesting higher education is associated with greater CON-FPN connectivity in females and PMN-DMN connectivity in males (eFigures 26 and 27, **<u>Supplement 3</u>**).

## DISCUSSION

To our knowledge, this is the largest study to systematically examine the interactions of aging, sex, the *APOE* gene, and MHRFs with brain functional measures. We identified significant sex differences in brain FC associated with factors such as age, *APOE4*, BMI, diabetes, smoking, alcohol consumption, and education. For example, smoking, alcohol, and BMI had more pronounced negative effects in males, while diabetes had a greater impact on females in specific networks. Education was linked to positive effects on CON-FPN networks in females and PMN-DMN networks in males. Additionally, *APOE4*’s effects were more pronounced between the CON and PMN networks in males. These findings reinforce current understandings of sex-specific effects of MHRFs and highlight the need for further explorations.

The sex-age interactions revealed that males experienced greater increases in NES within the FPN and between the DMN-LN and DAN-LN networks, along with larger reductions in the visual network. In contrast, females exhibited greater reductions in connectivity between the AN, DMN, VMN, and CON networks, suggesting distinct aging patterns: males undergoing more extensive reorganization (e.g., PASA) ^10,12,13^ and females demonstrating higher vulnerability to neurodegeneration ^67^. The negative sex- *APOE4* interaction suggests that male carriers are particularly vulnerable, with more pronounced reductions in connectivity between the L_p32pr-R_TPOJ2 regions and between the CON and PMN networks—areas critical for memory and executive function. While much of the literature focuses on the cognitive vulnerability of female *APOE4* carriers ^3,22,25^, our findings suggest that *APOE4* may have a larger impact on the decline of CON-PMN brain networks related to memory and executive functions in men ^24^. The sex-diabetes interaction suggests that diabetes affects brain connectivity differently by sex, with females showing a greater change in visual-auditory connectivity ^41^. A negative sex-smoking interaction was observed between the VMN and CON networks, indicating more severe smoking-related impairments in males. Similarly, the negative sex-excessive alcohol interaction showed a larger detrimental effect between the DAN and LAN networks in males, likely due to alcohol misuse leading to lower cortical volume, reduced white matter and hippocampal volume, and greater changes in brain function and behavior in men ^68–70^. The sex-BMI interaction highlights a larger impact of obesity on brain function in males, aligning with previous research that suggests men experience detrimental changes in brain connectivity starting from the overweight category, while women typically show declines only in the obese range, possibly due to obesity-induced chronic white matter damage in males ^71^.

Our findings on excessive alcohol consumption corroborate previous research, indicating that reduced connectivity in the precuneus, postcentral gyrus, insula, visual cortex, and left executive control network are key areas of rsfMRI NC reduction ^50,51^. Meanwhile, our study highlights a sex-specific dichotomy between excessive and moderate alcohol consumption on brain connectivity. In males, excessive alcohol had more detrimental effects between the DAN and LAN, while moderate consumption showed a protective effect on connectivity between the DAN and LAN, and FPN and PMN networks, suggesting a U-shaped nonlinear patterns between alcohol dose and brain health ^72^.

We observed that higher education positively influences connectivity within and between networks including the OAN, VMN, LAN, and FPN, potentially enhancing cognitive reserve ^53,54^. Sex-education interactions revealed that with higher education, females showed increased connectivity in the CON-FPN network, while males benefited more in the PMN-DMN network. This suggests that education may enhance neural pathways aligned with the cognitive needs of each sex, indicating the potential for tailored educational programs to optimize brain health and cognitive function.

Elevated blood pressure, a common cardiovascular risk factor, is linked to cognitive decline in later life. Hypertension broadly impairs connectivity within and between attentional, SMN, and DMN networks, likely due to its impact on neural inefficiencies ^73^. We observed reduced anti-correlation between the dorsolateral (L_p9-46v) and medial (R_9m) prefrontal cortices, suggesting decreased functional segregation. A recent study ^39^ demonstrated that higher blood pressure causally reduces brain functional segregation and worsening cognition in the aging population through observational and Mendelian randomization analyses.

### Strength and Limitations

Leveraging large-sample fMRI data from the UKB, we investigated a broad range of modifiable and non-modifiable risk factors, exploring their joint, conditional, and interaction effects on brain functions. These findings were validated across multiple atlases and ethnic populations, ensuring robustness against racial differences, atlas choices, outlier sensitivities, and sample size limitations. Our main analyses were based on parcellation-based full correlations. Although FMRIB’s ICA-based X-noiseifier (FIX) has been applied to the UKB dataset to remove scanner artifacts and motion effects, full correlation measures can be sensitive to remaining global artifacts ^74^, while measuring partial functional connectivity between paired brain regions can reduce global artifacts and remove dependencies on other brain regions ^75^. Future studies will explore parcellation-based partial correlation traits.

## CONCLUSION

Our study revealed sex differences in the effects of *APOE4* and MHRFs on brain FC measures, with male *APOE4* carriers experiencing larger declines in FC between the CON and PMN networks. Distinct aging patterns emerged, where males showed more neural reorganization, and females demonstrated greater vulnerability to neurodegeneration. Sex-specific effects of diabetes, smoking, BMI, and education underscore the importance of considering sex and demographic factors in brain health research.

Additionally, we observed a sex-specific dichotomy in the impact of alcohol consumption: excessive drinking reduces, while moderate consumption increases functional connectivity strength between dorsal attention and language networks in males.

## Data Availability

All data produced in the present study are available upon reasonable request to the authors.

## Author Contributions

Concept and design: Zhu, Wu and Li; Acquisition, analysis, or interpretation of data: Li and Chen; Drafting of the manuscript: Li and Chen; Critical revision of the manuscript for important intellectual content: Li, Chen, Wu, Zhu, Zhao, Giovanello and Garden; Statistical analysis: Chen and Li; Obtained funding: Wu and Zhu

## Data Sharing Statement

The individual-level data used in this study can be obtained from https://www.ukbiobank.ac.uk/.

