## Supplement for "The Interaction Effects of Sex, Age, APOE and Common Health Risk Factors on Human Brain Functions"

This PDF file includes:

Supplement 1. Supplementary Method
Supplement 2. eTables S1 to S6
Supplement 3. eFigures S1 to S50

### Supplement 1. Supplementary Method

#### Imaging Data Information

All UKB brain imaging datasets, including the resting-state fMRI (rsfMRI) and the T1-weighted structural MRI data were acquired from standard Siemens Skyra 3T scanners. The rsfMRI data were acquired using a blood oxygen level-dependent (BOLD) sequence and an echo-planar imaging (EPI) sequence (TR = 0.735 s, TE = 39 ms, FoV = 88 × 88 × 64, voxel resolution 2.4 mm × 2.4 mm × 2.4 mm, a multiband factor of 8, no iPAT, flip angle 52◦, and fat saturation), lasted approximately 6 minutes. The echo time (TE) and repetition time (TR) were 39 ms and 735 ms, respectively. In addition, the T1-weighted sMRI scans were acquired at the isotropic resolution of 1 mm and a dimension of 208×256×256 matrix, with a TR of 2000 ms, an inversion time of 880 ms, a TE of 2 ms, an in-plane acceleration of 2, and a scan time of 5 min using straight sagittal orientation.

#### Imaging Data Processing

Our imaging analyses followed the rsfMRI processing workflow provided by the UKB [1] (https://git.fmrib.ox.ac.uk/falmagro/UK_biobank_pipeline_v_1), which includes motion correction, grand-mean intensity normalization, high-pass temporal filtering, EPI unwarping, gradient distortion correction (GDC), Independent Component Analysis (ICA)-based X-noiseifier (FIX) artifact removal. Structural artifacts were removed using FMRIB's ICA-FIX processing [2–4]. The GDC-corrected rsfMRI data were co-registered with the high-resolution T1 MRI image, which were then registered to the MNI-152-2mm standard space. Subjects with unusable, low-quality T1 MRI scans, as identified by the UKB brain imaging team [1] were excluded in our analyses. Meanwhile, the head motion parameters (UKB data field 25741) were calculated by averaging the framewise displacement (FD) across the brain for each consecutive pair of time points for each rsfMRI scan, which were then averaged across all time points, which were included as confounding covariates in our association analyses.

The regions of interest (ROIs) used to construct the network imaging-derived phenotypes (IDPs) were selected as HCP-MMP atlas [5], corresponding to 360 cortical regions classified into twelve resting-state networks following the Ji-12 network atlas [6], namely, the somatomotor (SMN), auditory (AN), visual1 (Vi1), visual2 (Vi2), dorsal attention (DAN), default mode (DMN), frontoparietal (FPN), language (LAN), cingulo-opercular (CON), posterior multimodal (PMN), ventral multimodal (VMN), and orbito-Affective (OAN) networks. For each rsfMRI scan, the mean time series from each of 360 ROIs were extracted and the correlation between each pair of regional time series was transformed from Pearson correlations (r-values) to z-statistics using Fisher transformation. This yielded a 360 × 360 FC matrix for each subject in the datasets, representing 64,620 connectivity traits, using the FSLNets (http://fsl.fmrib.ox.ac.uk/fsl/fslwiki/FSLNets) toolbox. Next, for each pair of the 12 networks we calculated the between- and within-network functional connectivity (NFC) and edge strength (NES) measures as follows,

$$Between\text{–}network\ NFC(I1, I2) = \sum_{i1 \in I1, i2 \in I2} FC(i1, i2)/[\#I1\#I2]$$

$$Between\text{–}network\ NES(I1, I2) = \sum_{i1 \in I1, i2 \in I2} |FC(i1, i2)|/[\#I1\#I2],$$

$$Within\text{–}network\ NFC(I) = \sum_{i1, i2 \in I, i1 \neq i2} FC(i1, i2) * 2/[\#I(\#I - 1)],$$

$$Within\text{–}network\ NES(I) = \sum_{i1, i2 \in I, i1 \neq i2} |FC(i1, i2)| * 2/[\#I(\#I - 1)],$$

where $I$, $I1$ and $I2$ are the networks of interest, $FC(i1, i2)$ is the regional FC calculated between two ROIs $i1$ and $i2$, and $\#I$ denotes the number of ROIs in the network $I$. We also generated 100 × 100 FC matrices based on Schaefer 100 parcellation atlas and network-level traits for the Yeo-7 and Yeo-17 atlases [7]. For each of the FC, NFC and NES traits, subjects with extreme deviations—defined as values exceeding five times the median absolute deviation from the population median—were flagged as outliers. These outlier data points were excluded from the subsequent association analyses.

#### APOE Genotyping

The *APOE* gene, particularly the ε4 allele, is acknowledged as a significant risk factor for Alzheimer's disease (AD). Determination of the *APOE* genotype relies on two single nucleotide polymorphisms (SNPs), rs429358 and rs7412, which define three alleles—*APOE-ε2*, *APOE-ε3*, and *APOE-ε4*—and result in six possible genotypes. These genotypes are: *APOE-ε2/ε2*, *APOE-ε2/ε3*, *APOE-ε2/ε4*, *APOE-ε3/ε3*, *APOE-ε3/ε4*, and *APOE-ε4/ε4*. *APOE-ε2* is associated with a decreased risk of AD and is the rarest allele, whereas *APOE*-ε3, the most common allele, is considered neutral in terms of risk. *APOE-ε4*, conversely, is associated with an increased risk of AD [8].

Utilizing imputed SNP data from UKB resources, *APOE* genotyping was conducted on the 36,630 individuals. The genotype distribution is as follows: *APOE-ε4/ε4* is present in 806 subjects (2.2%), *APOE-ε2/ε2* in 211 subjects (0.6%), and *APOE-ε3/ε3* in 21,688 subjects (59.2%). Additionally, 4,545 subjects (12.4%) possess the *APOE-ε2/ε3* genotype, 877 subjects (2.4%) have *APOE-ε2/ε4*, and 8,503 subjects (23.2%) are *APOE-ε3/ε4*. Gender-wise distribution among these genotypes is detailed, with allele frequencies for *ε2*, *ε3*, and *ε4* being 8.0%, 77.1%, and 14.9%, respectively. Demographics for each genotype group of British and Non British participants are outlined in eTable 4 and 5, **Supplement 2**. Our analysis incorporated the count of the APOE-ε4 and APOE-ε2 alleles as individual covariates in statistical models. This approach allows for the assessment of the additive effects attributed to each allele. It differs from that of some prior studies which grouped individuals with the *APOE-ε4* allele into homozygotes and all others, or classified individuals as *APOE-ε2* carriers if they were homozygous for *APOE-ε2* or had *APOE-ε2/ε3* genotype, considering the remaining as controls.

#### Lifestyle and SES Factors

In our study, the selection of lifestyle variables followed the criteria established in the research on healthy lifestyle and dementia by Lourida et al. (2019) [9]. These variables, collected via touchscreen questionnaires during the UKB baseline assessment, include smoking status, physical activity, diet, and alcohol consumption—factors known to be associated with brain health. Specifically, we opted to use ever-smoking status (encompassing both ever-smokers and never-smokers) instead of current smoking status. This approach allows us to include a larger proportion of cases, thereby enhancing our statistical power. Regular physical activity, as per the American Heart Association (AHA) guidelines, is defined as engaging in at least 150 minutes of moderate exercise or 75 minutes of vigorous activity weekly. The diet was considered healthy if participants met at least four of the seven cardiometabolic health dietary recommendations outlined by Mozaffarian (2016) [10], which include the intake of vegetables, fruits, fish, unprocessed meat, processed meat, whole grains, and refined grains. Alcohol consumption was stratified into abstinence, moderate consumption, and excessive consumption. This categorization is due to the mixed evidence regarding the effects of moderate drinking, as highlighted by studies from Daviet et al. (2022) [11] and Sabia et al. (2018) [12]. Moderate alcohol consumption is capped at 14 grams per day for women and 28 grams per day for men, while abstinence encompasses individuals who do not drink or only consume alcohol on special occasions, amounting to zero units per day. Additionally, we have considered sleep duration as a lifestyle variable, given its association with dementia risk as described by Palpatzis et al. (2022) [13], categorizing short and long sleep duration as less than 6 hours or more than 8 hours, respectively. Beyond these lifestyle factors, our analysis also incorporated SES elements such as education level and social deprivation. Education level was divided into individuals with or without a college or university degree or higher. Social deprivation was measured using the Townsend deprivation (SoDep) index based on each participant's postcode, which we dichotomized at the median of the index into two groups: those above and those below the median. The methodology for generating these lifestyle and SES factors, along with a more detailed discussion, can be found in the eTable 1, **Supplement 2**.

#### CVRFs

Cardiovascular diseases (CVDs) are the leading cause of mortality worldwide, highlighting the critical need for a deeper understanding of risk factors to alleviate the growing health burden. Several lifestyle factors, including ever-smoking and alcohol consumption, are also recognized as CVRFs. In addition to the above lifestyle factors, diabetes, body mass index (BMI), and hypertension are three large CVRFs that contribute to the development and progression of CVD, which are the primary focus of our analyses. Diabetes mellitus, characterized by chronic hyperglycemia, not only damages the vascular endothelium but also intensifies the complexity of atherosclerotic plaque, thereby significantly increasing cardiovascular risk; obesity, measured by the a high body mass index (BMI), is another critical risk factor that frequently coexists with metabolic abnormalities, leading to atherosclerotic cardiovascular disease. Furthermore, hypertension is a silent yet formidable risk factor that plays a substantial role in most cardiovascular events. It imposes an excessive burden on arterial walls, leading to structural and functional impairments of the vasculature. In our analysis, BMI was treated as a continuous variable (UKB data field 21001), whereas diabetes and hypertension were categorized as binary disease status variables, based on their presence in medical records according to the International Classification of Diseases, 10th Revision (ICD-10; UKB data-Field 41270).

#### Confounding Covariates

In our analysis of the effects of the risk factors, including the APOE gene, lifestyle factors, SES, and CVRFs, we controlled for the following confounding variables: study phase of participants, age at the time of imaging, age squared, sex, interactions between sex and age, interactions between sex and age squared, partnership status (whether living with a partner; UKB data field 6141), head motion, brain volume scaling (UKB data field 25000), and brain positioning within the scanning field (UKB data fields 25756, 25757, and 25758).

#### Missing data

When we ran multiple regression models, data with missing responses (functional connectivities) were dropped automatically when we fitted the regression model. For covariates, all variables (age, sex, APOE, MHRFs) have less than 1% missing rate of the data except the marriage status (living with partners) and the education level. Approximately 23.4% of subjects do not have partner information and approximately 6.9% of subjects do not have education information. We imputed these missing values with the mode.

#### Additional Findings

For association results between *APOE*, results from Yeo-7 atlas exhibited increased between-network connectivity strength with aging, including DMN-DAN, DAN-LN, and LN-DMN. Using the Yeo-7 atlas, we identified the negative age-*APOE4*-education effects (**Figure 2C**) on the NFC within the DMN (ES: *-0.019*, $p = 1.0E\text{-}3$). In particular, in *APOE4* carriers with high education, the within-DMN NFC decreases faster during aging, indicating that the protective effects of education may diminish in *APOE4* carriers [14]. No ROI-level associations passed the Bonferroni significance level *0.05/m*; however, two associations exceeded $p < 1E\text{-}4$ threshold between R_47l and L_d32 (ES: *-0.026*, $p = 2.0E\text{-}5$) and between R_47l and L_23d (ES: *-0.025*, $p = 4.9E\text{-}5$) (**Figure 2D**). Based on Yeo-17 atlas, a positive age-sex-sleep three-way interaction term was found between the LN2 and CE3 networks, suggesting that the anticorrelation between LN2 and CE3 networks anticorrelation increases faster during aging in males with < 7 hours of sleep (eFigure 24, **Supplement 3**)

### Supplement 2. Supplementary tables

**eTable 1. Criteria for the ten modifiable health-related risk factors generated from the UKB data fields.** All variables are measured at the baseline.

| Variable | UK Biobank field ID | Defination | Details |
|---|---|---|---|
| Education | 6138 | 2 categories:<br>(1) Having college or university degree<br>(0) Not having college or univerisity degree | A levels/AS levels, O levels/GCSEs, CSEs, NVQ, HND, HNC or other professional qualifications, such as nursing; teaching and equivalents are considered as not having such degree |
| Social deprivation | 189 | Townsend deprivation index<br>(1) Above median (0) Below median | Median is caculated from the 500K UKB participants |
| Doing regular physical activity | 884, 894, 904, 914 | 2 catergories:<br>(1) Doing regular physical activity<br>(0) Not doing regular physical activity | Meeting any of the 5 criteria is considered regular physical activity (AHA recommendations):<br>1. Doing > 150 minutes of moderate activity each week<br>2. Doing > 75 minutes of rigorous activity each week<br>3. Equivalent combinations of moderate and rigorous activity (2 minutes of moderate eqauls 1 minutes of rigorous activity)<br>4. Doing moderate activity at least 5 days a week |
| Healthy diet | 1309, 1319, 1289, 1299, 1329, 1339, 1349, 1369, 1379, 1389, 1438, 1448, 1458, 1468 | 2 catergories:<br>(1) Meeting at least 4 of the 7 healthy eating criteria<br>(0) Only meeting 3 or less of the 7 healthy eating criteria | 7 healthy eating criteria:<br>1. Eating >= 3 servings of fruits a day<br>2. Eating >= 3 servings of vegetables a day<br>3. Eating >= 2 servings of vegetables a week<br>4. Eating >= 3 servings of whole grains a day<br>5. Eating <= 1.5 servings of refined grains a day<br>6. Eating <= 1 serving of processed meats a week<br>7. Eating <= 1.5 servings of unprocessed red meats a week |
| Alcohol consumption | 20117, 1558, 1568, 1578, 1588, 1598, 1608, 5364, 4407, 4418, 4429, 4440, 4451, 4462 | 3 catergories:<br>(2) Drinking excessive alcohol<br>(1) Drinking moderate alcohol<br>(0) Not drinking at all | Up to 1 drink/day for women and up to 2 drinks/ day for men are considered moderate alcohol; more than the above threshold is considered excessive alcohol ( 2015-2020 US dietary guidelines for American). 14g pure alcohol is considered 1 drink, 1 glass of wine (125ml) is equivalent to 0.85 drink, 1 glass of fortified wine (50ml) is equivalent to 0.56 drink, 1 shot of spirits (25ml) is equivalent to 0.57 drink and 1 |
| Sleep | 1160 | (1) Sleeping between 6-8 hours a day<br>(0) Sleeping outside 6-8 hours a day | According to previous studies, sleeping too long or too short are both associated with higher risks of dementia |
| Smoking Status | 20116 | (1) Ever had smoked<br>(0) Never smoked | (1) include current smokers and previous smokers |
| Hypertension Status | 20002 | (1) Having hypertension<br>(0) Never expreienced hypertension | Including essential hypertension4895 and gestational hypertension/pre-eclampsia |
| Diabete | 120007 | (1) Ever had Diabete<br>(0) Never had Diabete | Including both Type I and Type II |
| BMI | 21001 | Continuous variable with mean around 27 | Units of measurement are Kg/m2. |

**eTable 2. Models investigated in this study.** Each interaction term is tested separately, while controlling main effects of all risk factors, head motion, brain position, volumetric scaling, study site, and phase (age-squared and sex-age-squared interaction were also controlled for when sex, age, or sex-age was not a term of interest) as confounding factors. “A:B” denotes the interaction of covariates A and B; “A:B:C" denotes the three-way interaction of covariates A, B, and C. A three-way interaction term implied that all its relevant two-way interactions and main effects were also included; a two-way interaction term implied that all relevant main effects were also included. ROI, regions of interest; MHRF, modifiable health-related risk factors.

| **Models Investigated** | **Terms of interest** | **Models** |
|---|---|---|
| (M0) Main effect model | Age,Sex,APOE4,APOE2,Smoking_status,hyperten_status,Diabete,BMI,Edu,SoDep,Regular_physical_activity,Healthy_diet,Sleep_7,MOD.alcohol,EXCS.alcohol | 1) For effects of Sex and Age:<br>fMRI ~ Sex + Age+ APOE4+APOE2+MHRFs+head_motion+brain_position+volumetric_scaling+Partner+UKB_Phase+site;<br>2) For all other effects:<br>fMRI ~ APOE4+APOE2+MHRFs + head_motion+brain_position+volumetric_scaling+Partner+UKB_Phase+site+Age+Sex+Age:Sex+(Age^2)+(Age^2):Sex |
| (M1) Two-way effect model | Age:Smoking_status,Age:hyperten_status,Age:Diabete,Age:BMI,Age:Edu,Age:SoDep,Age:Regular_physical_activity,Age:Healthy_diet,Age:Sleep_7,Age:MOD.alcohol,Age:EXCS.alcohol,Sex:Smoking_status,Sex:hyperten_status,Sex:Diabete,Sex:BMI,Sex:Edu,Sex:SoDep,Sex:Regular_physical_activity,Sex:Healthy_diet,Sex:Sleep_7,Sex:MOD.alcohol,Sex:EXCS.alcohol,APOE4:Smoking_status,APOE4:hyperten_status,APOE4:Diabete,APOE4:BMI,APOE4:Edu,APOE4:SoDep,APOE4:Regular_physical_activity,APOE4:Healthy_diet,APOE4:Sleep_7,APOE4:MOD.alcohol,APOE4:EXCS.alcohol,APOE2:Smoking_status,APOE2:hyperten_status,APOE2:Diabete,APOE2:BMI,APOE2:Edu,APOE2:SoDep, APOE2:Regular_physical_activity,APOE2:Healthy_diet,APOE2:Sleep_7,APOE2:MOD.alcohol, APOE2:EXCS.alcohol,Age:Sex,Sex:APOE4,Age:APOE4,Sex:APOE2,Age:APOE2 | 3) For effects of Sex and Age:<br>fMRI ~ Sex*Age + APOE4+APOE2+MHRFs+head_motion+brain_position+volumetric_scaling+Partner+site;<br>4) For all other effects:<br>fMRI ~ A:B + APOE4+APOE2+SES+MHRFs +head_motion+brain_position+volumetric_scaling+Partner+UKB_Phase+site+Age:Sex+(Age^2):Sex, where A:B represent interaction terms between risk factors A and B, looping over Age:APOE, Sex:APOE, Age:MHRF, Sex:MHRF, APOE:MHRF interactions. |
| (M2) Three-way effect model | Age:Sex:Smoking_status,Age:Sex:hyperten_status,Age:Sex:Diabete,Age:Sex:BMI,Age:Sex:Edu,Age:Sex:SoDep,Age:Sex:Regular_physical_activity,Age:Sex:Healthy_diet,Age:Sex:Sleep_7,Age:Sex:MOD.alcohol,Age:Sex:EXCS.alcohol,Sex:APOE4:Smoking_status,Sex:APOE4:hyperten_status,Sex:APOE4:Diabete,Sex:APOE4:BMI,Sex:APOE4:Edu,Sex:APOE4:SoDep,Sex:APOE4:Regular_physical_activity,Sex:APOE4:Healthy_diet,Sex:APOE4:Sleep_7,Sex:APOE4:MOD.alcohol,Sex:APOE4:EXCS.alcohol,Age:APOE4:Smoking_status,Age:APOE4:hyperten_status,Age:APOE4:Diabete,Age:APOE4:BMI,Age:APOE4:Edu,Age:APOE4:SoDep,Age:APOE4:Regular_physical_activity,Age:APOE4:Healthy_diet,Age:APOE4:Sleep_7,Age:APOE4:MOD.alcohol,Age:APOE4:EXCS.alcohol,Age:Sex:APOE4,Age:Sex:APOE2 | 5) fMRI ~ A:B:C+APOE4+APOE2+MHRFs+ head_motion+brain_position+volumetric_scaling+Partner+UKB_Phase+site+Age:Sex+(Age^2):Sex, where A:B:C represent interaction terms between risk factors A, B and C, looping over Age:APOE:MHRF, Age:Sex:APOE, Age:Sex:MHRF, Sex:APOE:MHRF interactions. |

**eTable 3. A summary of basics and results for functional connectivity measures in this study.** Those include the number of signals (sex, age and sex*age not included; associations that pass the corresponding Bonferroni significance level 0.05/*m* in white British) and the number of signals that were validated by non-British populations (with the same directions as in white British). NFC: network-level mean functional connectivity; NES: network-level edge strength.

| FC Measures | Network | Phenotype | Number of connectivities *m* | Number of Signals | Number of Signals validated by non-British |
|---|---|---|---|---|---|
| Ji12-RestConn | Ji-12 | NFC | 66 (between) + 12 (within) =78 | 113 | 91 (80.5%) |
| Ji12-Abs-RestConn | Ji-12 | NES | 66 (between) + 12 (within) =78 | 113 | 102 (90.3%) |
| Yeo7-RestConn | Yeo-7 | NFC | 21 (between) + 7 (within) =28 | 68 | 55 (80.9%) |
| Yeo7-Abs-RestConn | Yeo-7 | NES | 21 (between) + 7 (within) =28 | 54 | 49 (90.7%) |
| Yeo17-RestConn | Yeo-17 | NFC | 136 (between) + 17 (within) =153 | 185 | 154 (83.2%) |
| Yeo17-Abs-RestConn | Yeo-17 | NES | 136 (between) + 17 (within) =153 | 123 | 110 (89.4%) |

**eTable 4. Demographic information, *APOE* gene status, and modifiable health-related risk factors of white British UKB subjects.** Percentiles may not add up to 1 due to missingness.

| Characteristic | Number(%) APOE4 homozygote (N=760) | APOE3 homozygote (N=19943) | APOE2 homozygote (N=196) | APOE4/APOE3 (N=7885) | APOE2/APOE3 (N=4209) | APOE4/APOE2 (N=831) | Total (N=33824) |
|---|---|---|---|---|---|---|---|
| Age at brain imaging | | | | | | | |
| Mean(sd) | 63.2 (7.24) | 63.9 (7.51) | 63.4 (7.85) | 63.4 (7.46) | 64.0 (7.47) | 63.9 (7.58) | 63.8 (7.49) |
| Median(range) | 64.0 [48.0, 80.0] | 65.0 [45.0, 81.0] | 63.5 [46.0, 80.0] | 64.0 [46.0, 81.0] | 65.0 [46.0, 81.0] | 64.0 [47.0, 80.0] | 64.0 [45.0, 81.0] |
| Sex | | | | | | | |
| Female | 419 (55.1%) | 10402 (52.2%) | 103 (52.6%) | 4244 (53.8%) | 2234 (53.1%) | 459 (55.2%) | 17861 (52.8%) |
| Male | 341 (44.9%) | 9541 (47.8%) | 93 (47.4%) | 3641 (46.2%) | 1975 (46.9%) | 372 (44.8%) | 15963 (47.2%) |
| Marriage status | | | | | | | |
| Living with a partner | 587 (77.2%) | 15104 (75.7%) | 151 (77.0%) | 5959 (75.6%) | 3186 (75.7%) | 614 (73.9%) | 25601 (75.7%) |
| Not living with a partner | 32 (4.2%) | 885 (4.4%) | 12 (6.1%) | 382 (4.8%) | 176 (4.2%) | 48 (5.8%) | 1535 (4.5%) |
| Education | | | | | | | |
| College/university degree or above | 360 (47.4%) | 9011 (45.2%) | 82 (41.8%) | 3537 (44.9%) | 1878 (44.6%) | 389 (46.8%) | 15257 (45.1%) |
| No college/university degree or above | 355 (46.7%) | 9493 (47.6%) | 100 (51.0%) | 3823 (48.5%) | 2015 (47.9%) | 378 (45.5%) | 16164 (47.8%) |
| Social Deprivation | | | | | | | |
| Above the Townsend deprivation index median | 291 (38.3%) | 8164 (40.9%) | 74 (37.8%) | 3248 (41.2%) | 1647 (39.1%) | 325 (39.1%) | 13749 (40.6%) |
| Below the Townsend deprivation index median | 469 (61.7%) | 11762 (59.0%) | 122 (62.2%) | 4631 (58.7%) | 2557 (60.8%) | 506 (60.9%) | 20047 (59.3%) |
| Lifestyle factors | | | | | | | |
| Smoking status | | | | | | | |
| Ever smoked | 467 (61.4%) | 12201 (61.2%) | 125 (63.8%) | 4822 (61.2%) | 2550 (60.6%) | 536 (64.5%) | 20701 (61.2%) |
| Never smoked | 293 (38.6%) | 7742 (38.8%) | 71 (36.2%) | 3063 (38.8%) | 1659 (39.4%) | 295 (35.5%) | 13123 (38.8%) |
| Doing regular physical activity | | | | | | | |
| Yes | 575 (75.7%) | 14964 (75.0%) | 144 (73.5%) | 5869 (74.4%) | 3174 (75.4%) | 604 (72.7%) | 25330 (74.9%) |
| No | 183 (24.1%) | 4861 (24.4%) | 52 (26.5%) | 1962 (24.9%) | 1015 (24.1%) | 221 (26.6%) | 8294 (24.5%) |
| Healthy diet | | | | | | | |
| Yes | 397 (52.2%) | 9693 (48.6%) | 86 (43.9%) | 3896 (49.4%) | 1997 (47.4%) | 397 (47.8%) | 16466 (48.7%) |
| No | 363 (47.8%) | 10247 (51.4%) | 110 (56.1%) | 3985 (50.5%) | 2211 (52.5%) | 434 (52.2%) | 17350 (51.3%) |
| Alcohol consumption | | | | | | | |
| Excessive alcohol consumption | 75 (9.9%) | 1989 (10.0%) | 27 (13.8%) | 826 (10.5%) | 433 (10.3%) | 88 (10.6%) | 3438 (10.2%) |
| Moderate alcohol consumption | 478 (62.9%) | 12213 (61.2%) | 112 (57.1%) | 4743 (60.2%) | 2557 (60.8%) | 525 (63.2%) | 20628 (61.0%) |
| Not current drinking | 207 (27.2%) | 5735 (28.8%) | 57 (29.1%) | 2315 (29.4%) | 1218 (28.9%) | 218 (26.2%) | 9750 (28.8%) |
| Sleeping between 6 hours and 8 hours | | | | | | | |
| Yes | 338 (44.5%) | 8640 (43.3%) | 89 (45.4%) | 3535 (44.8%) | 1754 (41.7%) | 380 (45.7%) | 14736 (43.6%) |
| No | 420 (55.3%) | 11265 (56.5%) | 107 (54.6%) | 4339 (55.0%) | 2442 (58.0%) | 449 (54.0%) | 19022 (56.2%) |
| CVRFs factors | | | | | | | |
| Hypertension Status | | | | | | | |
| Yes | 148 (19.5%) | 4092 (20.5%) | 55 (28.1%) | 1487 (18.9%) | 826 (19.6%) | 160 (19.3%) | 6768 (20.0%) |
| No | 612 (80.5%) | 15851 (79.5%) | 141 (71.9%) | 6398 (81.1%) | 3383 (80.4%) | 671 (80.7%) | 27056 (80.0%) |
| Diabetes | | | | | | | |
| Yes | 23 (3.0%) | 863 (4.3%) | 8 (4.1%) | 274 (3.5%) | 180 (4.3%) | 42 (5.1%) | 1390 (4.1%) |
| No | 737 (97.0%) | 19080 (95.7%) | 188 (95.9%) | 7611 (96.5%) | 4029 (95.7%) | 789 (94.9%) | 32434 (95.9%) |
| BMI | | | | | | | |
| Mean(sd) | 26.2 (3.88) | 26.5 (4.18) | 26.8 (4.72) | 26.6 (4.23) | 26.7 (4.22) | 26.7 (4.39) | 26.6 (4.20) |
| Median(range) | 25.9 [16.4, 44.4] | 26.0 [15.2, 56.6] | 26.2 [18.0, 45.3] | 26.0 [14.7, 56.0] | 26.2 [16.2, 50.5] | 25.9 [18.0, 44.7] | 26.0 [14.7, 56.6] |

*: Percentiles do not add up to 1 is due to missingness.

*:All variables are measured at the baseline except the age at imaging, hypertension status, and diabetes.

**eTable 5. Demographic information, *APOE* gene status, and modifiable health-related risk factors of non-British UKB subjects.** Percentiles may not add up to 1 due to missingness.

| | Number(%) | | | | | | |
|---|---|---|---|---|---|---|---|
| **Characteristic** | **APOE4 homozygote (N=46)** | **APOE3 homozygote (N=1745)** | **APOE2 homozygote (N=15)** | **APOE4/APOE3 (N=618)** | **APOE2/APOE3 (N=336)** | **APOE4/APOE2 (N=46)** | **Total (N=2806)** |
| Age at brain imaging | | | | | | | |
| Mean(sd) | 60.9 (7.63) | 61.9 (7.89) | 62.7 (9.18) | 61.6 (7.65) | 62.0 (8.13) | 61.5 (7.81) | 61.8 (7.86) |
| Median(range) | 60.5 [48.0, 79.0] | 62.0 [44.0, 81.0] | 63.0 [48.0, 78.0] | 61.0 [47.0, 78.0] | 62.0 [45.0, 80.0] | 61.5 [50.0, 79.0] | 62.0 [44.0, 81.0] |
| Sex | | | | | | | |
| Female | 23 (50.0%) | 932 (53.4%) | 10 (66.7%) | 358 (57.9%) | 187 (55.7%) | 24 (52.2%) | 1534 (54.7%) |
| Male | 23 (50.0%) | 813 (46.6%) | 5 (33.3%) | 260 (42.1%) | 149 (44.3%) | 22 (47.8%) | 1272 (45.3%) |
| Marriage status | | | | | | | |
| Living with a partner | 31 (67.4%) | 1233 (70.7%) | 13 (86.7%) | 410 (66.3%) | 220 (65.5%) | 24 (52.2%) | 1931 (68.8%) |
| Not living with a partner | 4 (8.7%) | 128 (7.3%) | 1 (6.7%) | 53 (8.6%) | 28 (8.3%) | 6 (13.0%) | 220 (7.8%) |
| Education | | | | | | | |
| College/university degree or above | 31 (67.4%) | 1030 (59.0%) | 10 (66.7%) | 373 (60.4%) | 190 (56.5%) | 28 (60.9%) | 1662 (59.2%) |
| No college/university degree or above | 14 (30.4%) | 618 (35.4%) | 4 (26.7%) | 224 (36.2%) | 130 (38.7%) | 16 (34.8%) | 1006 (35.9%) |
| Social Deprivation | | | | | | | |
| Above the Townsend deprivation index median | 27 (58.7%) | 999 (57.2%) | 8 (53.3%) | 360 (58.3%) | 197 (58.6%) | 27 (58.7%) | 1618 (57.7%) |
| Below the Townsend deprivation index median | 19 (41.3%) | 743 (42.6%) | 7 (46.7%) | 257 (41.6%) | 137 (40.8%) | 19 (41.3%) | 1182 (42.1%) |
| Lifestyle factors | | | | | | | |
| Smoking status | | | | | | | |
| Ever smoked | 31 (67.4%) | 1010 (57.9%) | 9 (60.0%) | 369 (59.7%) | 181 (53.9%) | 25 (54.3%) | 1625 (57.9%) |
| Never smoked | 15 (32.6%) | 735 (42.1%) | 6 (40.0%) | 249 (40.3%) | 155 (46.1%) | 21 (45.7%) | 1181 (42.1%) |
| Doing regular physical activity | | | | | | | |
| Yes | 35 (76.1%) | 1323 (75.8%) | 12 (80.0%) | 481 (77.8%) | 252 (75.0%) | 33 (71.7%) | 2136 (76.1%) |
| No | 11 (23.9%) | 401 (23.0%) | 3 (20.0%) | 131 (21.2%) | 81 (24.1%) | 12 (26.1%) | 639 (22.8%) |
| Healthy diet | | | | | | | |
| Yes | 27 (58.7%) | 887 (50.8%) | 6 (40.0%) | 315 (51.0%) | 159 (47.3%) | 28 (60.9%) | 1422 (50.7%) |
| No | 19 (41.3%) | 852 (48.8%) | 9 (60.0%) | 300 (48.5%) | 176 (52.4%) | 18 (39.1%) | 1374 (49.0%) |
| Alcohol consumption | | | | | | | |
| Excessive alcohol consumption | 11 (23.9%) | 373 (21.4%) | 4 (26.7%) | 130 (21.0%) | 63 (18.8%) | 7 (15.2%) | 588 (21.0%) |
| Moderate alcohol consumption | 30 (65.2%) | 979 (56.1%) | 9 (60.0%) | 347 (56.1%) | 185 (55.1%) | 33 (71.7%) | 1583 (56.4%) |
| Not current drinking | 5 (10.9%) | 387 (22.2%) | 2 (13.3%) | 141 (22.8%) | 88 (26.2%) | 6 (13.0%) | 629 (22.4%) |
| Sleeping between 6 hours and 8 hours | | | | | | | |
| Yes | 18 (39.1%) | 752 (43.1%) | 5 (33.3%) | 266 (43.0%) | 153 (45.5%) | 22 (47.8%) | 1216 (43.3%) |
| No | 28 (60.9%) | 984 (56.4%) | 10 (66.7%) | 349 (56.5%) | 183 (54.5%) | 24 (52.2%) | 1578 (56.2%) |
| CVRFs factors | | | | | | | |
| Hypertension Status | | | | | | | |
| Yes | 10 (21.7%) | 359 (20.6%) | 3 (20.0%) | 112 (18.1%) | 57 (17.0%) | 10 (21.7%) | 551 (19.6%) |
| No | 36 (78.3%) | 1386 (79.4%) | 12 (80.0%) | 506 (81.9%) | 279 (83.0%) | 36 (78.3%) | 2255 (80.4%) |
| Diabetes | | | | | | | |
| Yes | 1 (2.2%) | 103 (5.9%) | 1 (6.7%) | 30 (4.9%) | 20 (6.0%) | 3 (6.5%) | 158 (5.6%) |
| No | 45 (97.8%) | 1642 (94.1%) | 14 (93.3%) | 588 (95.1%) | 316 (94.0%) | 43 (93.5%) | 2648 (94.4%) |
| BMI | | | | | | | |
| Mean(sd) | 27.1 (5.08) | 26.3 (4.20) | 25.1 (3.93) | 26.2 (4.14) | 26.7 (4.61) | 27.0 (4.95) | 26.4 (4.26) |
| Median(range) | 26.4 [18.6, 42.7] | 25.7 [17.4, 48.0] | 24.2 [20.2, 31.1] | 25.9 [16.5, 48.4] | 25.7 [18.1, 49.4] | 26.1 [19.7, 38.4] | 25.8 [16.5, 49.4] |

*: Percentiles do not add up to 1 is due to missingness.

*:All variables are measured at the baseline except the age at imaging, hypertension status, and diabetes.

**eTable 6. Summary of the numbers of associations.** Associations in the main findings are identified between brain functional connectivity (FC) measures and *APOE4* and ten modifiable health-related risk factors (MHRFs), as well as their interactions with age and sex, by the British population across three network atlases (Ji-12, Yeo-7, and Yeo-17). Findings validated from non-British populations are listed in parentheses if they have the same directions as main findings. Entries that are 100% validated by the non-British population are highlighted in yellow. The explanations of FC measures are provided in **eTable 3**. P: Positive (association); N: negative; Ex.Alcoh: excessive alcohol consumption; Mod.Alcoh: moderate alcohol consumption; SoDep: the index of social deprivation; Edu: education.

| FC Measures | APOE4 | Hypertension | BMI | Smoking | Alcohol | Physical Activity | Sleep | Education | Age & APOE Interaction | Other (Sex) Interaction |
|---|---|---|---|---|---|---|---|---|---|---|
| Ji12-NFC | N: 6 (5) | N: 12 (12) | P: 33 (27)<br>N: 7 (5) | P: 9 (7)<br>N: 6 (5) | **Ex. Alcoh**<br>N: 4 (0) | P: 1 (1)<br>N: 1 (1) | P: 1 (1)<br>N: 1 (1) | P: 13 (11)<br>N: 4 (4) | **Age*Hypertension**<br>P: 1 (1)<br>**Age*Sex**<br>P: 3 (3); N: 1 (0)<br>**APOE4*Sex**<br>N: 1 (1) | **Sex*Smoking**<br>N: 2 (1)<br>**Sex*Diabetes**<br>P: 1 (1)<br>**Sex*BMI**<br>P: 6 (4); N: 2 (2)<br>**Sex*Edu**<br>N: 2 (1) |
| Ji12-NES | N: 7 (5) | N: 33 (33) | P: 6 (4)<br>N: 20 (20) | P: 1 (1)<br>N: 9 (9) | **Ex. Alcoh**<br>N: 5 (5) | P: 1 (1) | P: 1 (1) | P: 16 (16)<br>N: 1 (0) | **Age*BMI**<br>N: 1 (1)<br>**Age*Sex**<br>P: 18 (18) | **Sex*Smoking**<br>N: 2 (0)<br>**Sex*BMI**<br>P: 1 (0); N: 4 (2)<br>**Sex*Edu**<br>P: 1 (1)<br>**Sex*Mod.Alcoh**<br>P: 2 (2)<br>**Sex*Ex.Alcoh**<br>N: 2 (1) |
| Yeo7-NFC | N: 7 (5) | N: 2 (2) | P: 15 (14)<br>N: 1 (0) | P: 6 (5)<br>N: 3 (2) | **Mod. Alcoh**<br>P: 1 (0)<br>**Ex. Alcoh**<br>P: 1 (1)<br>N: 3 (0) | P: 2 (2) | P: 3 (3) | P: 8 (8) | **Age*Hypertension**<br>P: 1 (1)<br>**Age*Sex**<br>P: 3 (3); N: 2 (1)<br>**APOE4*Sex**<br>N: 1 (0)<br>**APOE4*Sex*Diabetes**<br>P: 1 (0)<br>**APOE4*Sex*BMI**<br>P: 1 (0) | **Sex*BMI**<br>P: 11 (11)<br>**Sex*Ex.Alcoh**<br>N: 1 (1) |
| Yeo7-NES | N: 4 (4) | N: 7 (7) | P: 10 (9)<br>N: 6 (6) | P: 1 (0)<br>N: 5 (4) | **Ex. Alcoh**<br>N: 4 (4) | P: 2 (2) | P: 2 (2) | P: 6 (6) | **Age*BMI**<br>N: 1 (1)<br>**Age*Sex**<br>P: 7 (7); N: 1 (0)<br>**Age*APOE4*Edu**<br>N: 1 (1)<br>**APOE4*Sex*Diabetes**<br>P: 1 (0) | **Sex*BMI**<br>P: 2 (2)<br>**Sex*Ex.Alcoh**<br>N: 1 (1)<br>**Sex*SoDep**<br>N : 1 (0) |
| Yeo17-NFC | N: 15 (8) | P: 2 (2)<br>N: 8 (7) | P: 66 (62)<br>N: 2 (1) | P: 9 (7)<br>N: 6 (6) | **Ex. Alcoh**<br>P: 3 (2)<br>N: 9 (3) | P: 4 (4) | P: 2 (1) | P: 27 (25)<br>N: 3 (2) | **Age*BMI**<br>N: 1 (1)<br>**Age*Sex**<br>P: 8 (6); N: 5 (3)<br>**Age*APOE4*Edu**<br>N: 1 (1)<br>**Age*APOE4*Sleep**<br>P: 1 (1)<br>**APOE4*Sex**<br>N: 1 (0)<br>**APOE4*Sex*BMI**<br>P: 1 (0)<br>**APOE4*Sex*Diabetes**<br>P: 2 (0) | **Sex*BMI**<br>P: 21 (20);<br>N: 1 (1) |
| Yeo17-NES | N: 3 (3) | N: 21 (21) | P: 32 (28)<br>N: 18 (17) | P: 1 (1)<br>N: 7 (7) | **Ex. Alcoh**<br>N: 3 (3) | P: 3 (3) | P: 1 (1) | P: 14 (14)<br>N: 3 (2) | **Age*BMI**<br>N: 1 (1)<br>**Age*Diet**<br>N: 1 (0)<br>**Age*Sex**<br>P: 11 (10); N: 2 (0)<br>**Age*APOE4*Edu**<br>N: 1 (1)<br>**APOE4*Sex*Diabetes**<br>P: 2 (0) | **Sex*BMI**<br>P: 6 (5); N: 1 (1)<br>**Sex*Edu**<br>P: 1 (1)<br>**Sex*Mod.Alcoh**<br>P: 1 (0)<br>**Sex*Ex.Alcoh**<br>N: 3 (1) |

#### Supplement 3. Supplementary figures

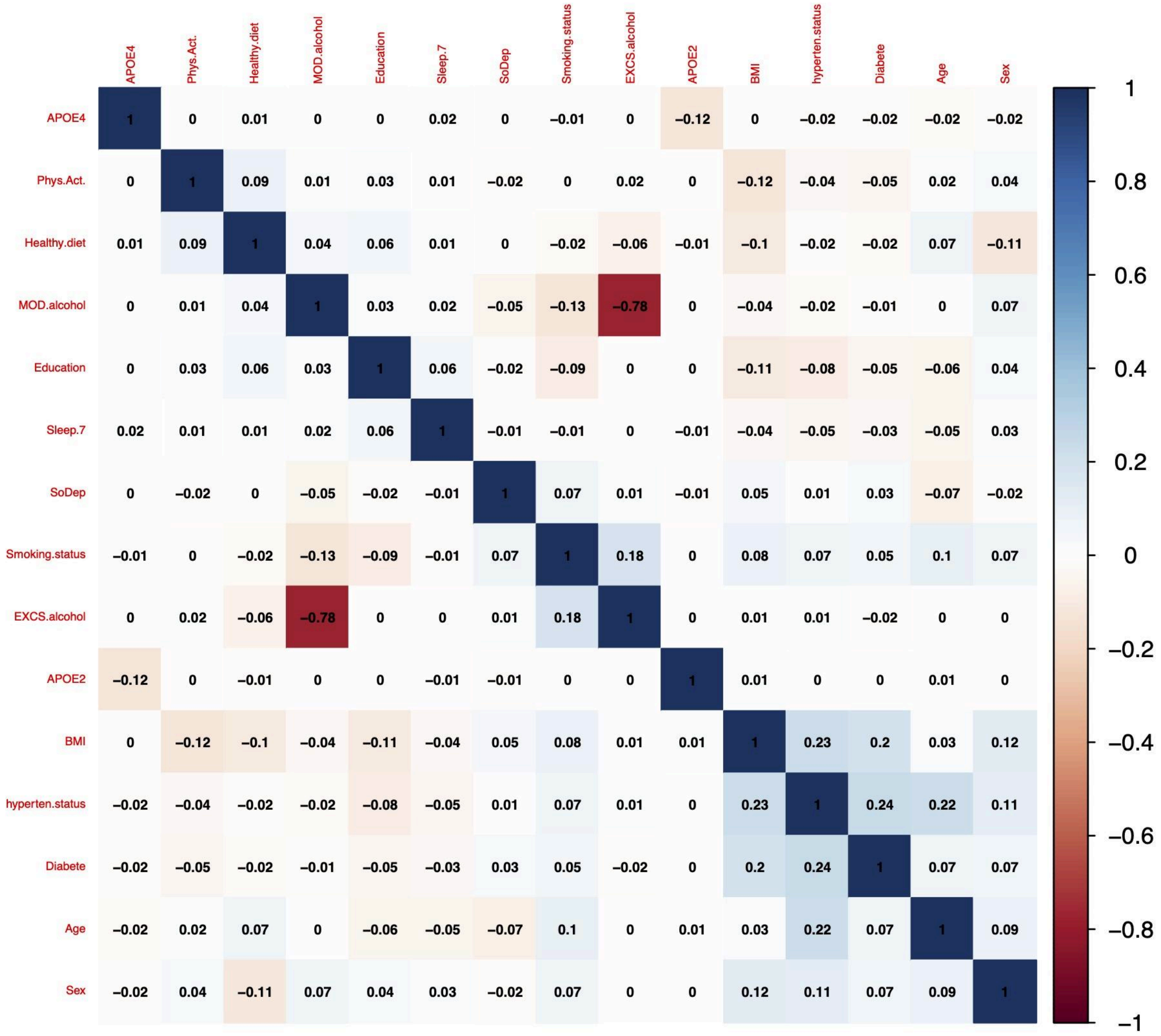


**eFigure 1 Correlation plot of age, sex, *APOE* and ten modifiable health-related risk factors (MHRFs)**. SoDep, Social deprivation Index; NotSmoking, not-current-smoker; Phys.Act, physical activity; Healthy.diet, healthy diet; Sleep.7, healthy sleep; MOD.alcohol, moderate alcohol; EXCS.alcohol, excessive alcohol.

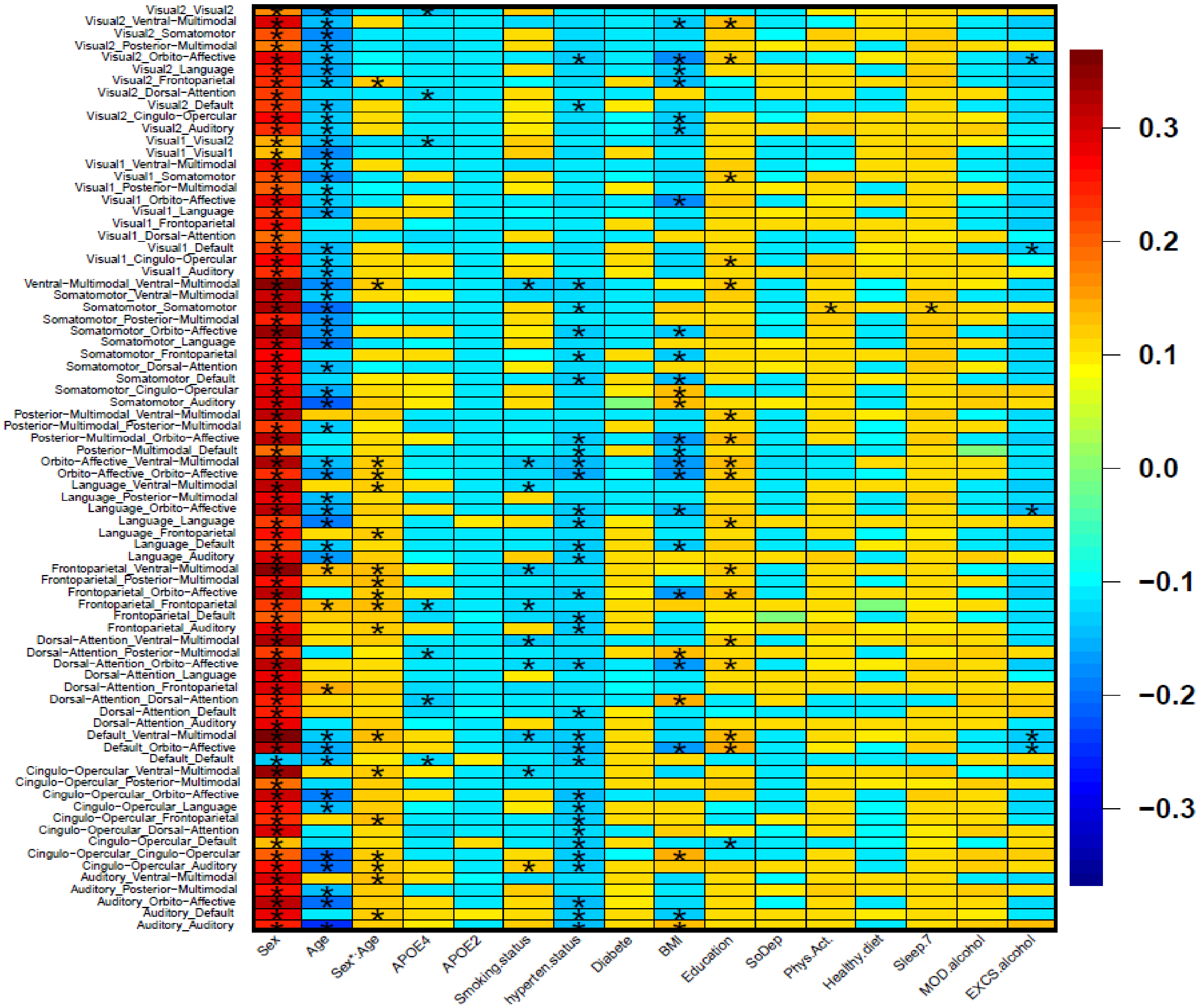


**eFigure 2. Heatmap of all the identified main effects of age, sex, *APOE* gene, and ten modifiable health-related risk factors (MHRFs) on the network edge strength (NES) measures based on the Ji-12 network atlas**. Results that passed the Bonferroni-corrected significance level of *6.4E-4* will be shown by (*). The effect sizes (standardized coefficients) for each association are illustrated using a color gradient from blue to red.

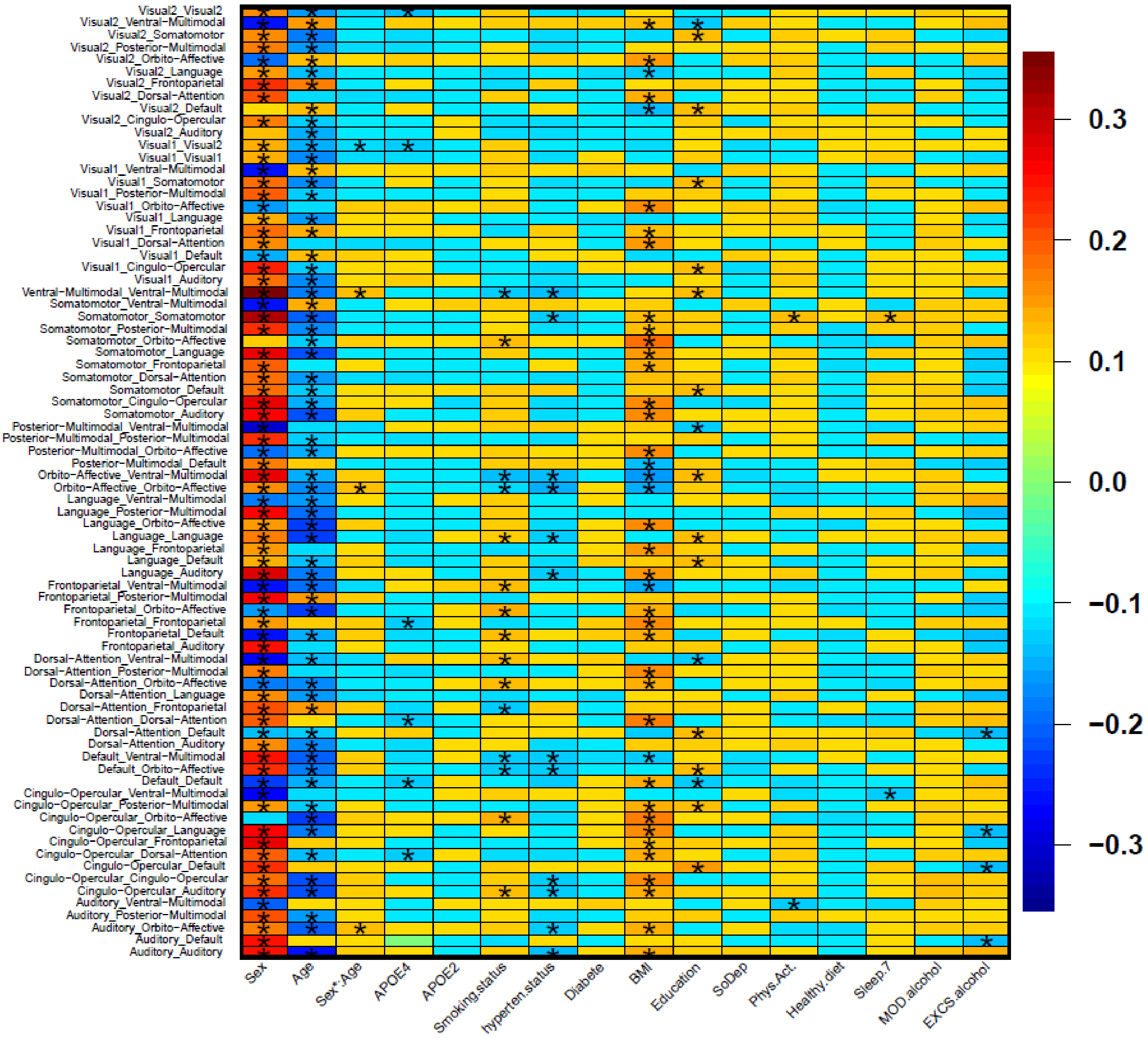


**eFigure 3. Heatmap of all the identified main effects of age, sex, *APOE* gene, and ten modifiable health-related risk factors (MHRFs) on the network functional connectivity (NFC) measures, based on the Ji-12 network atlas**. Results that passed the Bonferroni-corrected significance level of *6.4E-4* will be shown by (*). The effect sizes (standardized coefficients) for each association are illustrated using a color gradient from blue to red.

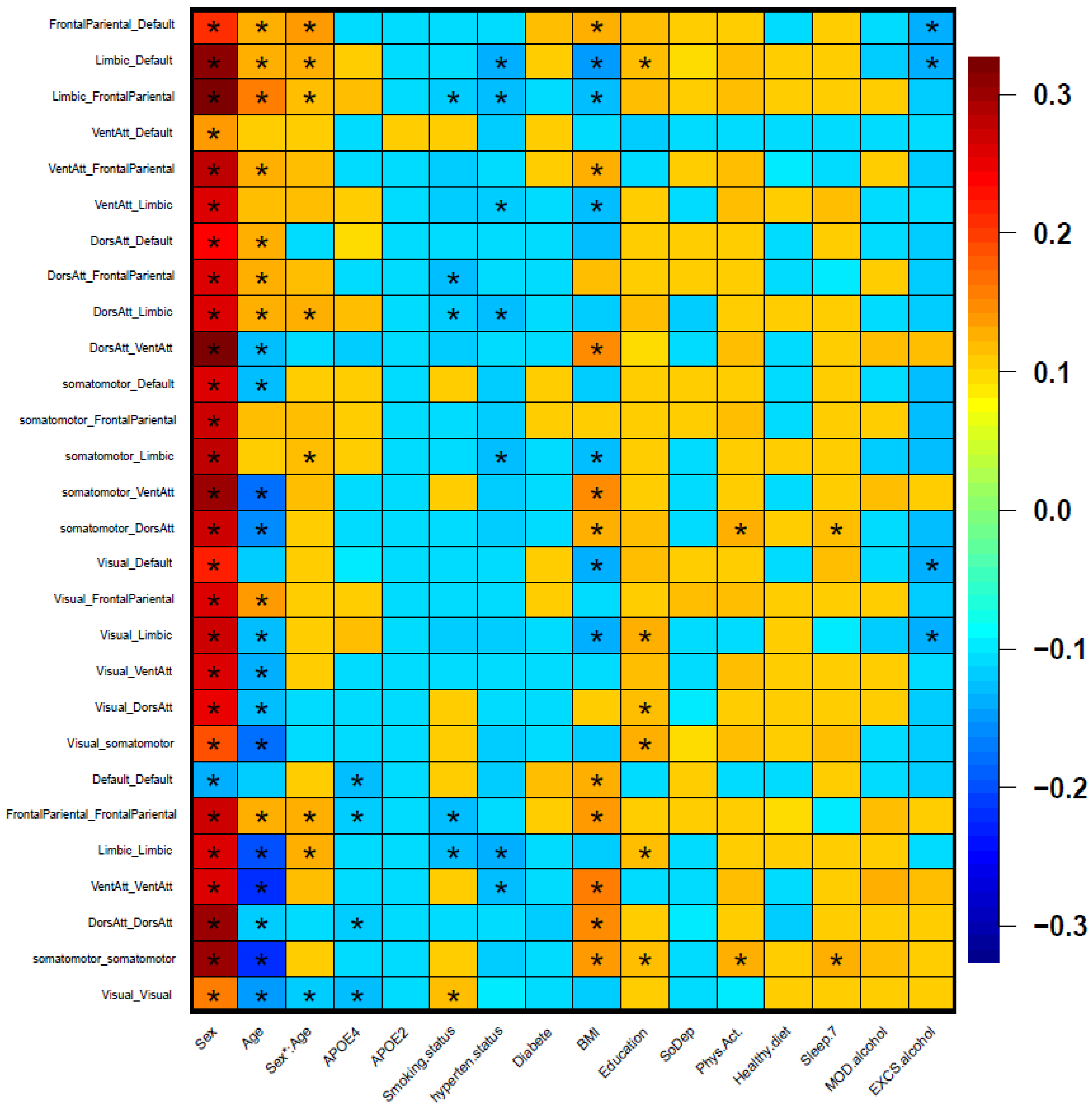


**eFigure 4. Heatmap of all the identified main effects of age, sex, *APOE* gene, and ten modifiable health-related risk factors (MHRFs) on the network edge strength (NES) measures, based on the Yeo-7 network atlas**. Results that passed the Bonferroni-corrected significance level of *1.8E-3* are shown by (*). The effect sizes (standardized coefficients) for each association are illustrated using a color gradient from blue to red.

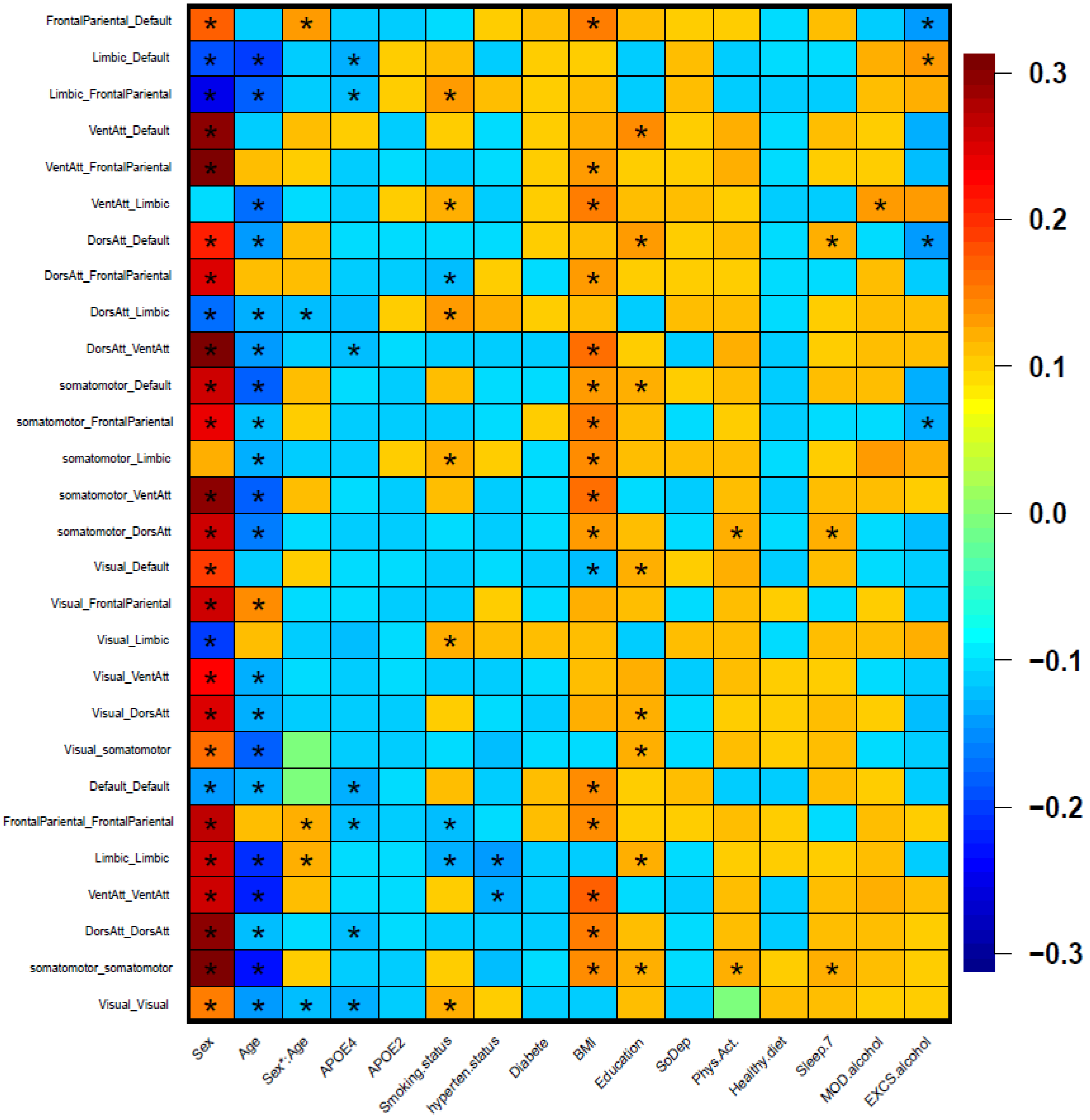


**eFigure 5. Heatmap of all the identified main effects of age, sex, *APOE* gene, and ten modifiable health-related risk factors (MHRFs) on the network functional connectivity (NFC) measures, based on the Yeo-7 network atlas**. Results that passed the Bonferroni-corrected significance level of *1.8E-3* are shown by (*). The effect sizes (standardized coefficients) for each association are illustrated using a color gradient from blue to red.

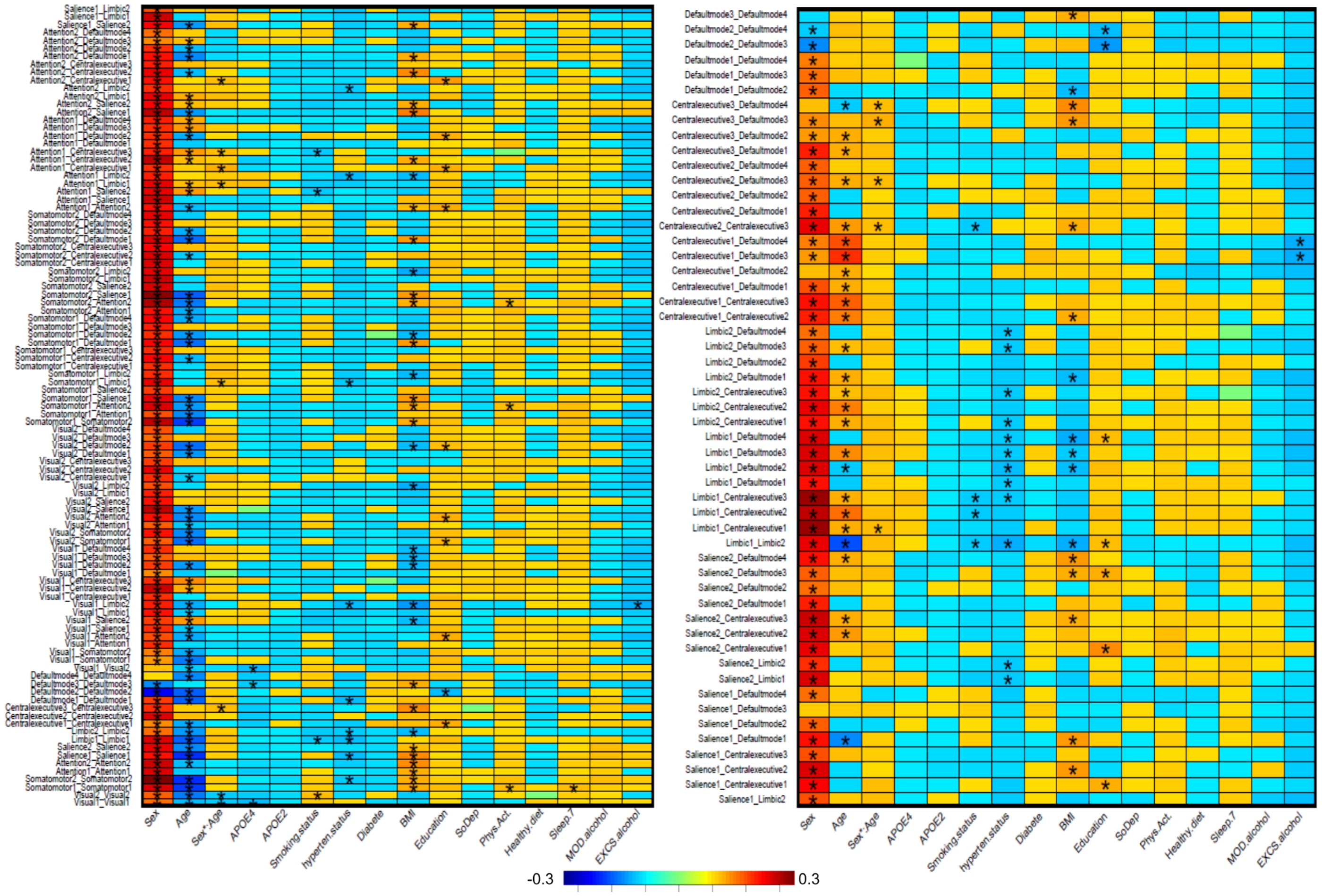


**eFigure 6. Heatmap of all the identified main effects of age, sex, *APOE* gene, and ten modifiable health-related risk factors (MHRFs) on the network edge strength (NES) measures, based on the Yeo-17 network atlas**. Results that passed the Bonferroni-corrected significance level of *3.3E-4* are shown by (*). The effect sizes (standardized coefficients) for each association are illustrated using a color gradient from blue to red.

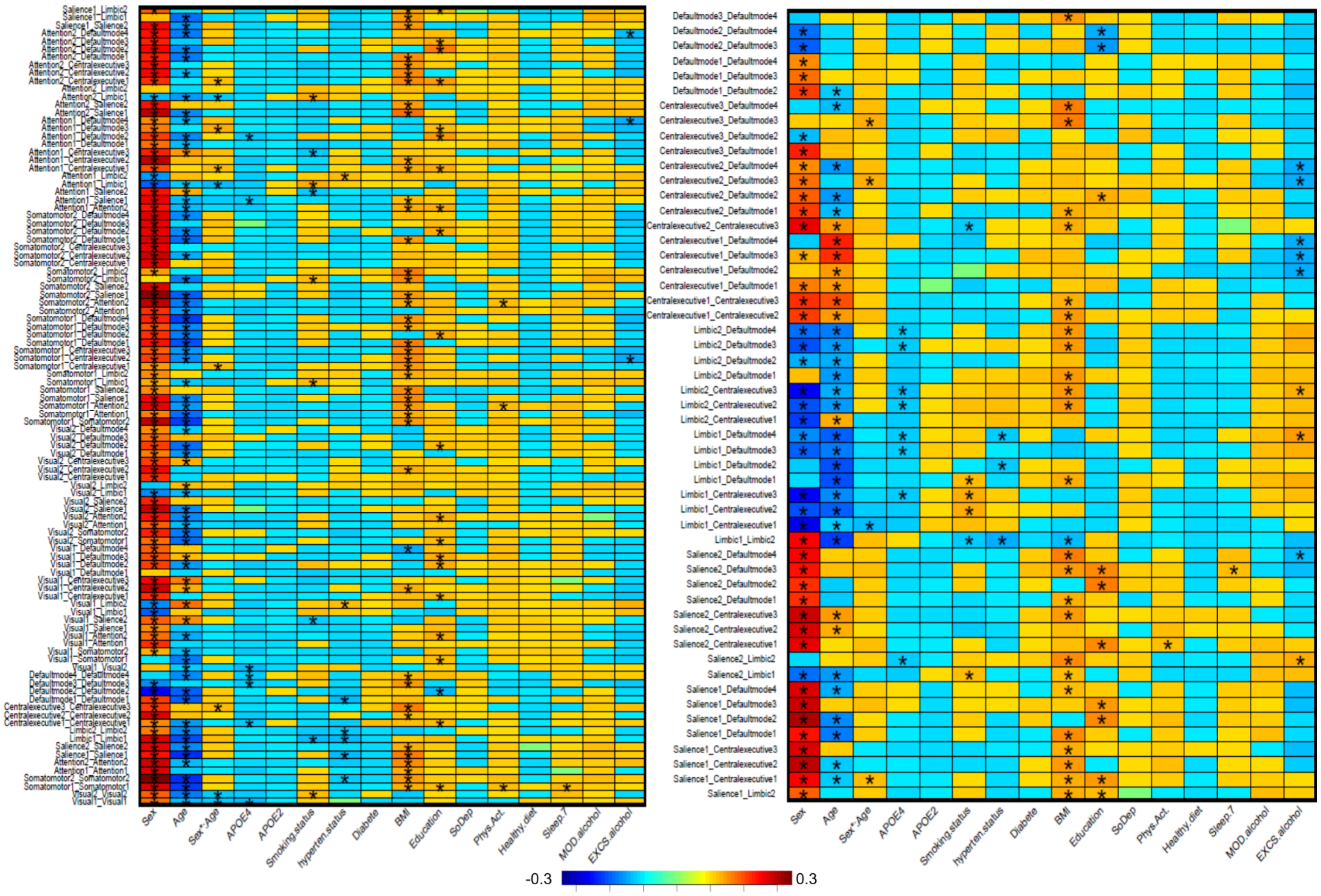


**eFigure 7. Heatmap of all the identified main effects of age, sex, *APOE* gene, and ten modifiable health-related risk factors (MHRFs) on the network functional connectivity (NFC) measures, based on the Yeo-17 network atlas**. Results that passed the Bonferroni-corrected significance level of *3.3E-4* are shown by (*). The effect sizes (standardized coefficients) for each association are illustrated using a color gradient from blue to red.

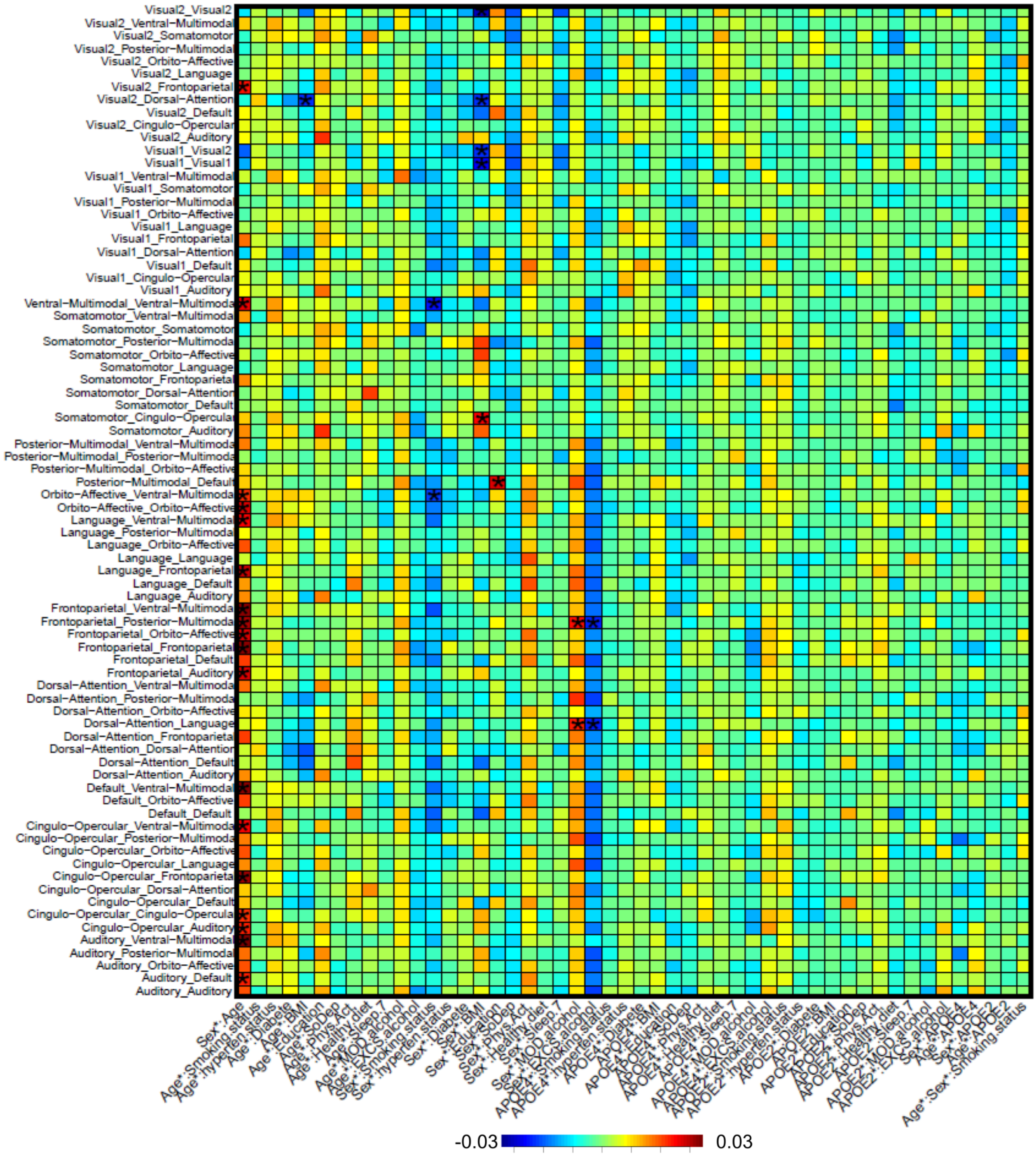

**eFigure 8. Heatmap of all the identified two-way interaction effects of age, sex, *APOE* gene, and ten modifiable health-related risk factors (MHRFs) on the network edge strength (NES) measures, based on the Ji-12 network atlas**. Results that passed the Bonferroni-corrected significance level of *6.4E-4* are shown by (*). The effect sizes (standardized coefficients) for each association are illustrated using a color gradient from blue to red.

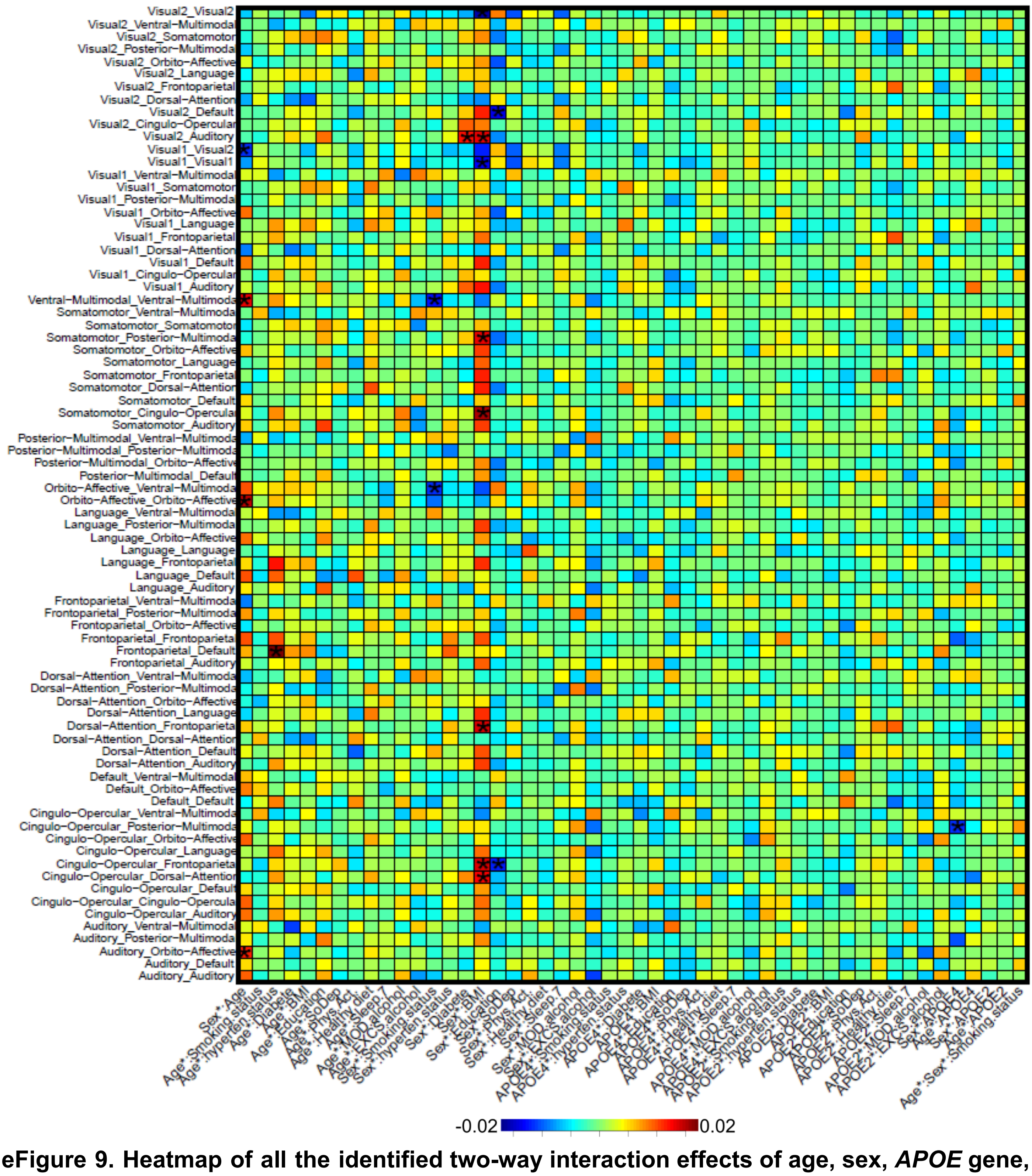

**eFigure 9. Heatmap of all the identified two-way interaction effects of age, sex, *APOE* gene, and ten modifiable health-related risk factors (MHRFs) on the network functional connectivity (NFC) measures, based on the Ji-12 network atlas**. Results that passed the Bonferroni-corrected significance level of *6.4E-4* are shown by (*). The effect sizes (standardized coefficients) for each association are illustrated using a color gradient from blue to red.

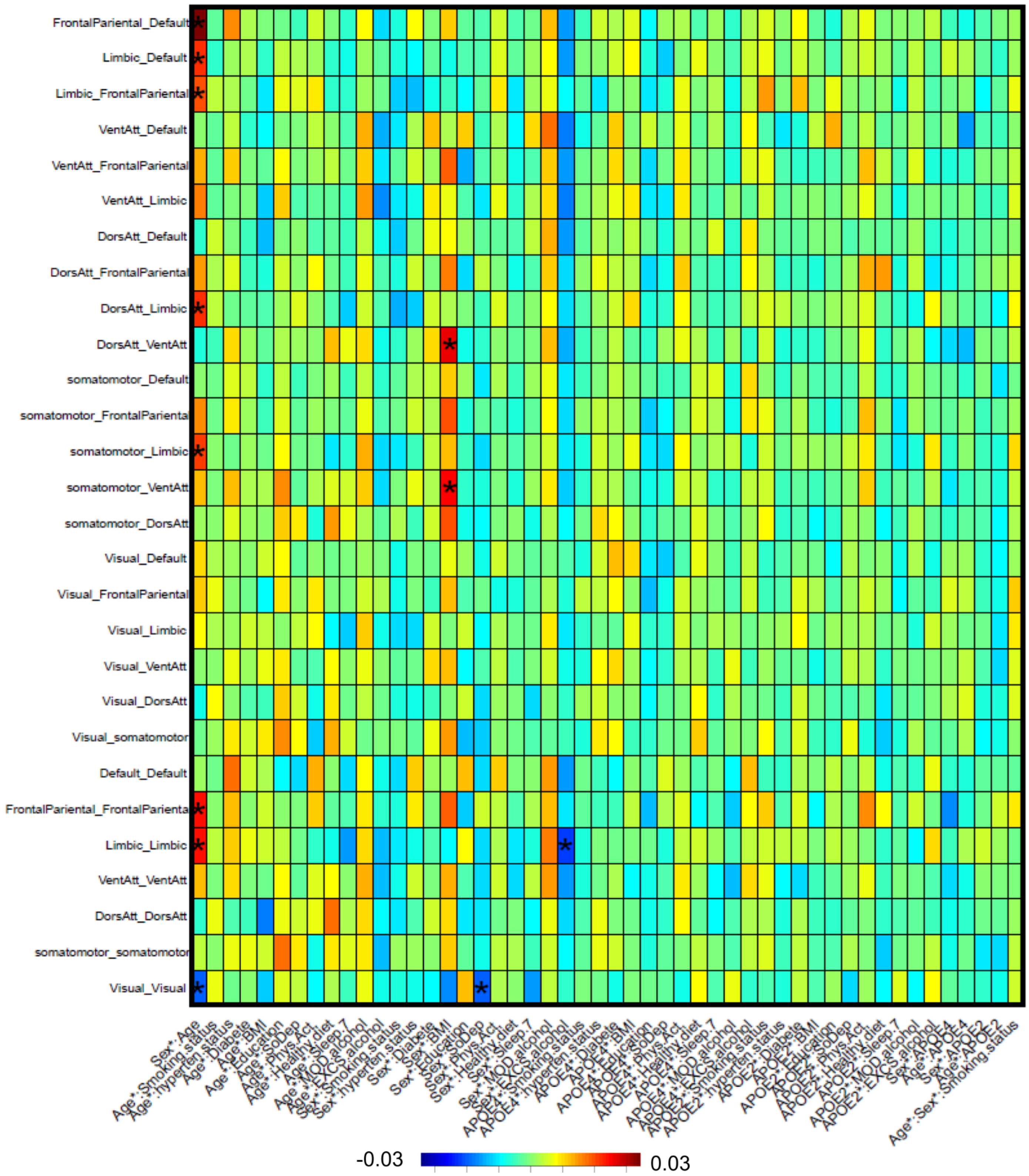


**eFigure 10. Heatmap of all the identified two-way interaction effects of age, sex, *APOE* gene, and ten modifiable health-related risk factors (MHRFs) on the network edge strength (NES) measures, based on the Yeo-7 network atlas**. Results that passed the Bonferroni-corrected significance level of *1.8E-3* are shown by (*). The effect sizes (standardized coefficients) for each association are illustrated using a color gradient from blue to red.

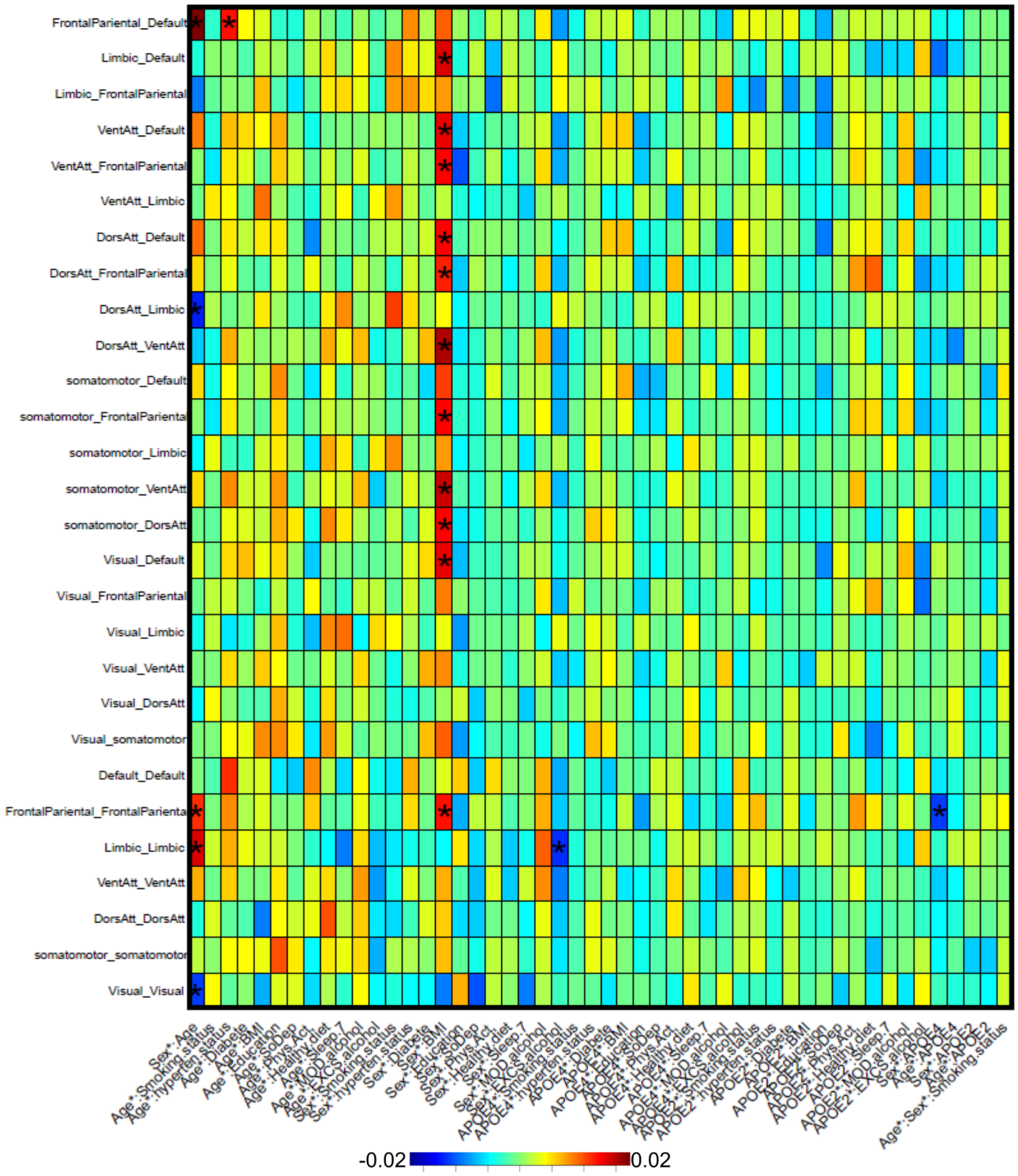


**eFigure 11. Heatmap of all the identified two-way interaction effects of age, sex, *APOE* gene, and ten modifiable health-related risk factors (MHRFs) on the network functional connectivity (NFC) measures, based on the Yeo-7 network atlas**. Results that passed the Bonferroni-corrected significance level of *1.8E-3* are shown by (*). The effect sizes (standardized coefficients) for each association are illustrated using a color gradient from blue to red.

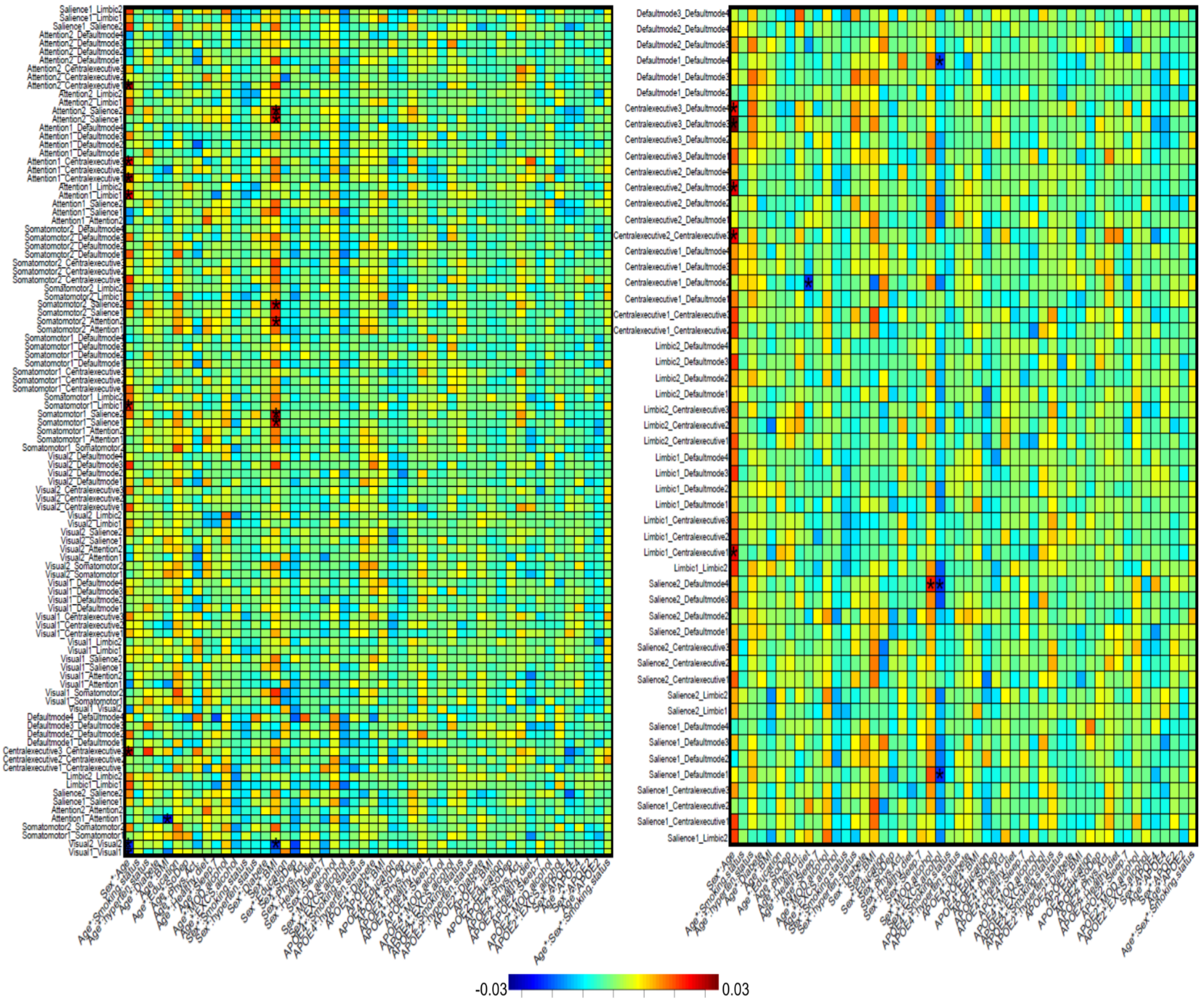


**eFigure 12. Heatmap of all the identified two-way interaction effects of age, sex, *APOE* gene, and ten modifiable health-related risk factors (MHRFs) on the network edge strength (NES) measures, based on the Yeo-17 network atlas**. Results that passed the Bonferroni-corrected significance level of *3.3E-4* are shown by (*). The effect sizes (standardized coefficients) for each association are illustrated using a color gradient from blue to red.

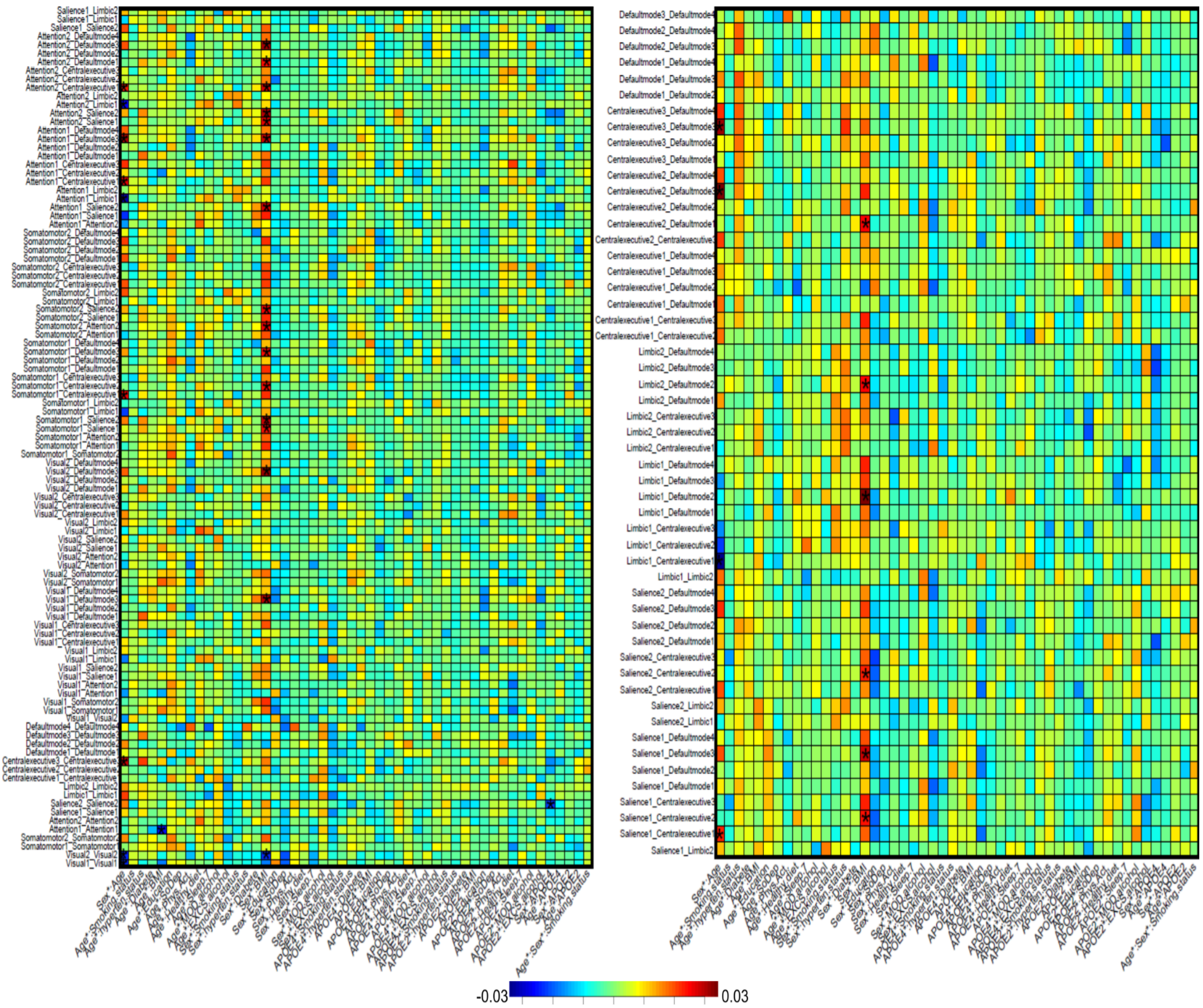


**eFigure 13. Heatmap of all the identified two-way interaction effects of age, sex, *APOE* gene, and ten modifiable health-related risk factors (MHRFs) on the network functional connectivity (NFC) measures, based on the Yeo-17 network atlas**. Results that passed the Bonferroni-corrected significance level of *3.3E-4* are shown by (*). The effect sizes (standardized coefficients) for each association are illustrated using a color gradient from blue to red.

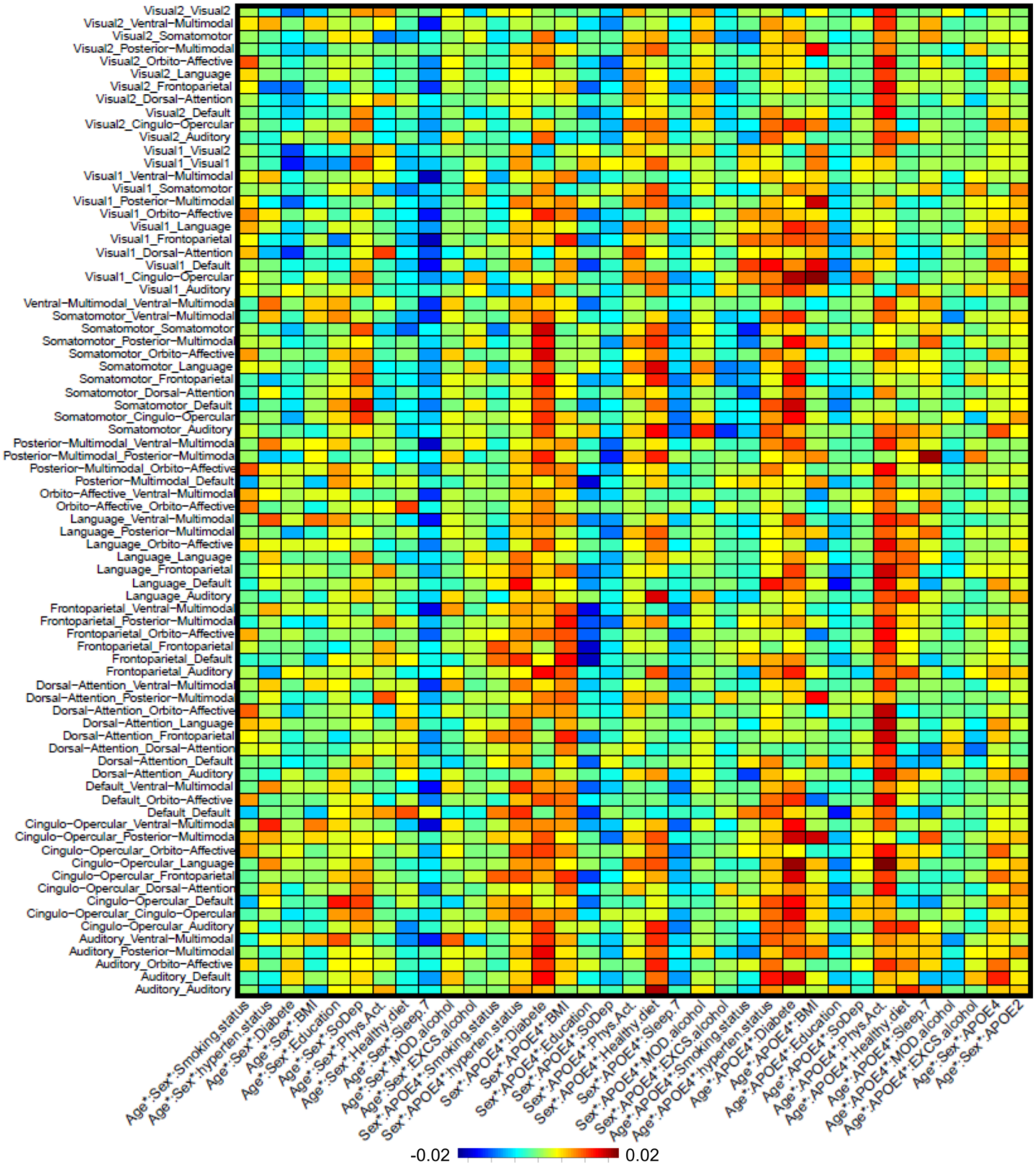


**eFigure 14. Heatmap of all the identified three-way interaction effects of age, sex, *APOE* gene, and ten modifiable health-related risk factors (MHRFs) on the network edge strength (NES) measures, based on the Ji-12 network atlas**. Results that passed the Bonferroni-corrected significance level of *6.4E-4* are shown by (*). The effect sizes (standardized coefficients) for each association are illustrated using a color gradient from blue to red.

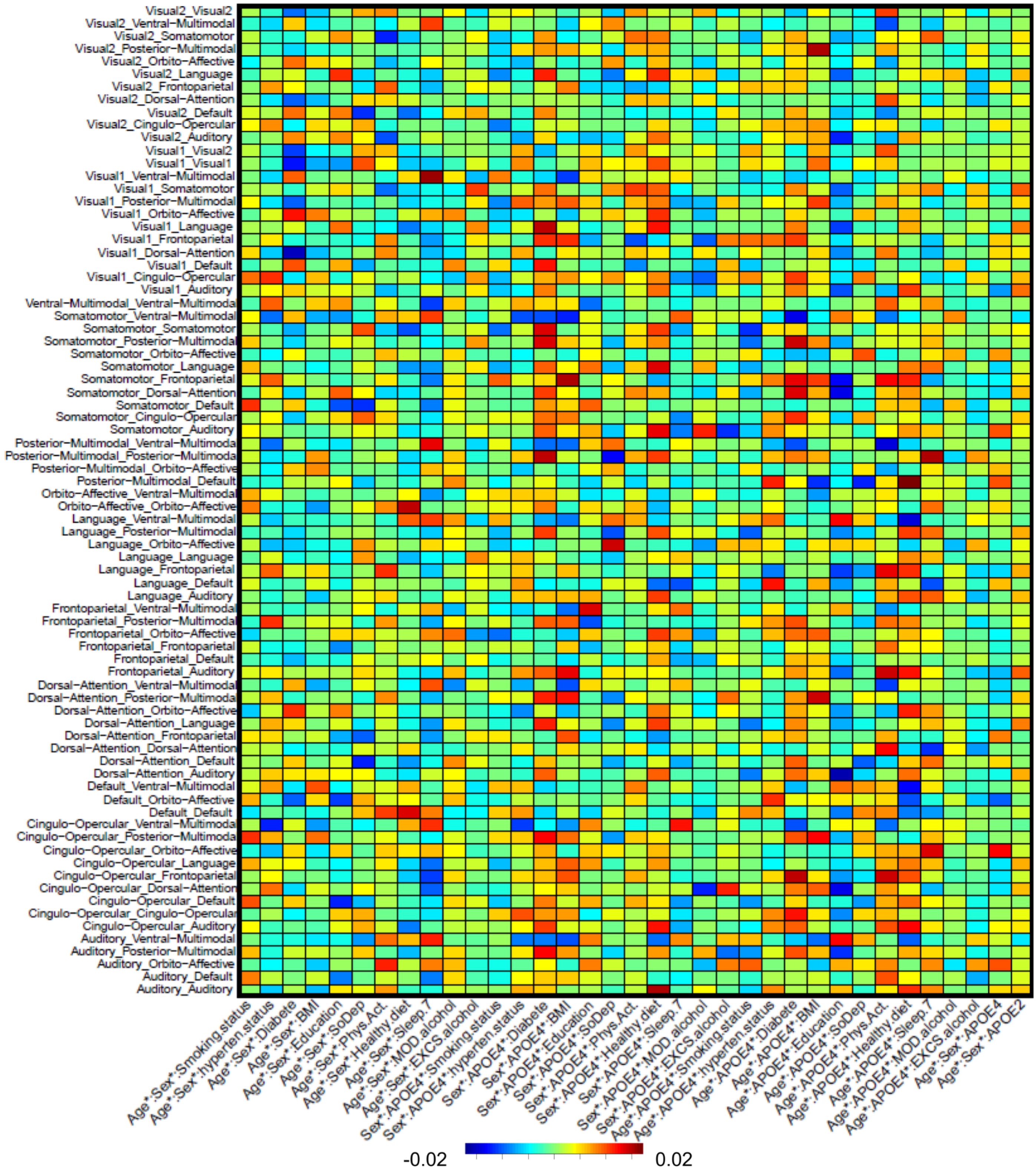


**eFigure 15. Heatmap of all the identified three-way interaction effects of age, sex, *APOE* gene, and ten modifiable health-related risk factors (MHRFs) on the network functional connectivity (NFC) measures, based on the Ji-12 network atlas**. Results that passed the Bonferroni-corrected significance level of *6.4E-4* are shown by (*). The effect sizes (standardized coefficients) for each association are illustrated using a color gradient from blue to red.

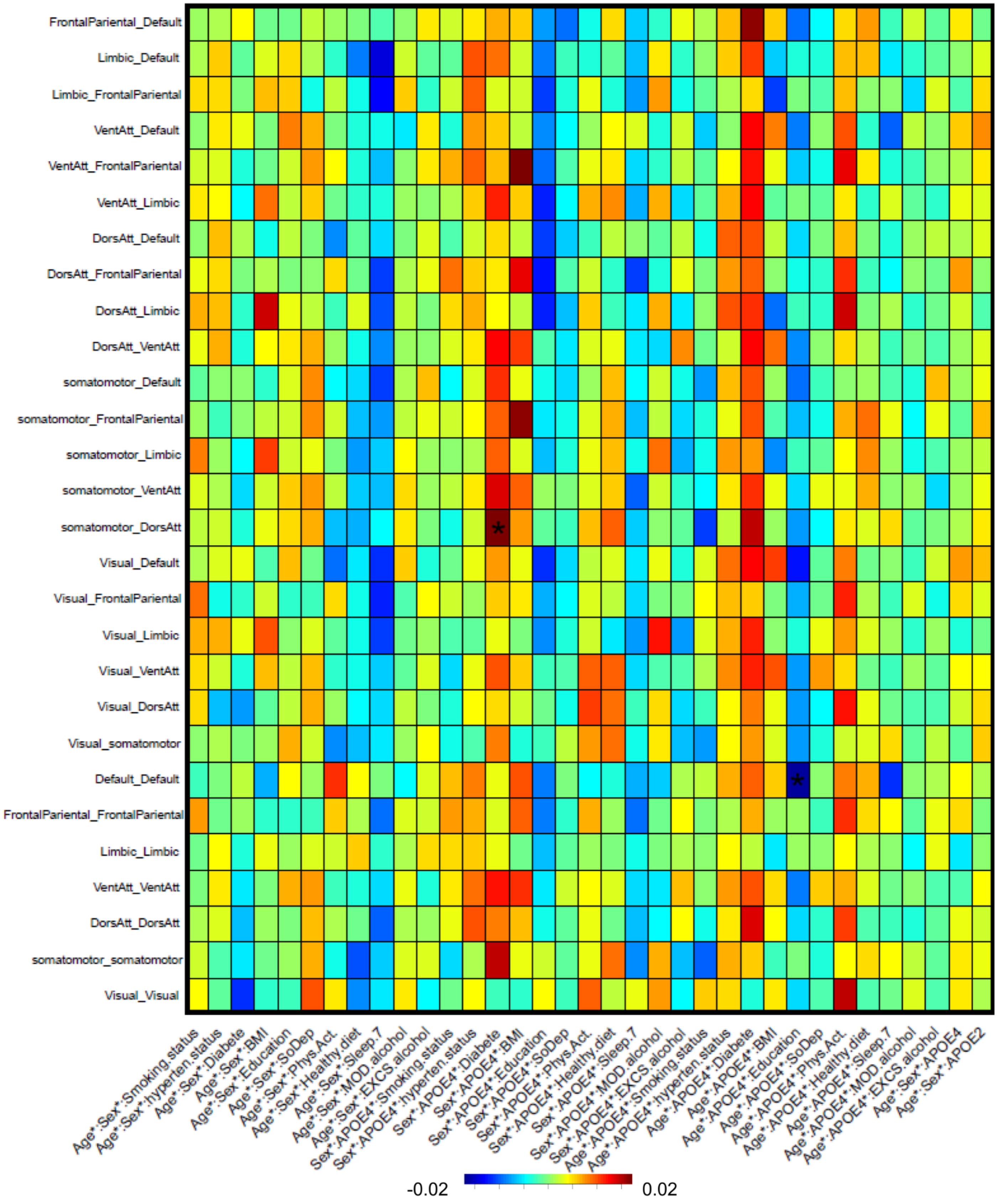


**eFigure 16. Heatmap of all the identified three-way interaction effects of age, sex, *APOE* gene, and ten modifiable health-related risk factors (MHRFs) on the network edge strength (NES) measures, based on the Yeo-7 network atlas**. Results that passed the Bonferroni-corrected significance level of *1.8E-3* are shown by (*). The effect sizes (standardized coefficients) for each association are illustrated using a color gradient from blue to red.

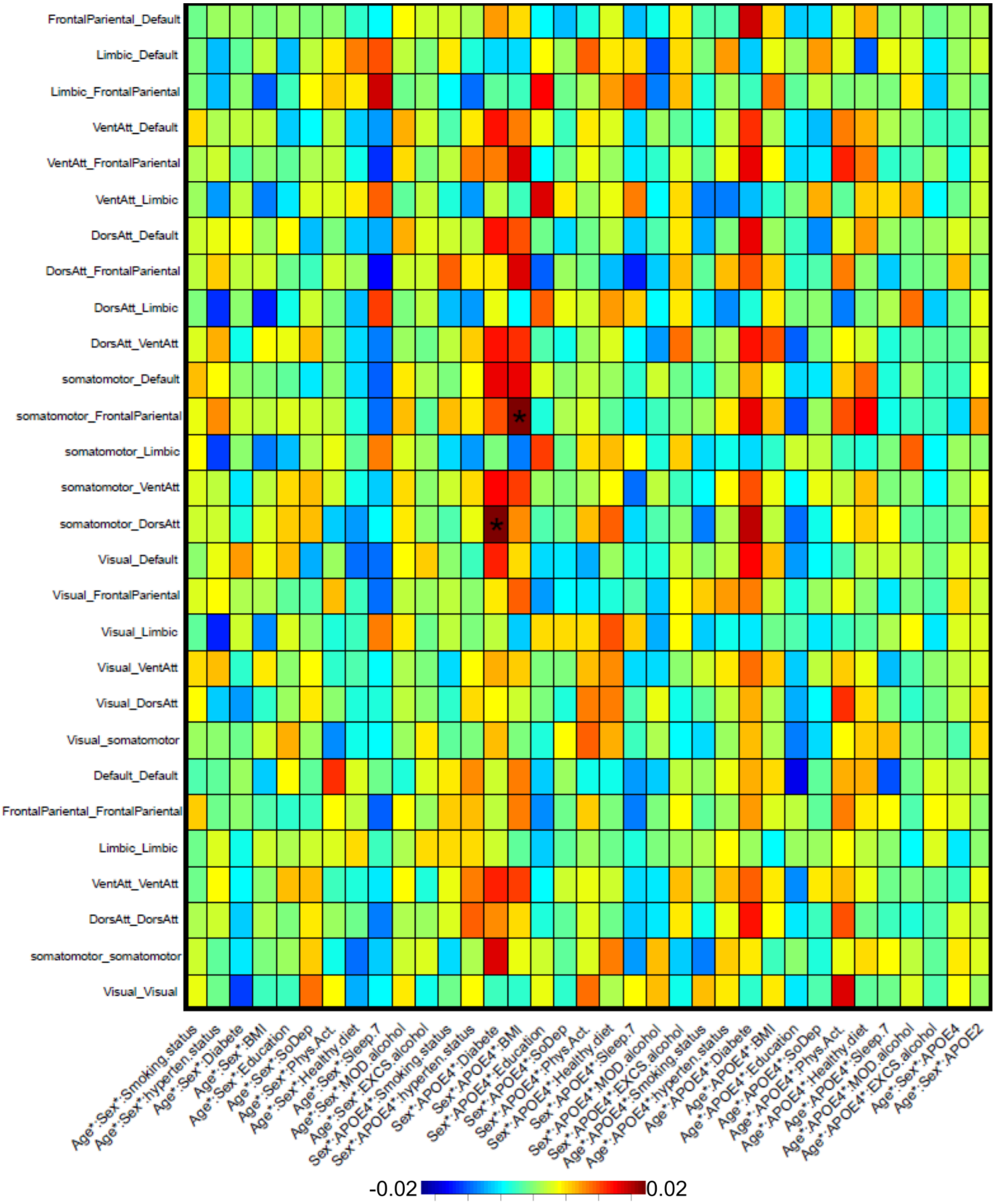


**eFigure 17. Heatmap of all the identified three-way interaction effects of age, sex, *APOE* gene, and ten modifiable health-related risk factors (MHRFs) on the network functional connectivity (NFC) measures, based on the Yeo-7 network atlas**. Results that passed the Bonferroni-corrected significance level of *1.8E-3* are shown by (*). The effect sizes (standardized coefficients) for each association are illustrated using a color gradient from blue to red.

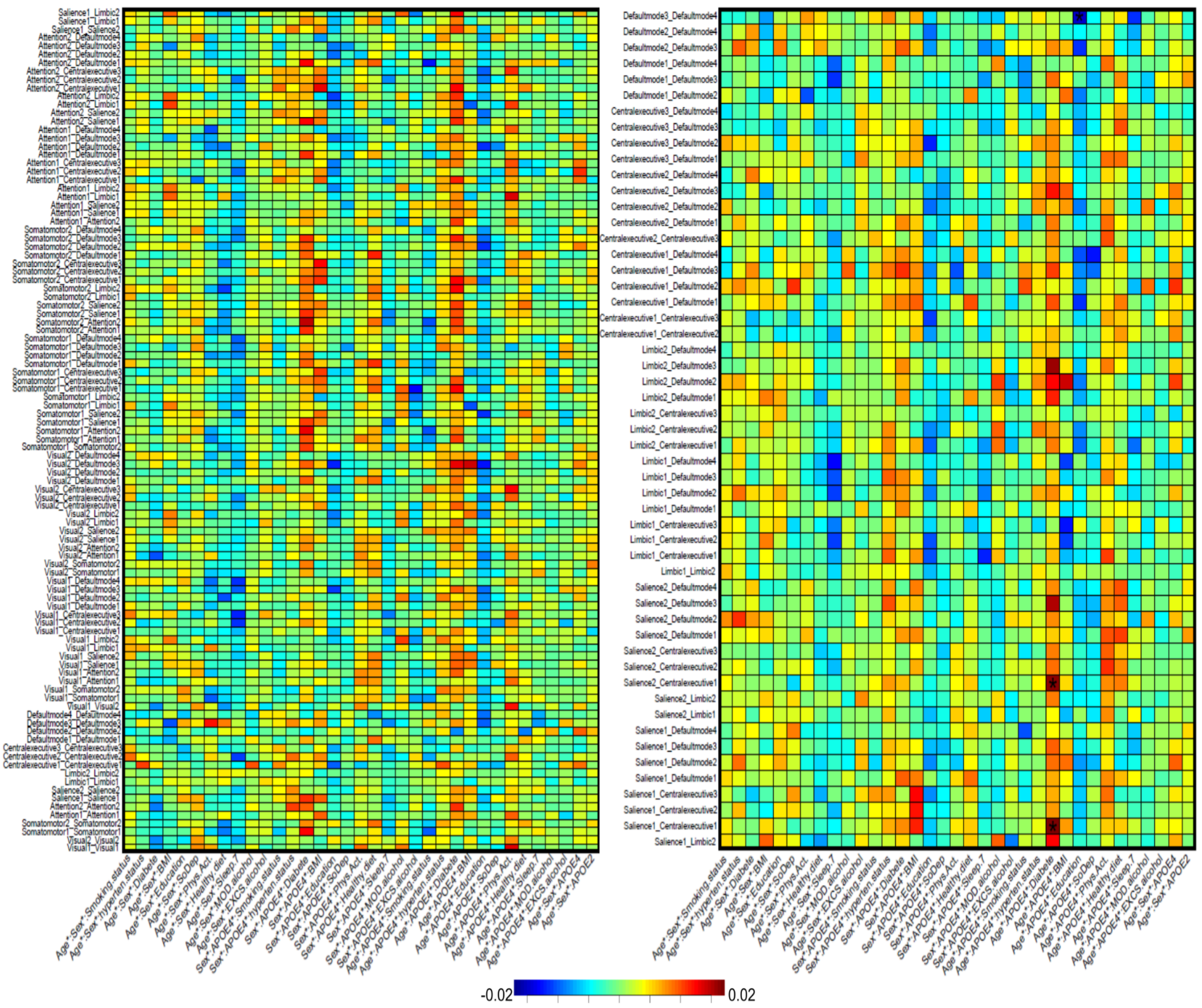


**eFigure 18. Heatmap of all the identified three-way interaction effects of age, sex, *APOE* gene, and ten modifiable health-related risk factors (MHRFs) on the network edge strength (NES) measures, based on the Yeo-17 network atlas**. Results that passed the Bonferroni-corrected significance level of *3.3E-4* are shown by (*). The effect sizes (standardized coefficients) for each association are illustrated using a color gradient from blue to red.

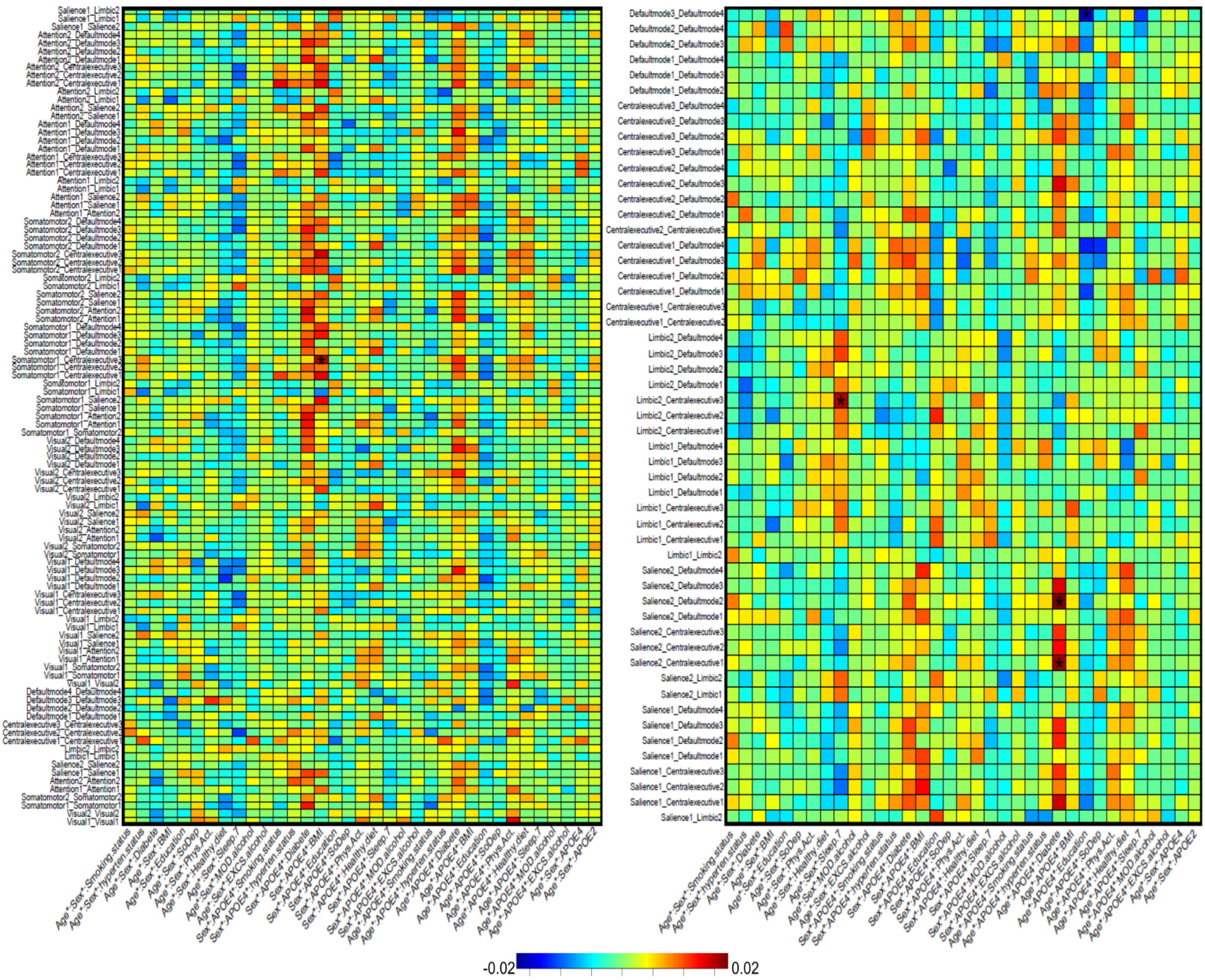


**eFigure 19. Heatmap of all the identified three-way interaction effects of age, sex, *APOE* gene, and ten modifiable health-related risk factors (MHRFs) on the network functional connectivity (NFC) measures, based on the Yeo-17 network atlas.** Results that passed the Bonferroni-corrected significance level of *3.3E-4* are shown by (*). The effect sizes (standardized coefficients) for each association are illustrated using a color gradient from blue to red.

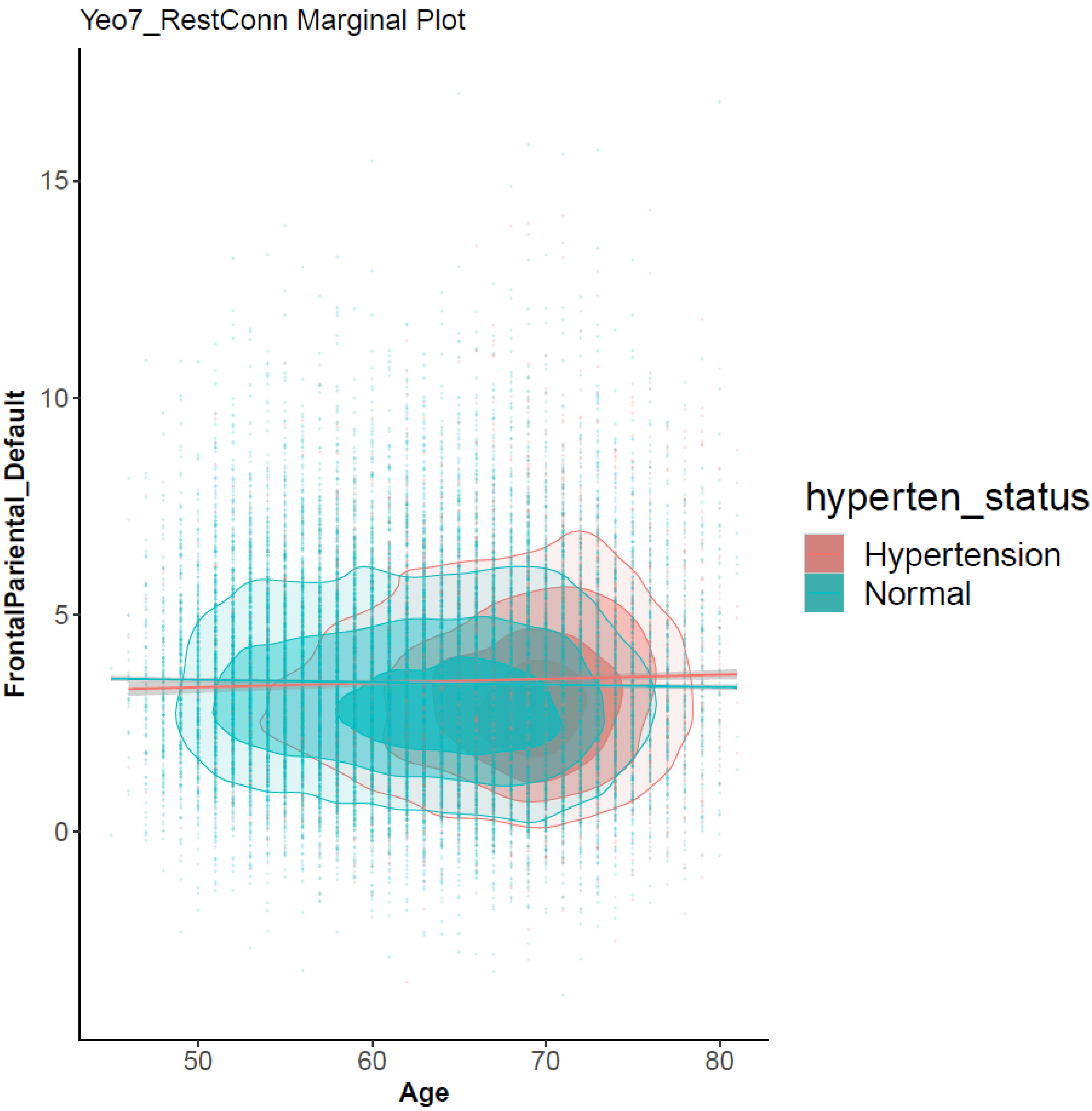


**eFigure 20. The scatterplots of FPN-DMN between-network functional connectivity (NFC) against age grouped by hypertension.** Results are based on the Yeo-7 network atlas. Contour plots are used to show the density of two-dimensional distributions; regression lines and 95% confidence intervals are also drawn. FPN, frontoparietal network; DMN, default mode network.

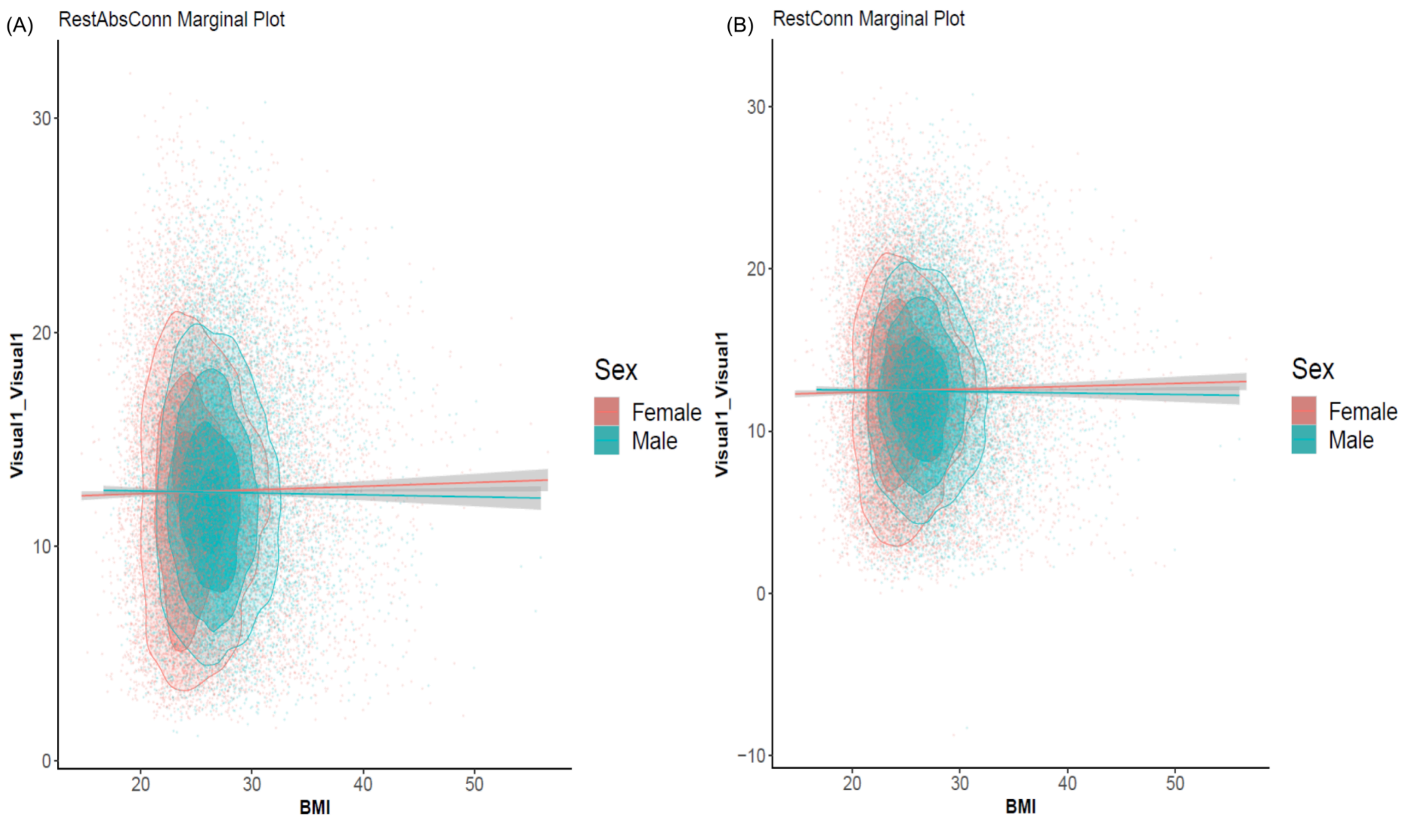


**eFigure 21. The scatterplots of (A) within-Vi1-network edge strength (NES) and (B) network-level functional connectivity (NFC) against BMI grouped by sex. Results are based on the Ji-12 network atlas.** Contour plots are used to show the density of two-dimensional distributions; regression lines and 95% confidence intervals are also drawn. Vi1, visual 1 network.

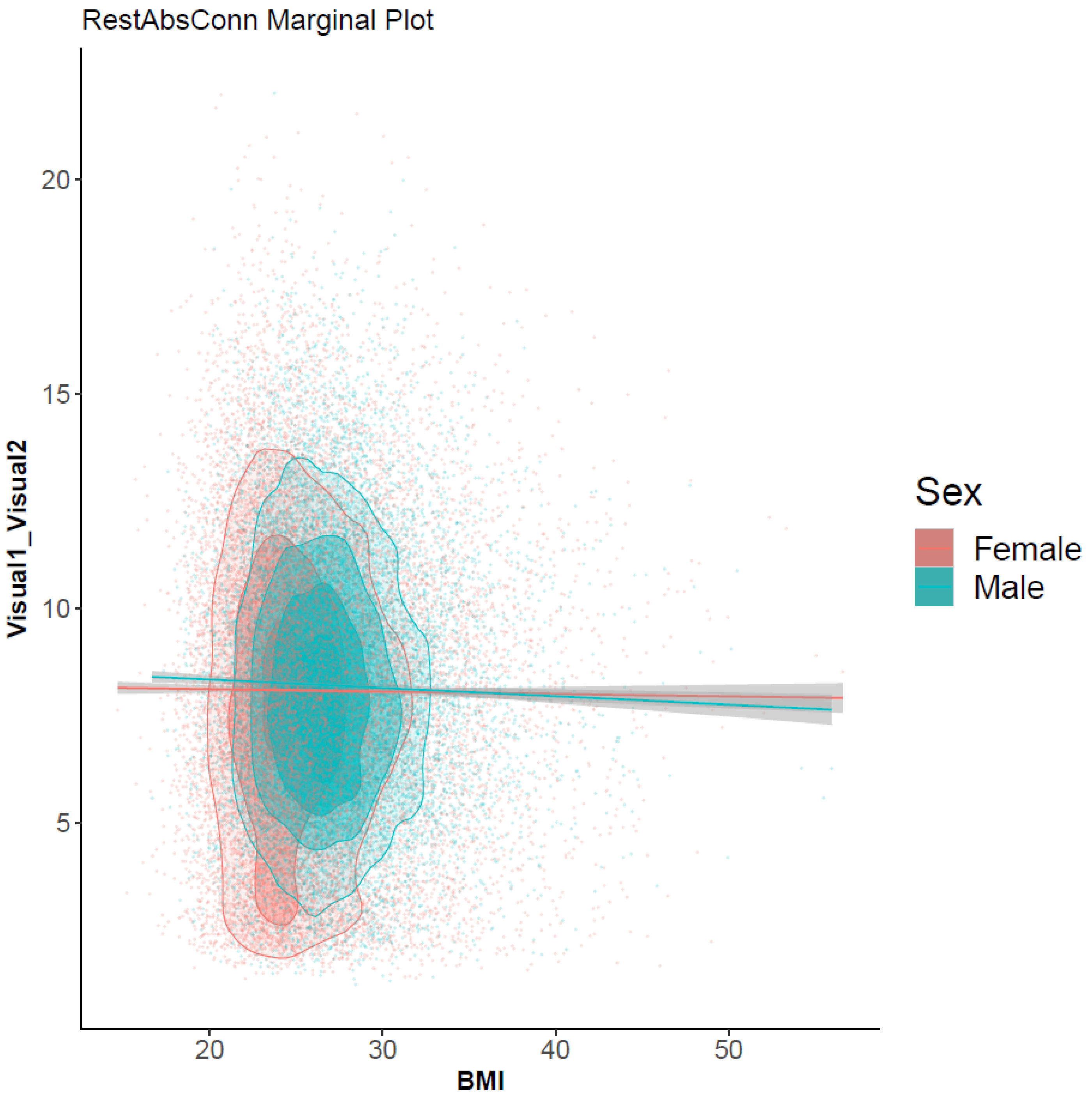


**eFigure 22. The scatterplots of Vi1-Vi2 between-network edge strength (NES) against BMI grouped by sex.** Results are based on the Ji-12 network atlas. Contour plots are used to show the density of two-dimensional distributions; regression lines and 95% confidence intervals were also drawn. Vi1, visual 1 network; Vi2, visual 2 network.

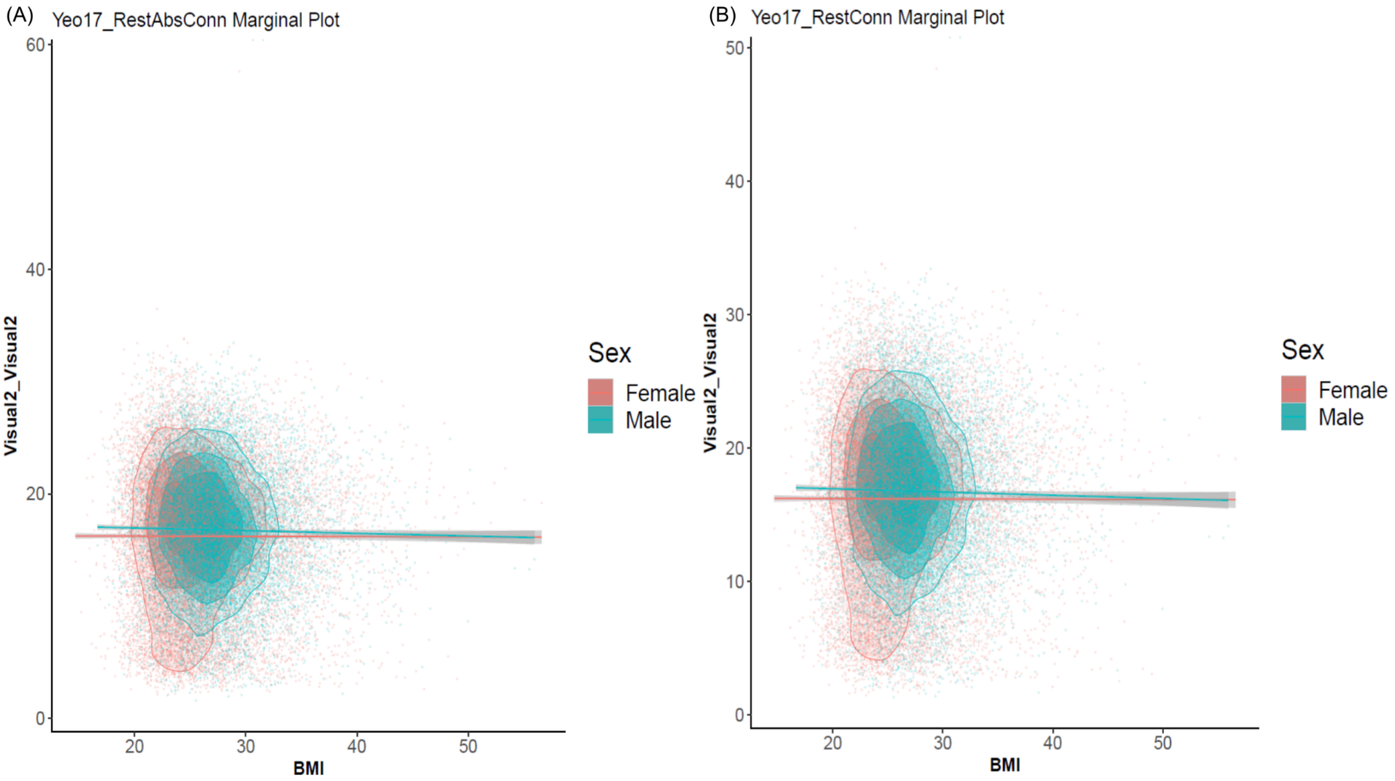


**eFigure 23. The scatterplots of (A) within-Vi2 network edge strength (NES) and (B) network-level functional connectivity (NFC) against BMI grouped by sex.** Results are based on the Yeo-17 network atlas. Contour plots are used to show the density of two-dimensional distributions; regression lines and 95% confidence intervals are also drawn. Vi2, visual 2 network.

intervals were also drawn.

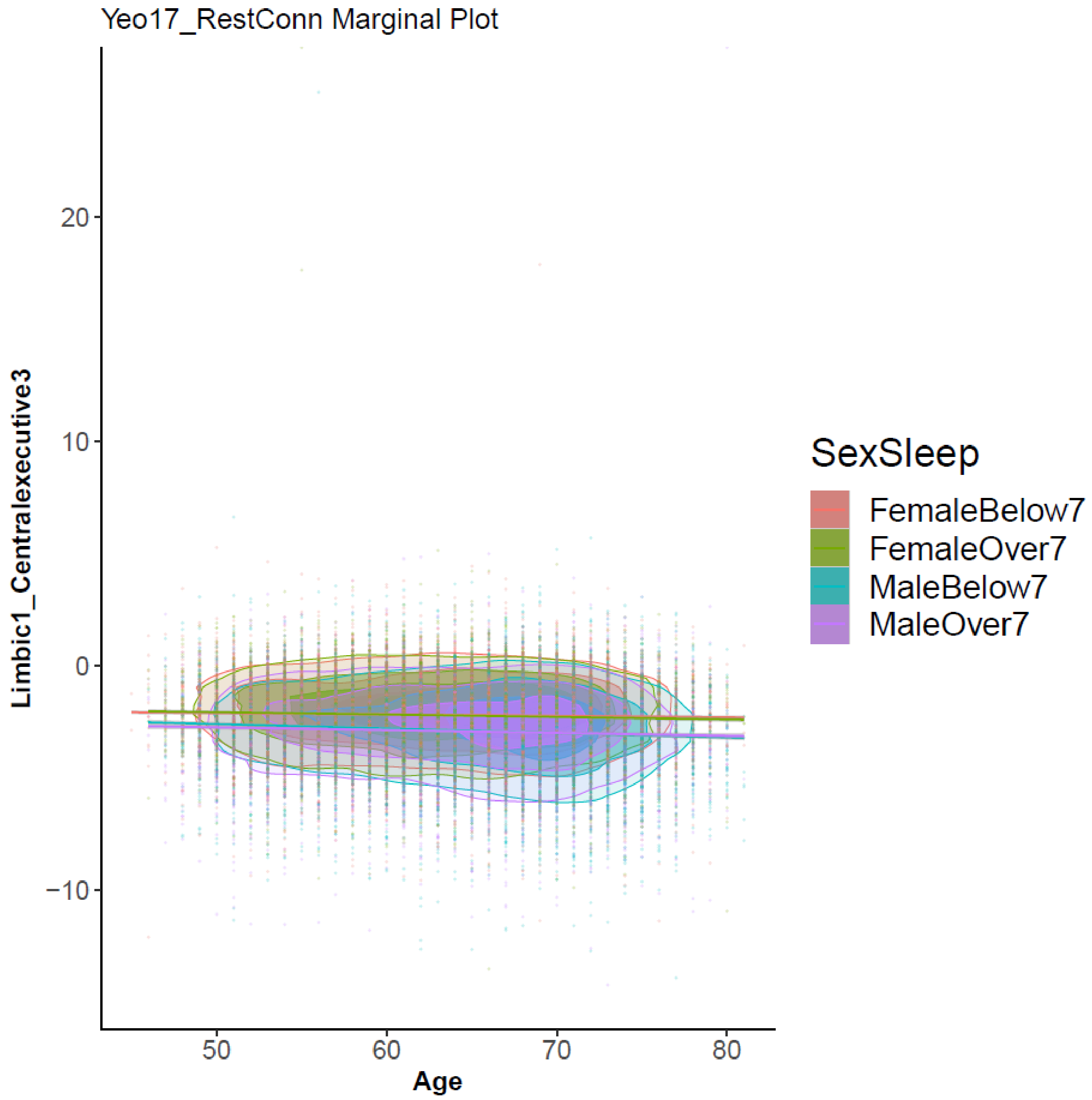


**eFigure 24. The scatterplots of LN1-CE3 between-network functional connectivity (NFC) against age grouped by sex.** Results are based on the Yeo-17 network atlas. Contour plots are used to show the density of two-dimensional distributions; regression lines and 95% confidence intervals are also drawn. LN1, limbic 1 network; CE3, central executive 3 network.

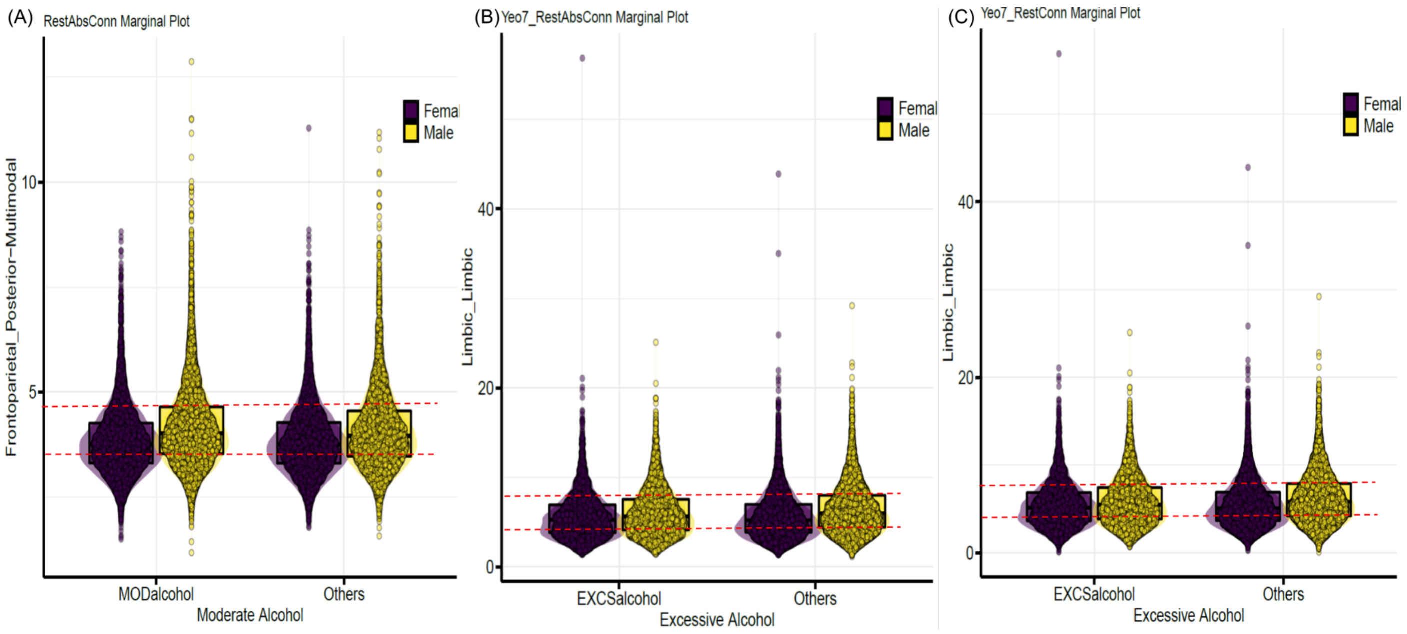


**eFigure 25. The boxplots of brain functional connectivity measures against different alcohol consumptions grouped by sex.** (A) FPN-PMN between-network edge strength (NES), (B) within-LN NES and (C) within-LN network functional connectivity (NFC), are based on the Ji-12, Yeo-7 and Yeo-7 network atlases, respectively. FPN, frontoparietal network; PMN, posterior multimodal network; LN, limbic network.

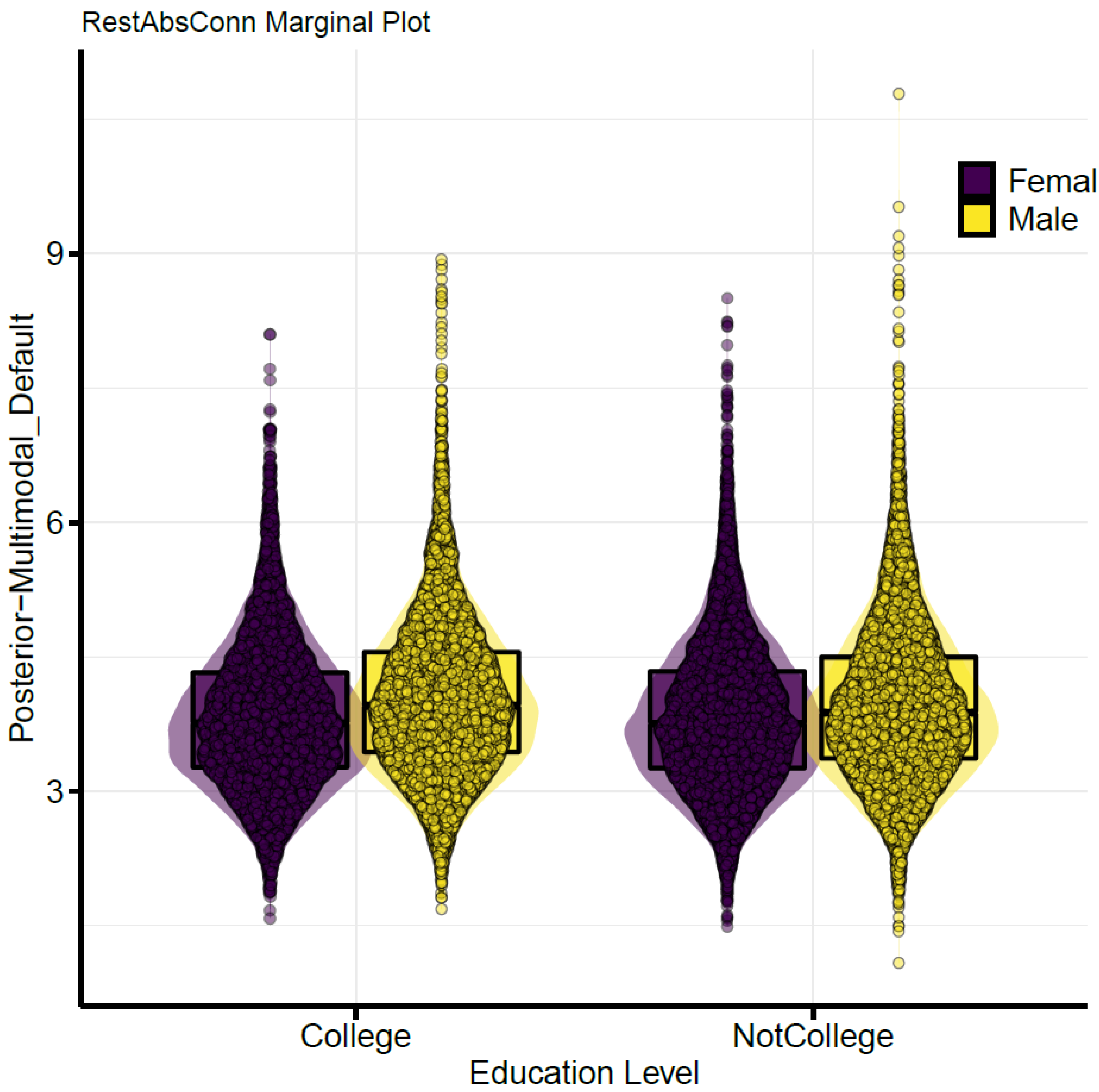


**eFigure 26. The boxplots of PMN-DAN between-network edge strength (NES) against different education levels (with or without college degree) grouped by sex.** Results are based on the Ji-12 network atlas. PMN, posterior multimodal network; DAN, dorsal attention network.

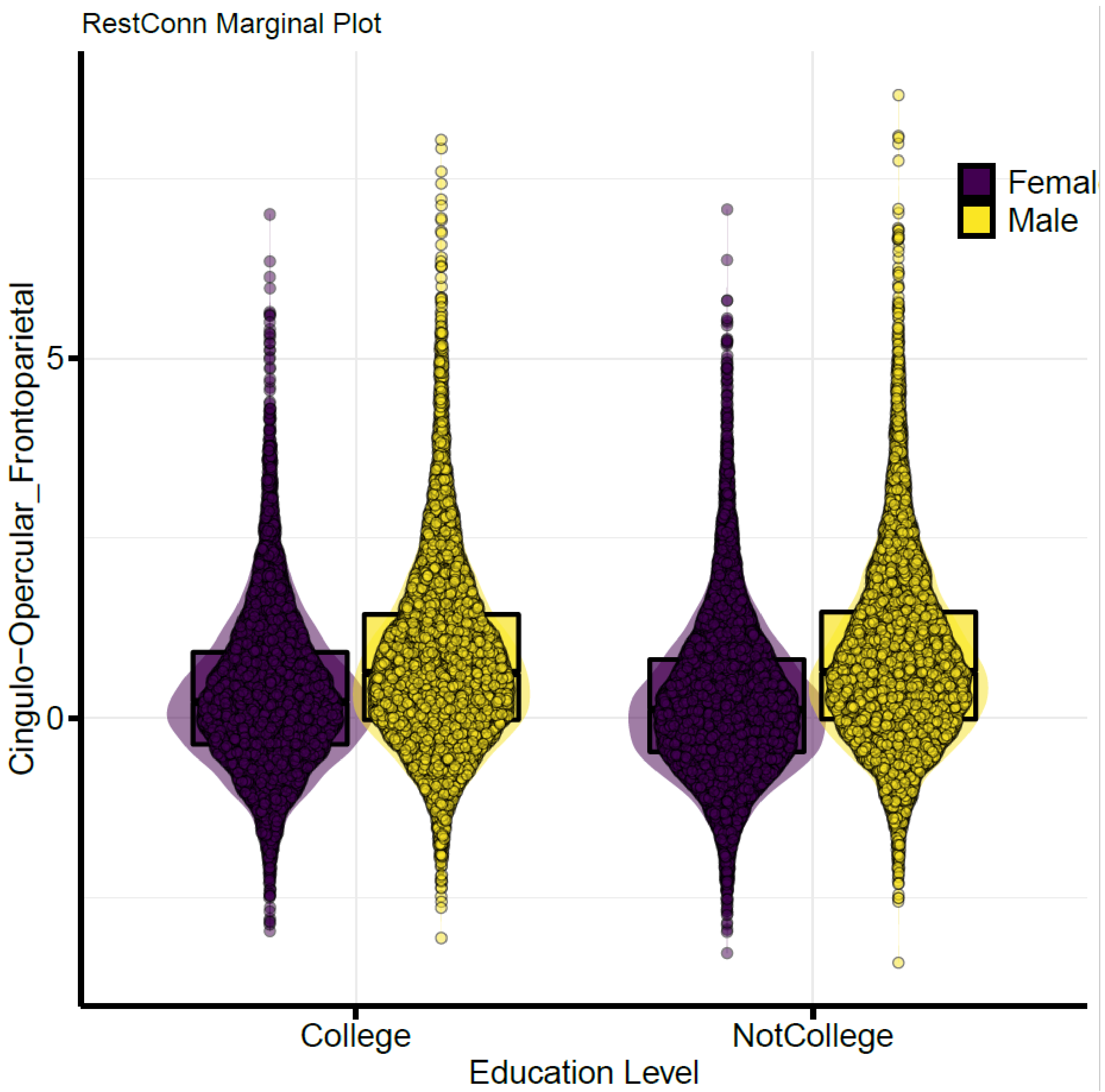


**eFigure 27. The boxplot of CON-FPN between-network functional connectivity (NFC) against different education levels (with or without college degree) grouped by sex**. Results are based on the Ji-12 network atlas. CON, cingulo-Opercular network; FPN, frontoparietal network.

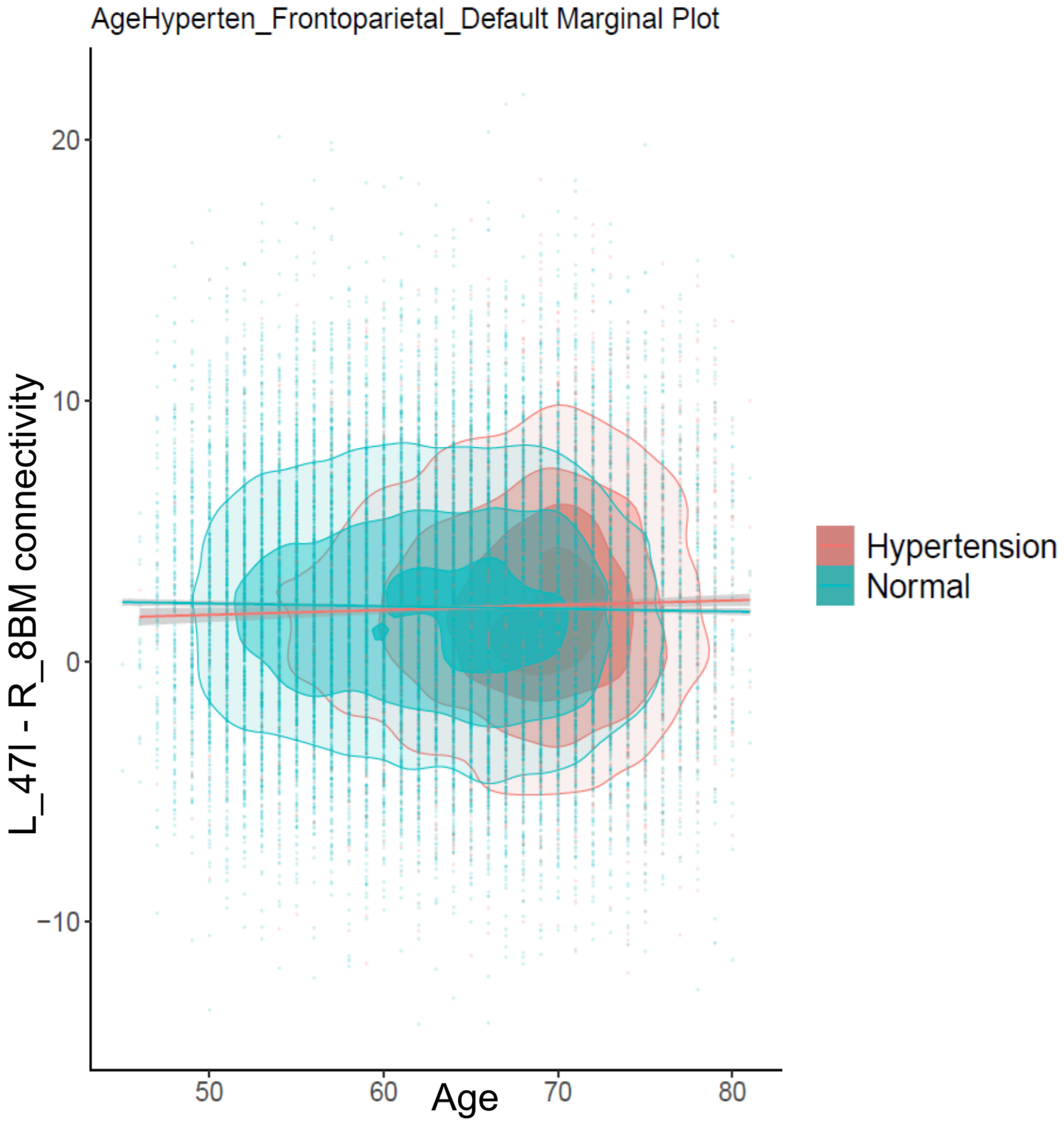


**eFigure 28. The scatterplots of brain region L_47l and R_8BM connectivity against age grouped by hypertension status.** Contour plots are used to show the density of two-dimensional distributions; regression lines and 95% confidence intervals are also drawn.

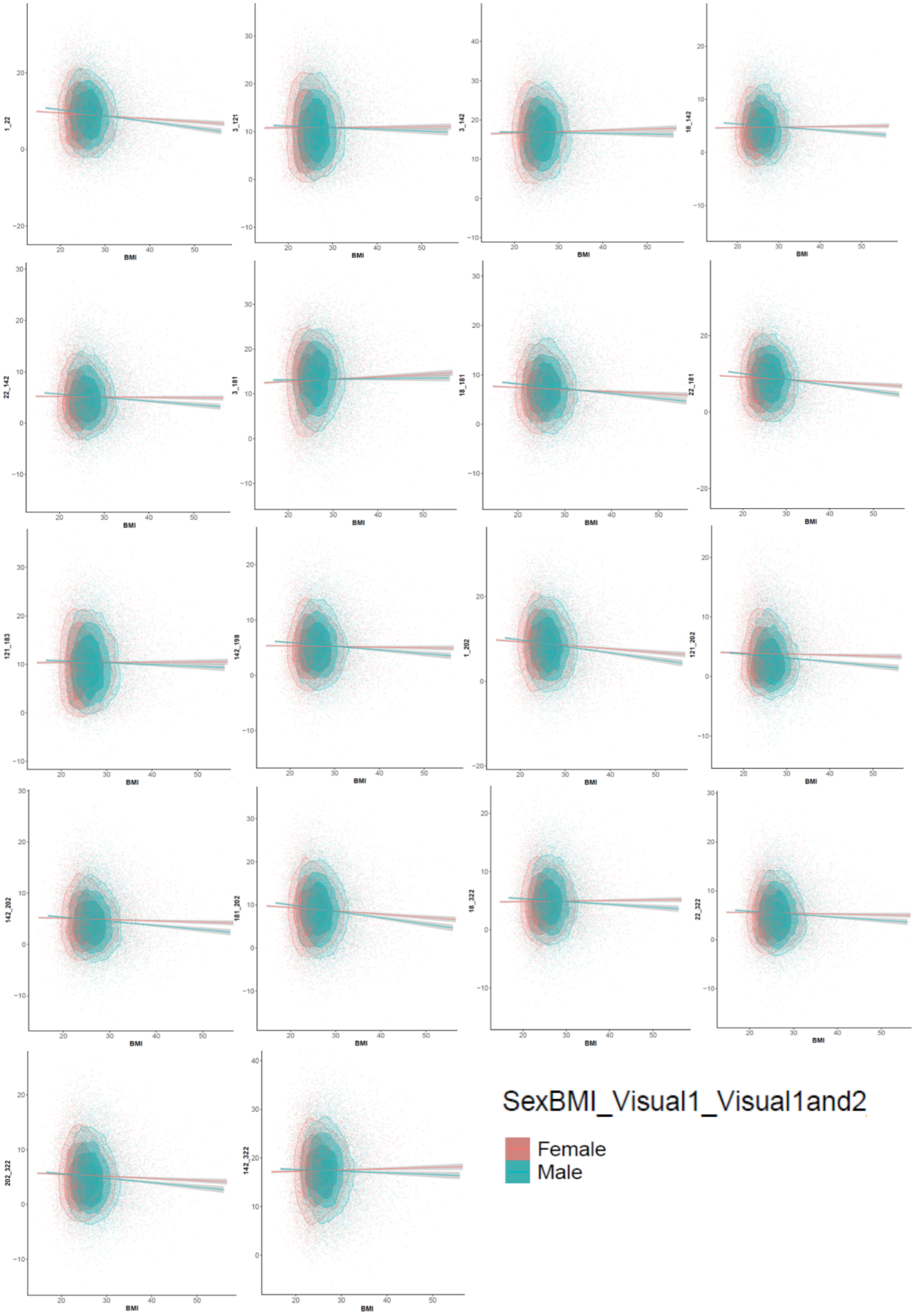


**eFigure 29. The scatterplots of brain functional connectivities between the 18 pairs of brain regions in brain visual regions (Vi1 and Vi2; Ji-12 network atlas) against BMI grouped by sex, respectively**. Contour plots are used to show the density of two-dimensional distributions; regression lines and 95% confidence intervals were also drawn. Vi1, visual 1 network; Vi2, visual 2 network.

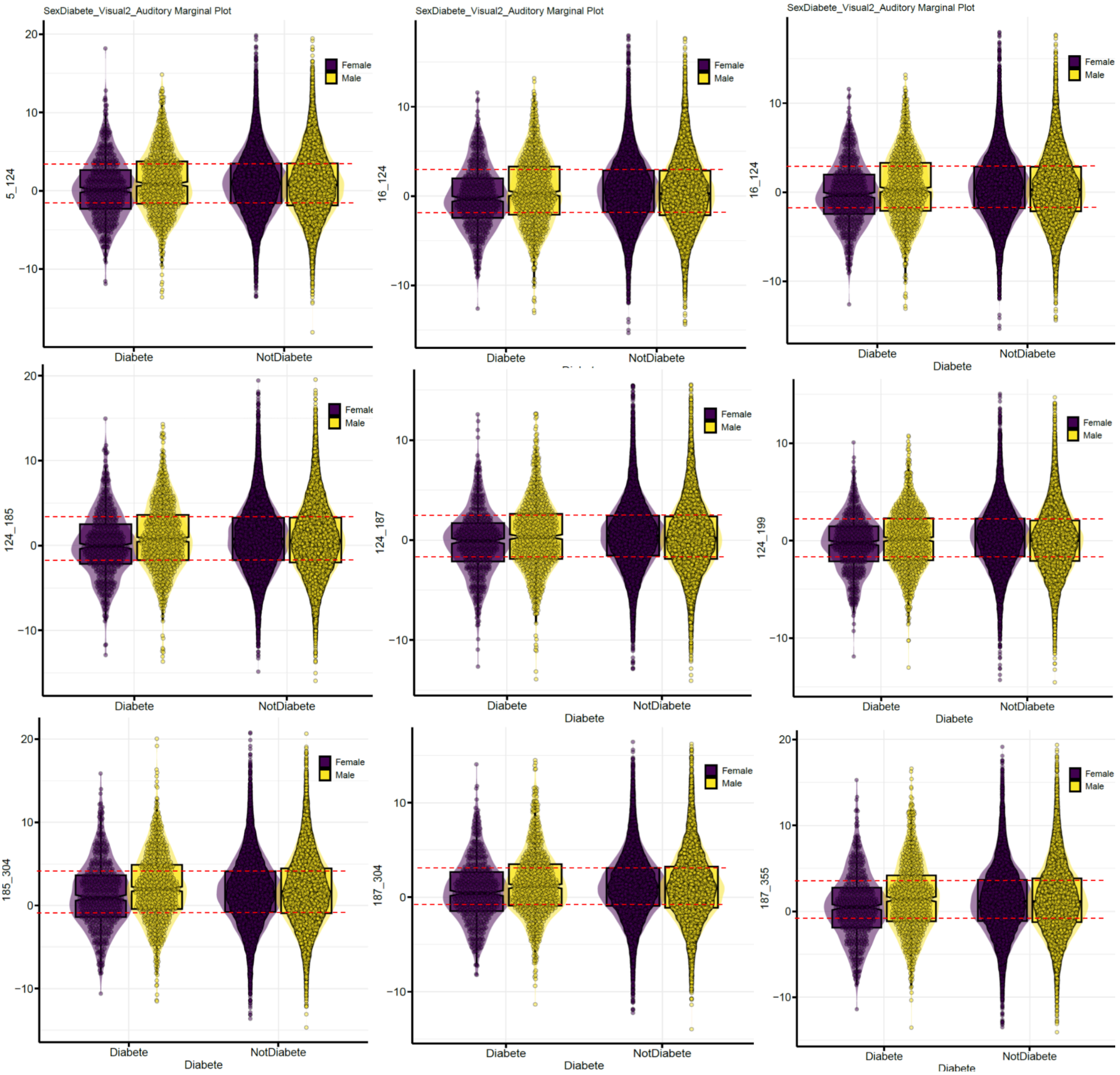


**eFigure 30. The boxplots of the brain functional connectivities for male and female subjects with and without diabetes**. Results are between 9 pairs of brain regions from Visual 2 and auditory networks (Ji-12 network atlas).

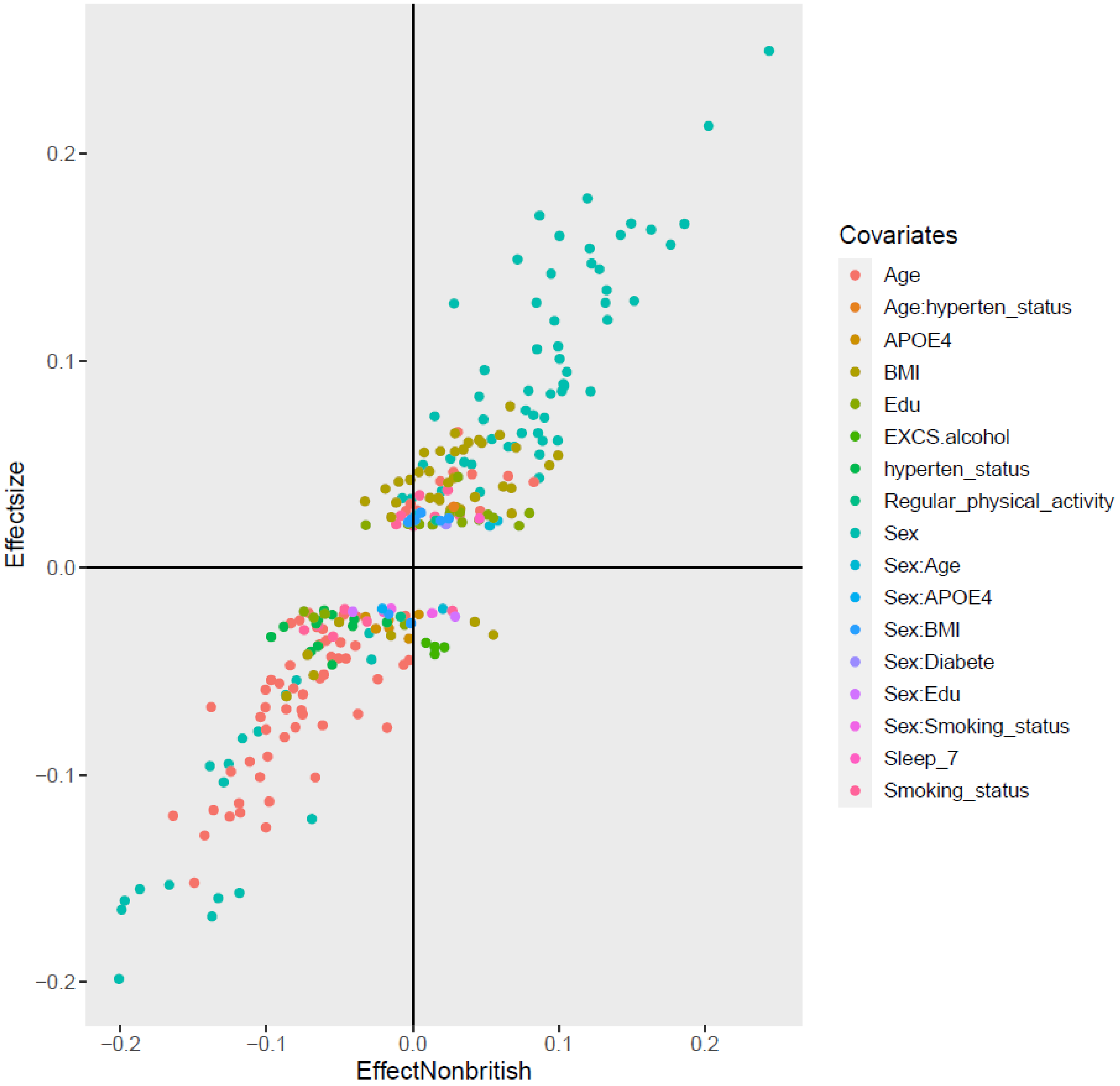


**eFigure 31. A comparison of effect sizes between British and non-British populations based on network-level functional connectivities (NFCs) using the Yeo-7 network atlas**. Results are based on the effect sizes of identified signals that pass Bonferroni-corrected significance level in the main discovery (n = 33,824) versus their corresponding effect sizes in non-British dataset (n = 2806) based on the Ji-12 network atlas. The identified effect sizes are reproducible, with a Pearson correlation of *0.92* (Spearman correlation = *0.90*).

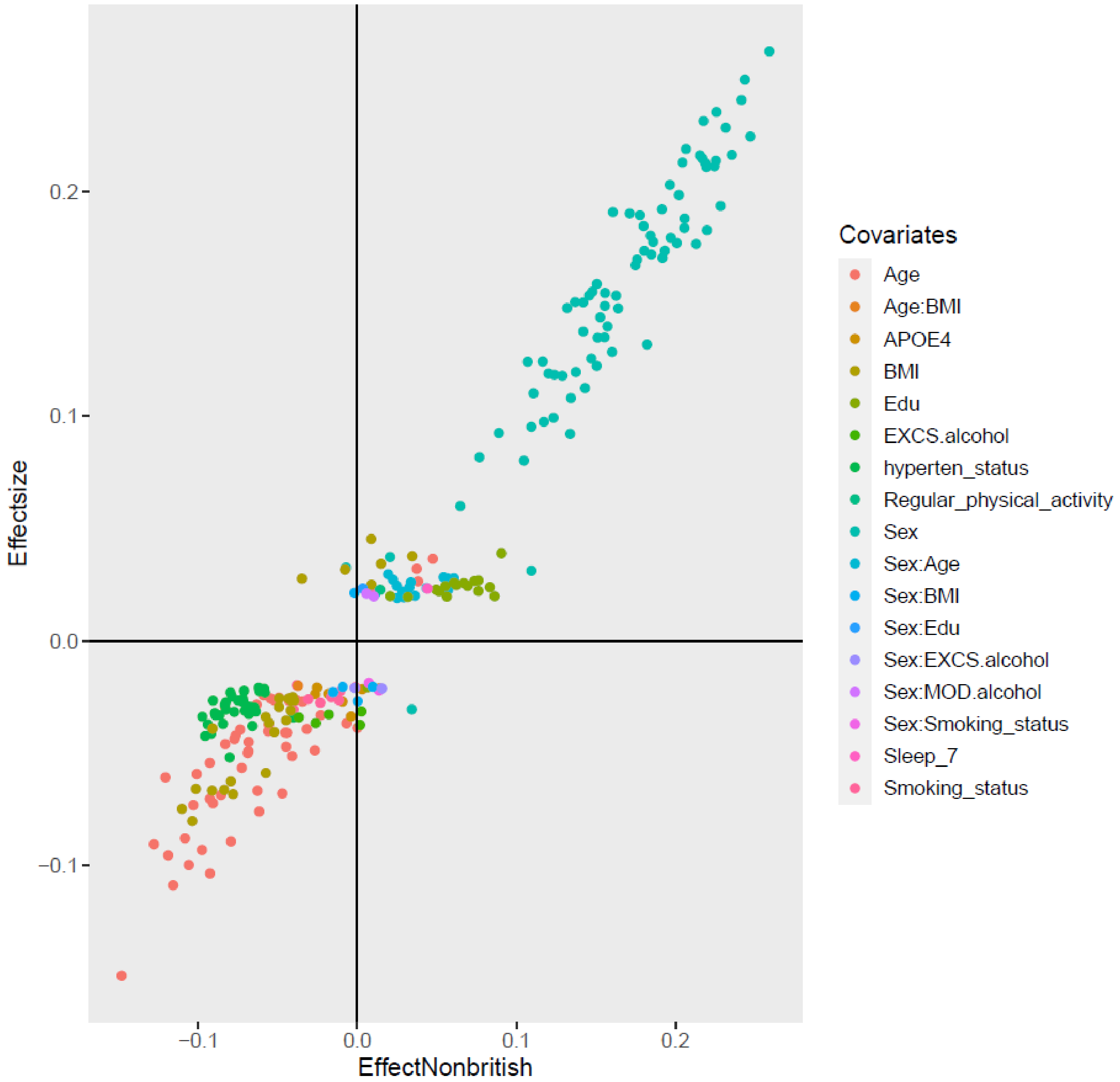


**eFigure 32. A comparison of effect sizes between British and non-British populations based on network edge strength (NES) measures using the Ji-12 network atlas**. Results are based on the effect sizes of identified signals that pass Bonferroni-corrected significance level in the main discovery (n = 33,824) versus their corresponding effect sizes in non-British dataset (n = 2806). The identified effect sizes are reproducible, with a Pearson correlation of *0.97* (Spearman correlation = *0.94*).

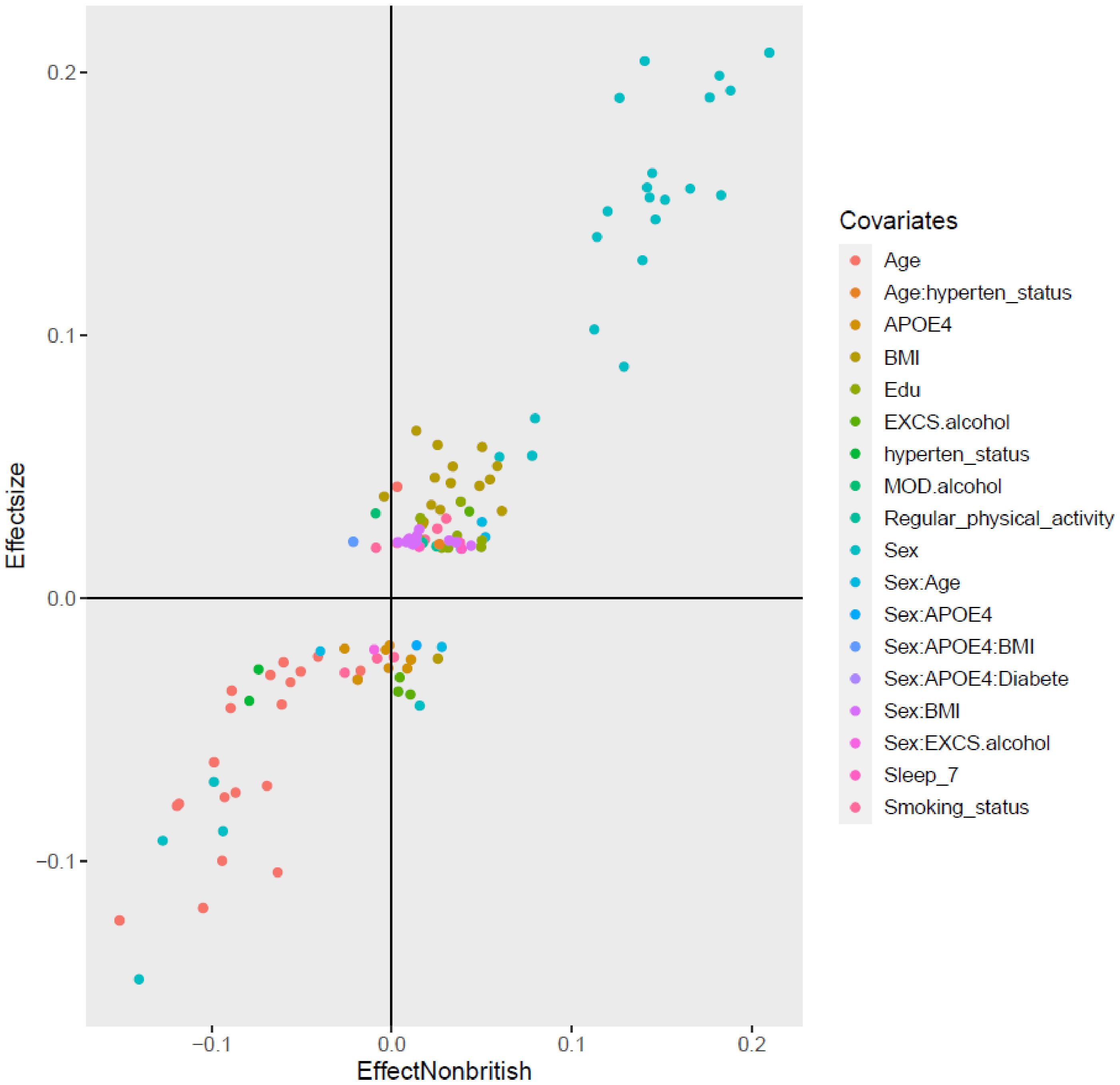


**eFigure 33. A comparison of effect sizes between British and non-British populations based on network-level functional connectivities (NFCs) using the Yeo-7 network atlas**. Results are based on the effect sizes of identified signals that pass Bonferroni-corrected significance levels in the main discovery (n = 33,824) versus their corresponding effect sizes in non-British dataset (n = 2806). The identified effect sizes are reproducible, with a Pearson correlation of *0.94* (Spearman correlation = *0.87*).

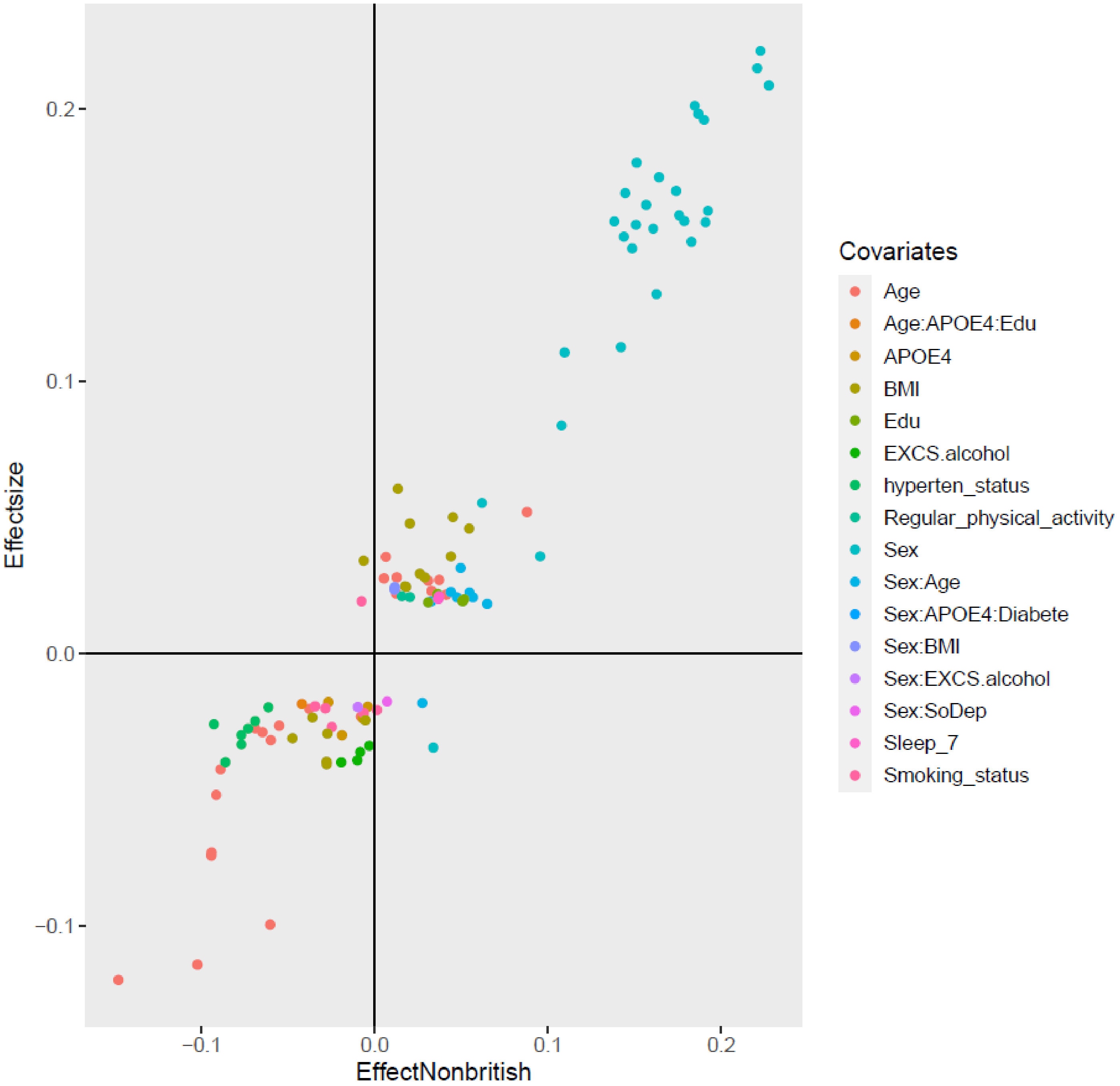


**eFigure 34. A comparison of effect sizes between British and non-British populations based on network edge strength (NES) measures using the Yeo-7 network atlas**. Results are based on the effect sizes of identified signals that pass Bonferroni-corrected significance levels in the main discovery (n = 33,824) versus their corresponding effect sizes in non-British dataset (n = 2806). The identified effect sizes are reproducible, with a Pearson correlation of *0.95* (Spearman correlation = *0.89*).

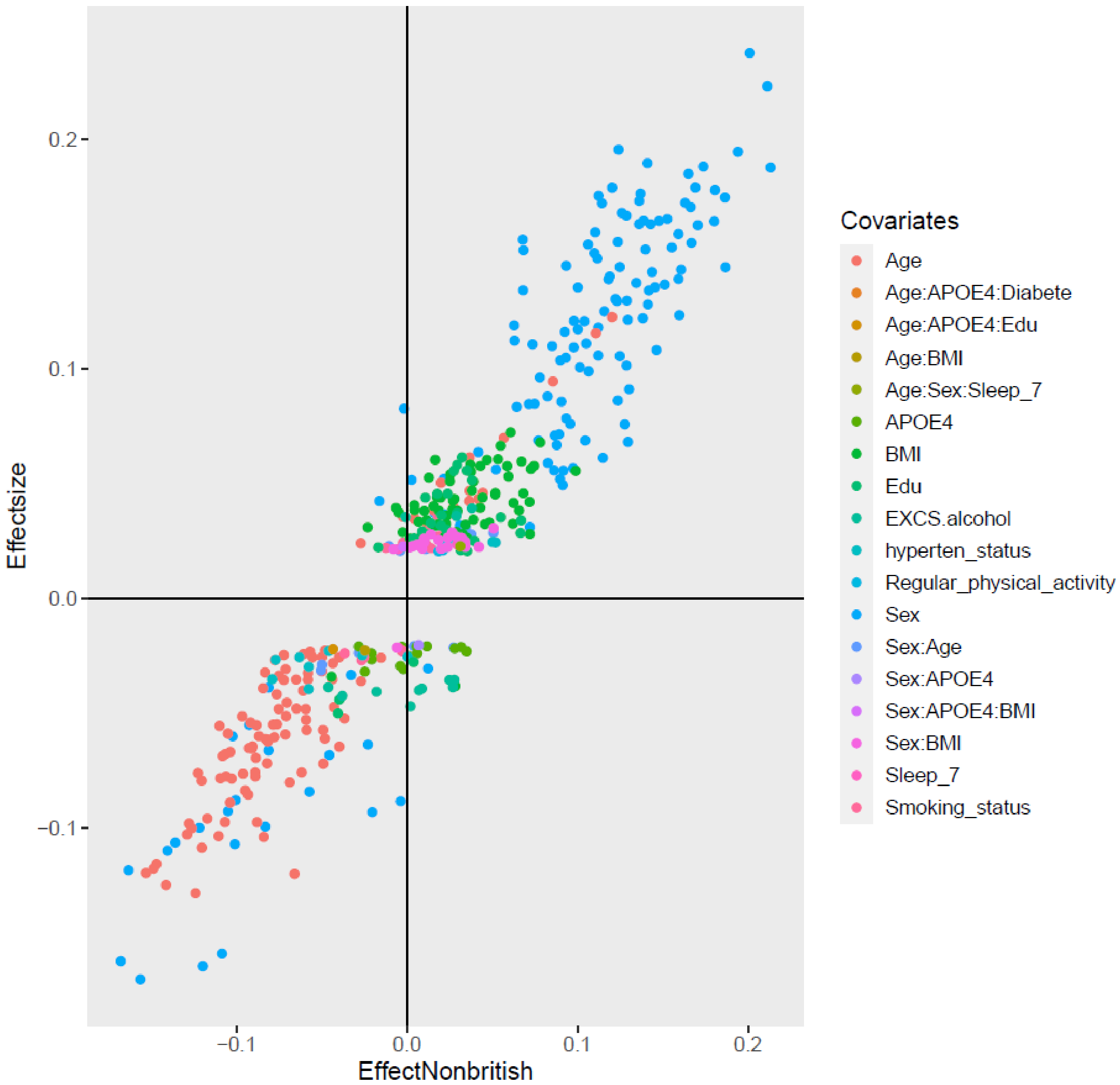


**eFigure 35. A comparison of effect sizes between British and non-British populations based on network-level functional connectivities (NFCs) using the Yeo-17 network atlas**. Results are based on the effect sizes of identified signals that pass Bonferroni-corrected significance levels in the main discovery (n = 33,824) versus their corresponding effect sizes in non-British dataset (n = 2806). The identified effect sizes are reproducible, with a Pearson correlation of *0.94* (Spearman correlation = *0.93*).

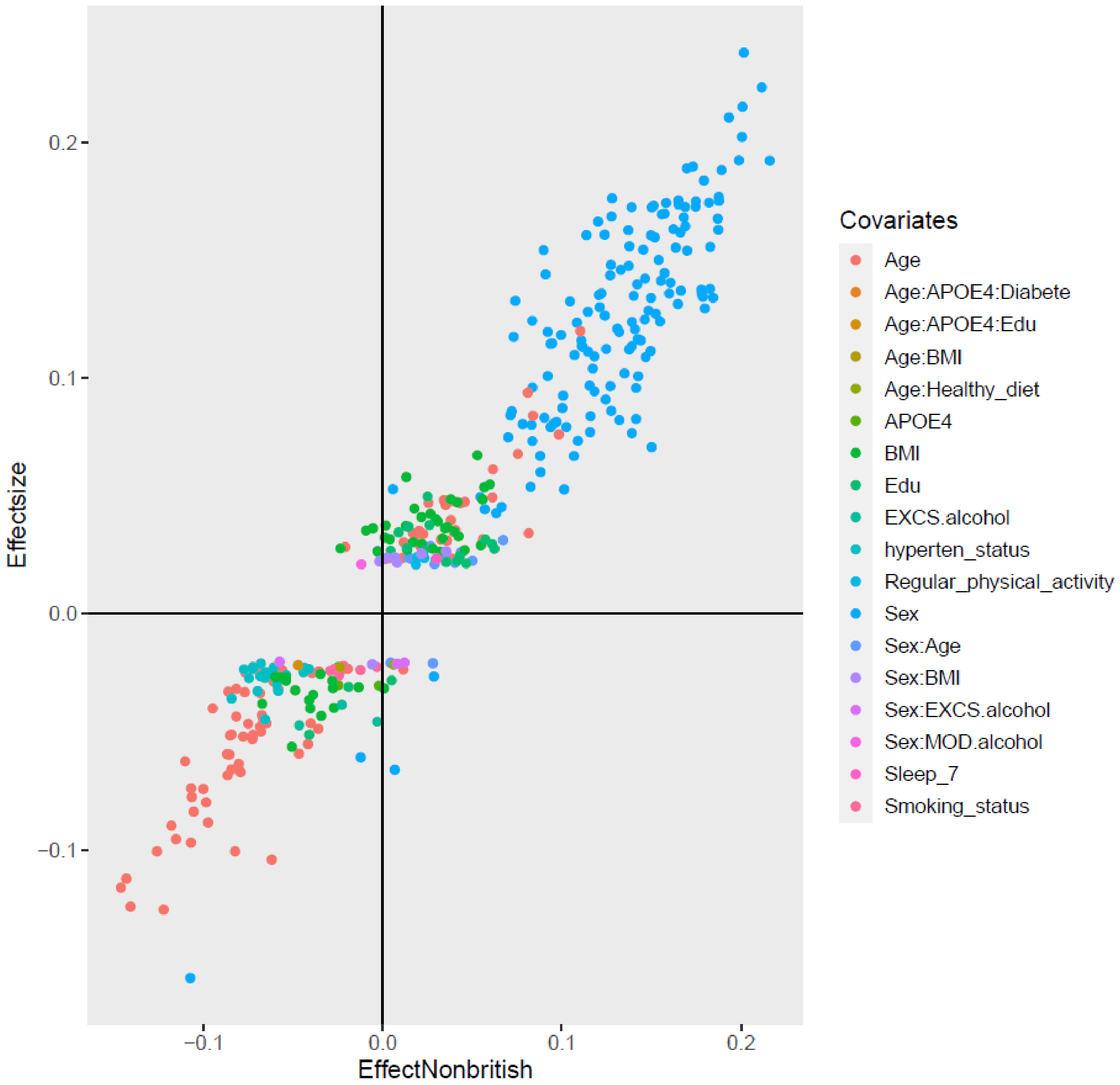


**eFigure 36. A comparison of effect sizes between British and non-British populations based on network edge strength (NES) measures using the Yeo-17 network atlas**. Results are based on the effect sizes of identified signals that pass Bonferroni-corrected significance levels in the main discovery (n = 33,824) versus their corresponding effect sizes in non-British dataset (n = 2806). The identified effect sizes are reproducible, with a Pearson correlation of *0.96* (Spearman correlation = *0.95*).

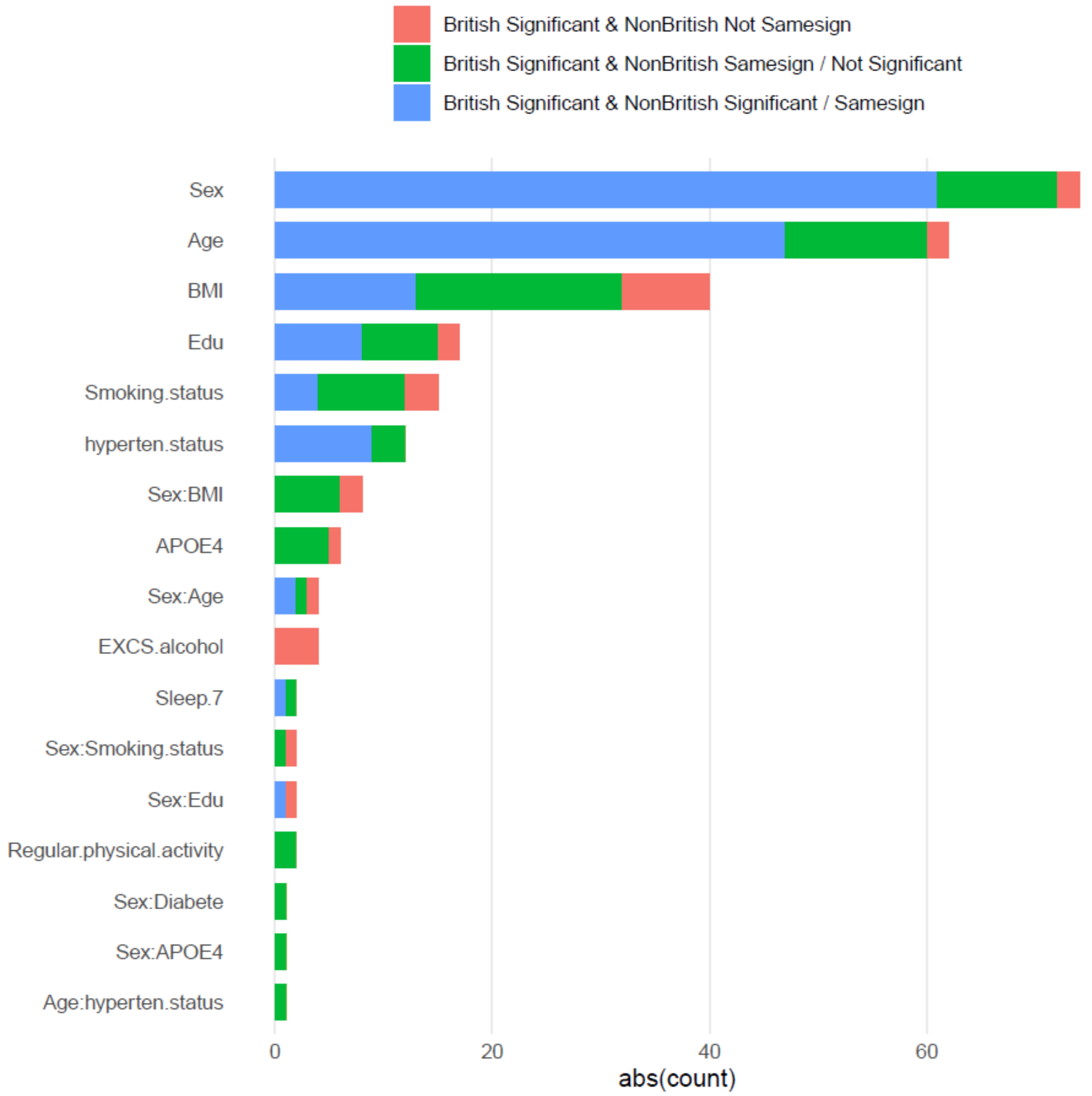


**eFigure 37. Barplots of number of signals on network-level functional connectivities (NFCs) using Ji-12 network atlas.** Results are based on the Bonferroni significance level in the main discovery (n = 33,824) that have inconsistent directions (red) in non-British dataset (n = 2806), have consistent directions that are significant (blue) or non-significant (green).

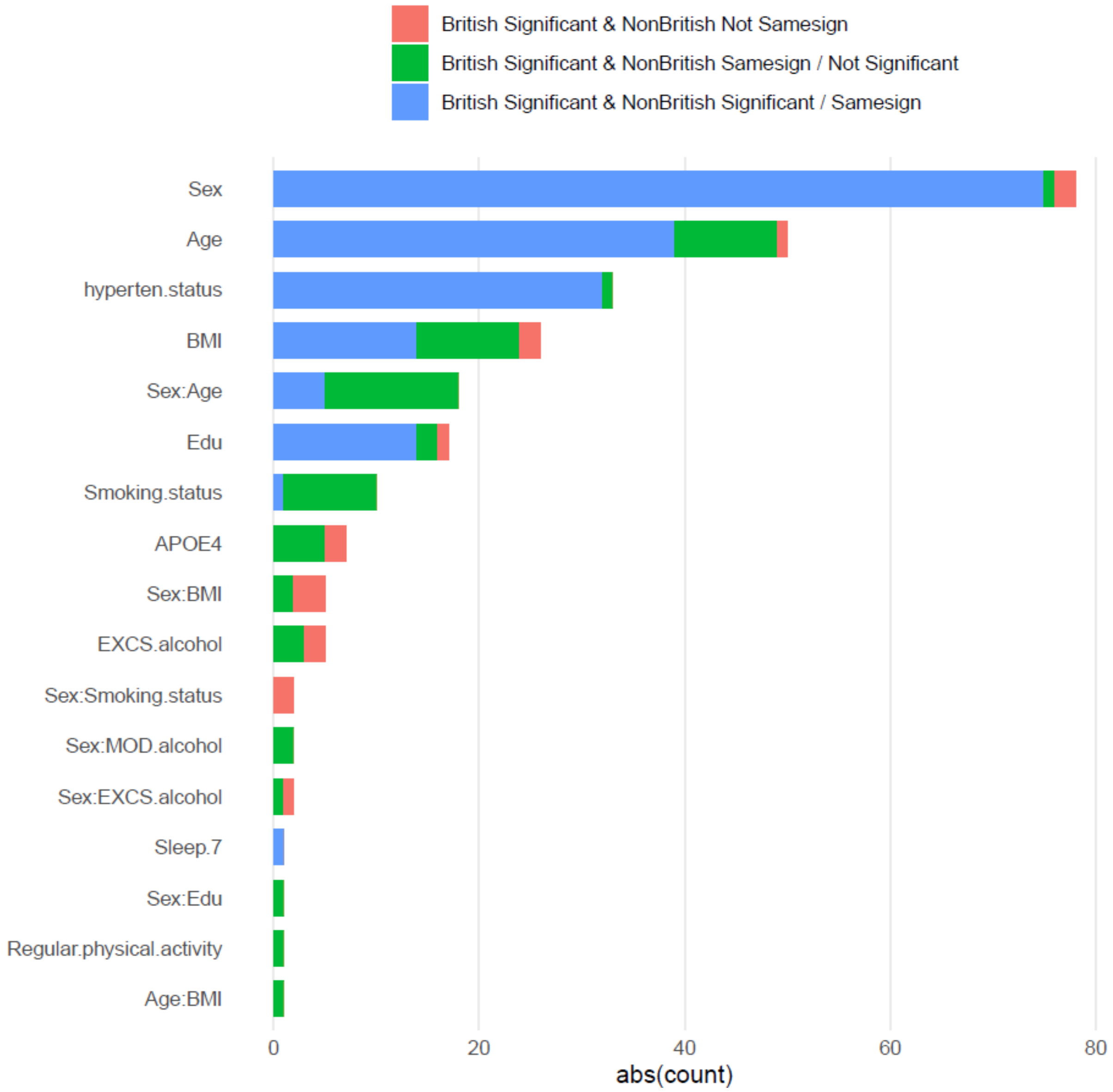


**eFigure 38. Barplots of number of signals on network edge strength (NESs) using Ji-12 network atlas**. Results are based on the Bonferroni significance level in the main discovery (n = 33,824) that have inconsistent directions (blue) in non-British dataset (n = 2806), have consistent directions that are significant (green) or non-significant (red).

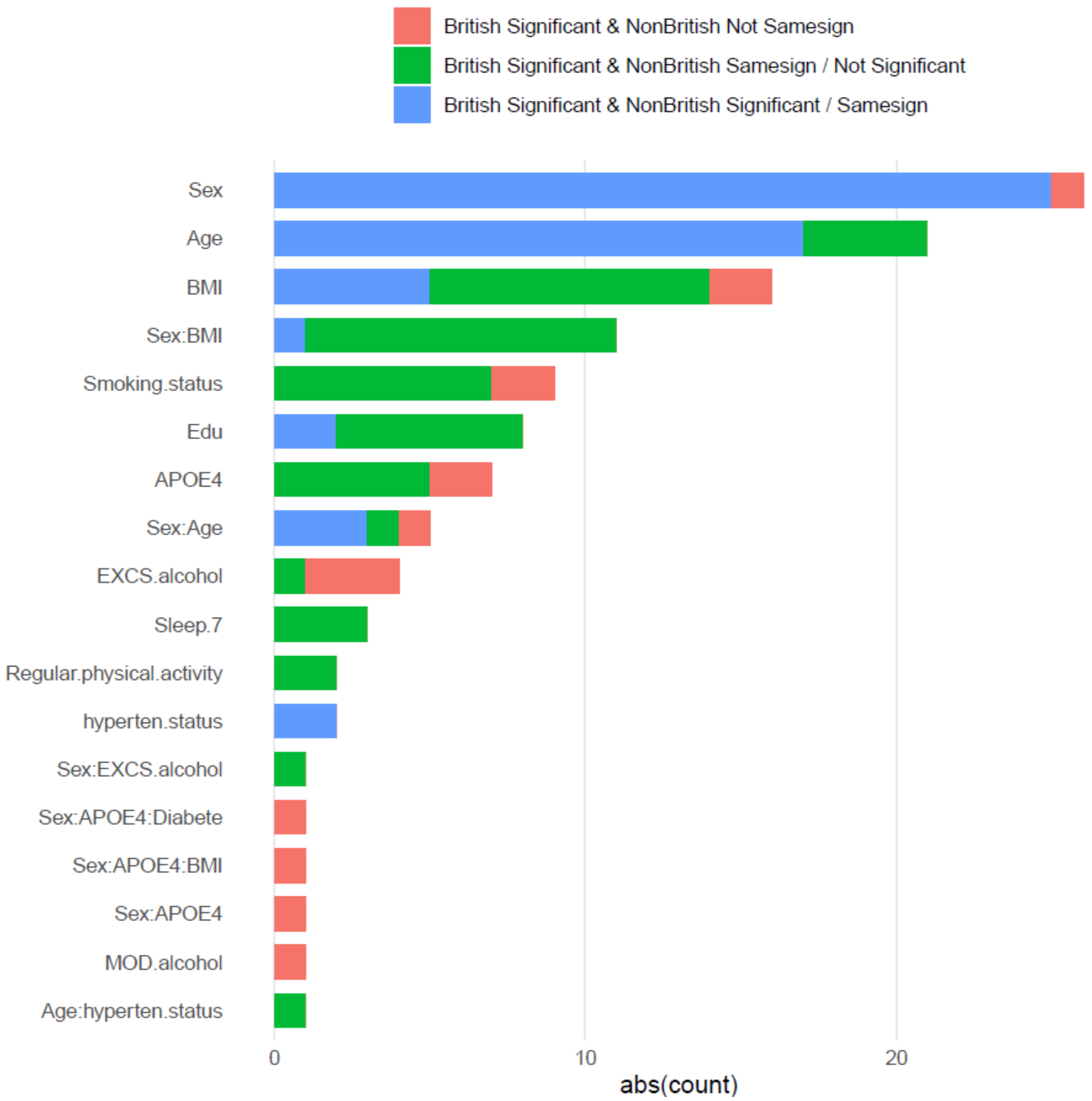


**eFigure 39. Barplots of number of signals on network-level functional connectivities (NFCs) using Yeo-7 network atlas.** Results are based on the Bonferroni significance level in the main discovery (n = 33,824) that have inconsistent directions (red) in non-British dataset (n = 2806), have consistent directions that are significant (blue) or non-significant (green).

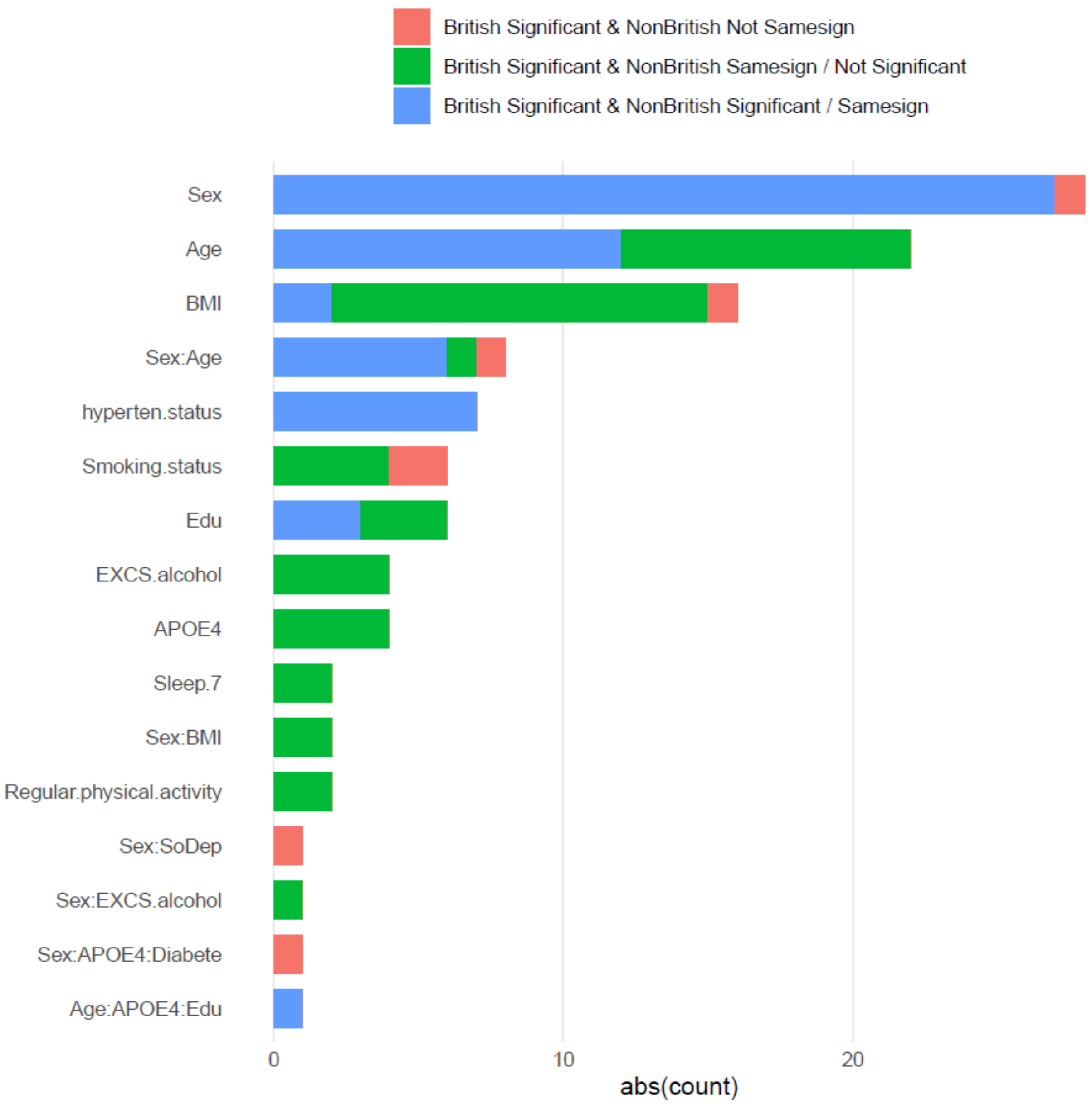


**eFigure 40. Barplots of number of signals on network edge strength (NESs) using Yeo-7 network atlas**. Results are based on the Bonferroni significance level in the main discovery (n = 33,824) that have inconsistent directions (red) in non-British dataset (n = 2806), have consistent directions that are significant (blue) or non-significant (green).

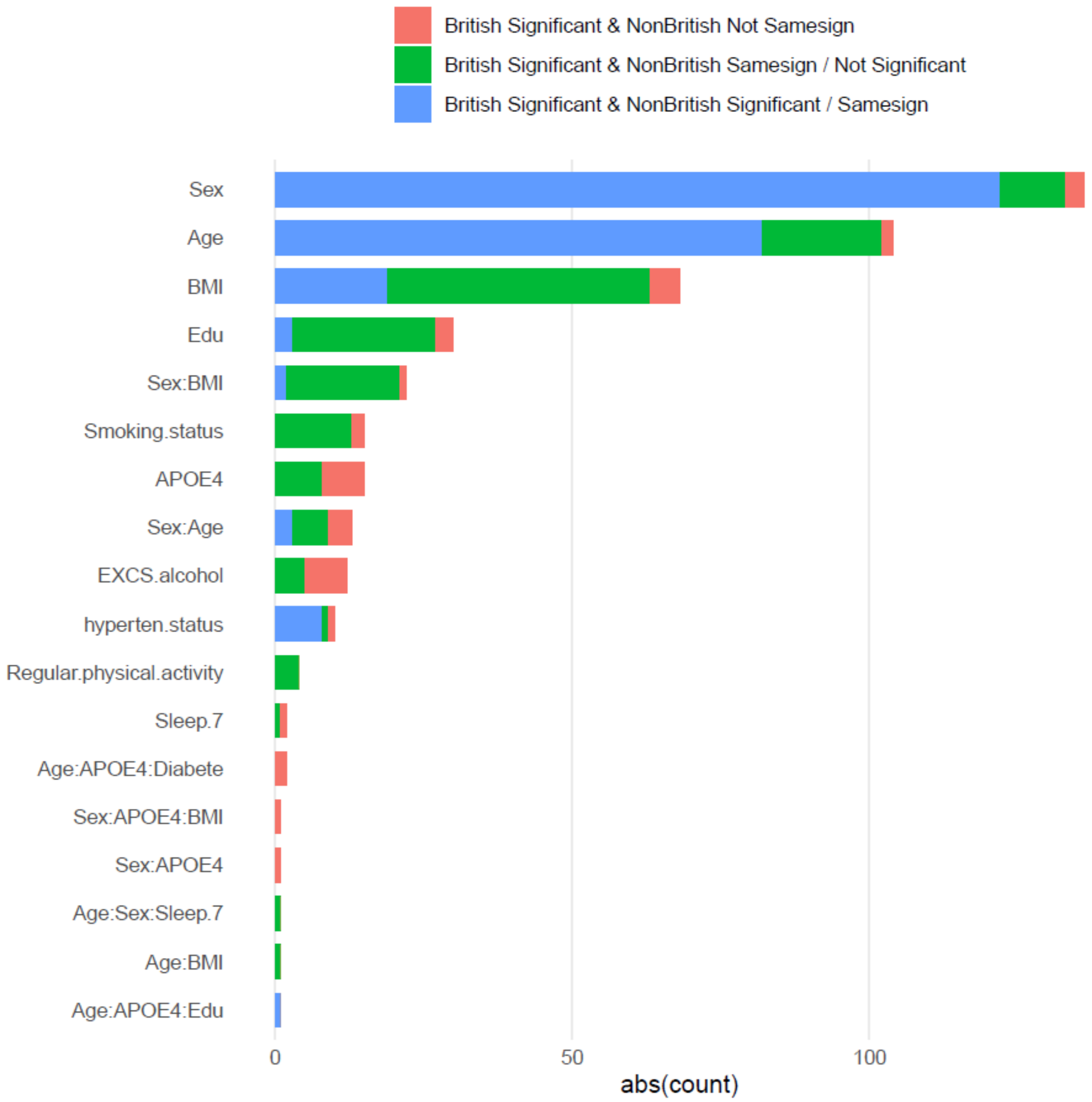


**eFigure 41. Barplots of number of signals on network-level functional connectivities (NFCs) using Yeo-17 network atlas**. Results are based on the Bonferroni significance level in the main discovery (n = 33,824) that have inconsistent directions (red) in non-British dataset (n = 2806), have consistent directions that are significant (blue) or non-significant (green).

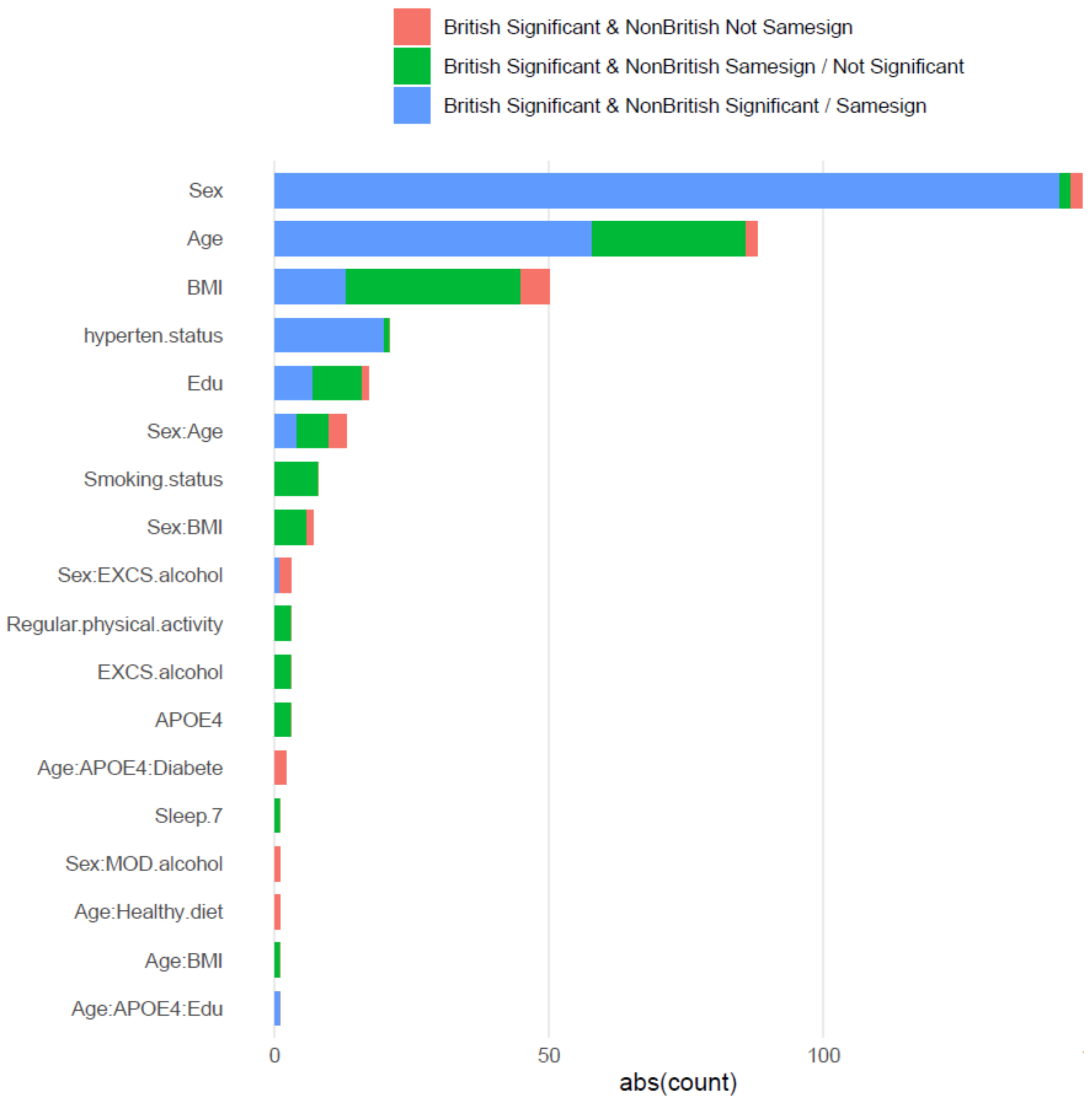


**eFigure 42. Barplots of number of signals on network edge strength (NESs) using Yeo-17 network atlas**. Results are based on the Bonferroni significance level in the main discovery (n = 33,824) that have inconsistent directions (red) in non-British dataset (n = 2806), have consistent directions that are significant (blue) or non-significant (green).

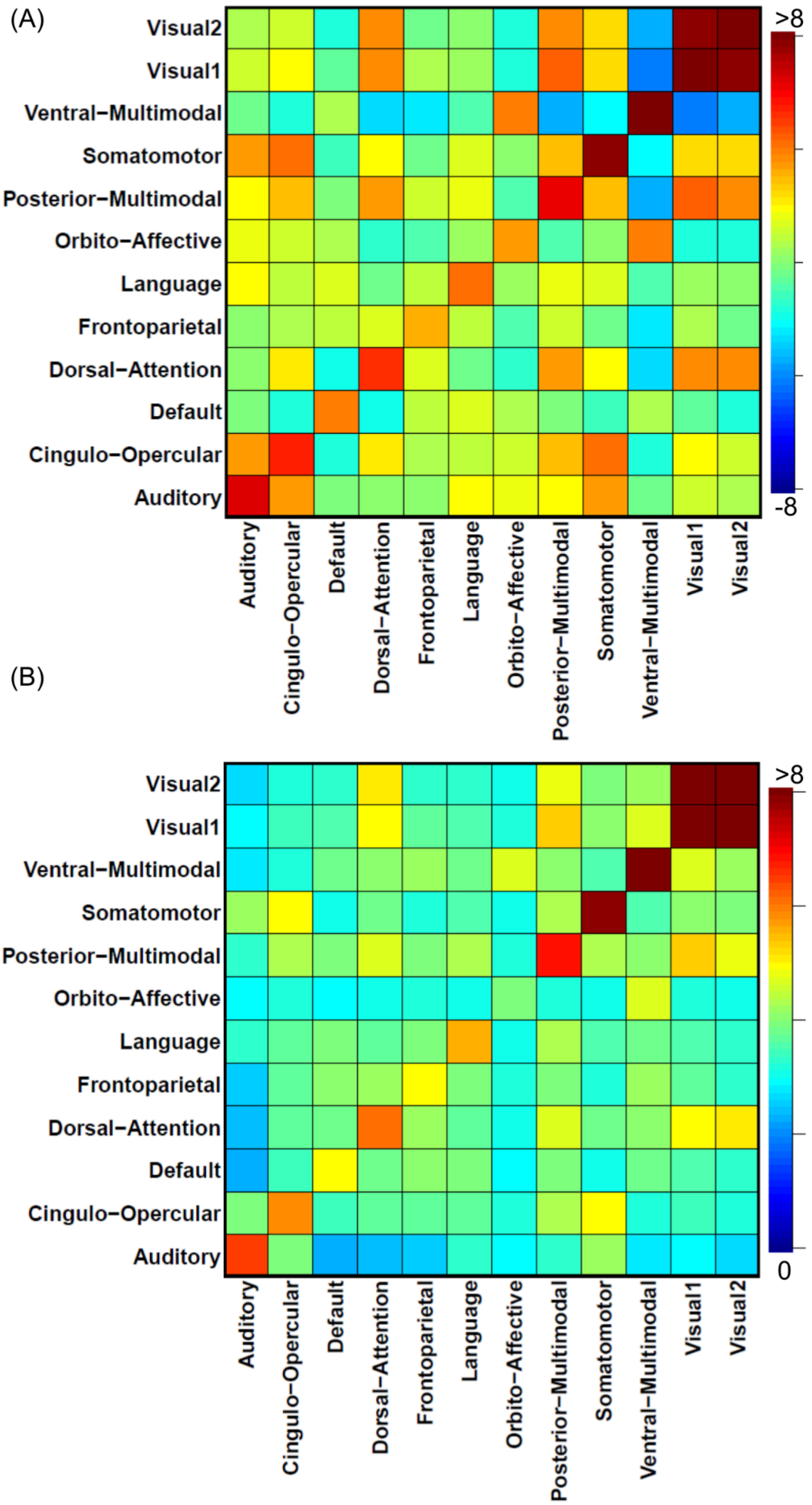


**eFigure 43. Heatmap to illustrate the mean network-level functional connectivity (NFC) (A) and network edge strength (NES) (B) matrices based on the Ji-12 network atlas across the population (36,630 subjects).**

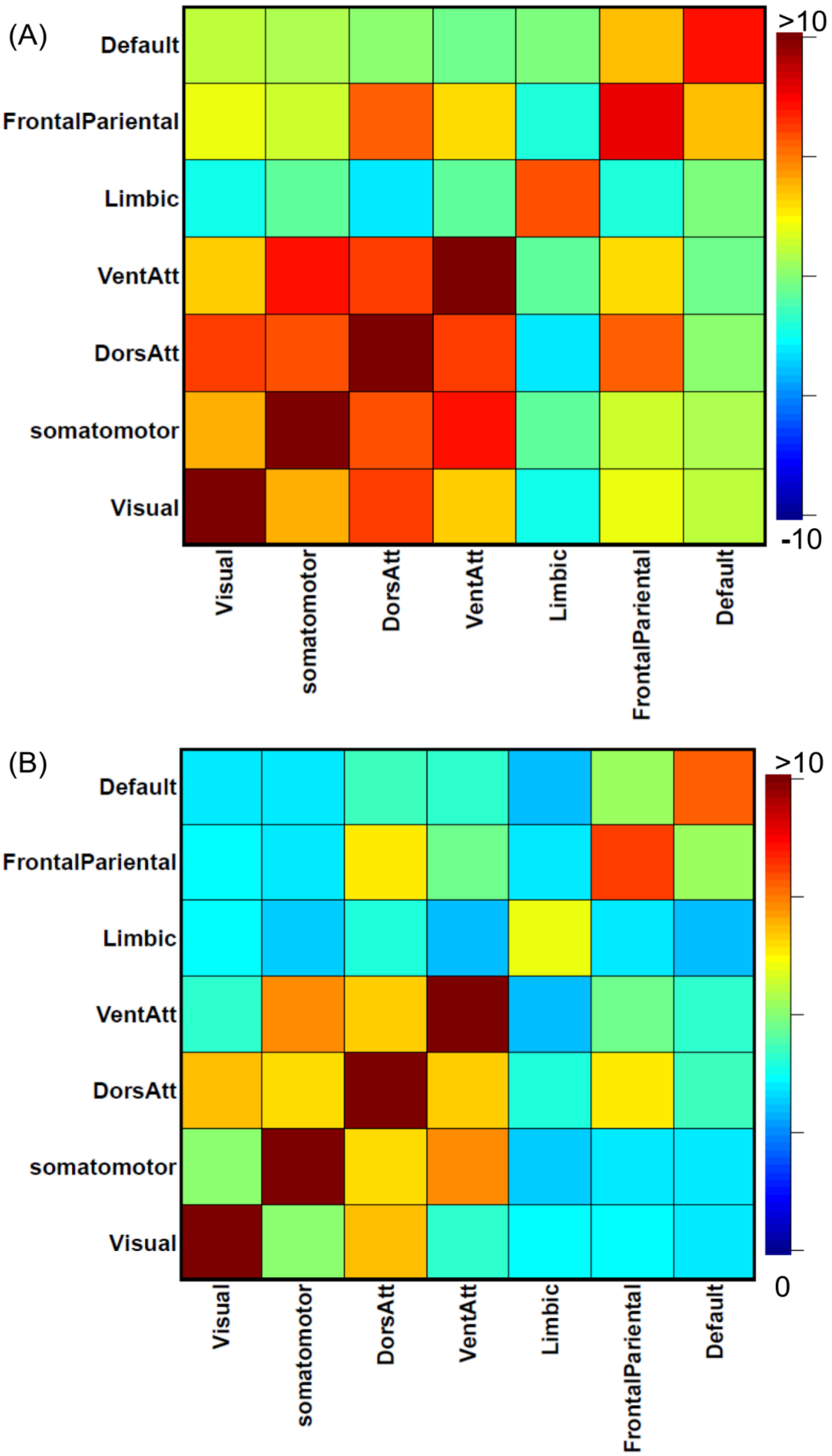


**eFigure 44. Heatmap to illustrate the population-mean network-level functional connectivity (NFC) (A) and network edge strength (NES) (B) matrices based on the Yeo-7 network atlas across the population (36,630 subjects).**

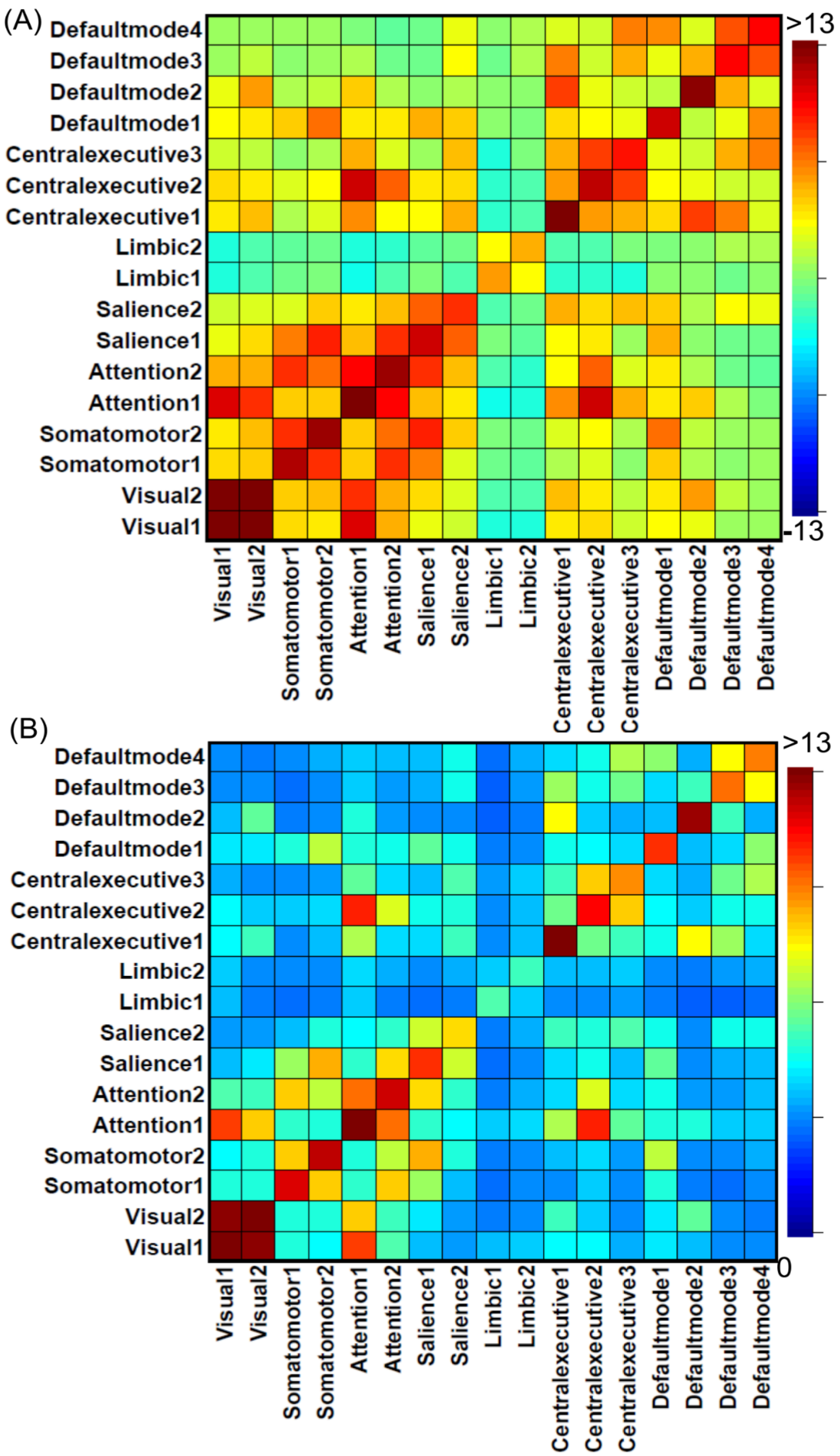


eFigure 45. Heatmap to illustrate the population-mean network-level functional connectivity (NFC) (A) and network edge strength (NES) (B) matrices based on the Yeo-17 network atlas across the population (36,630 subjects).

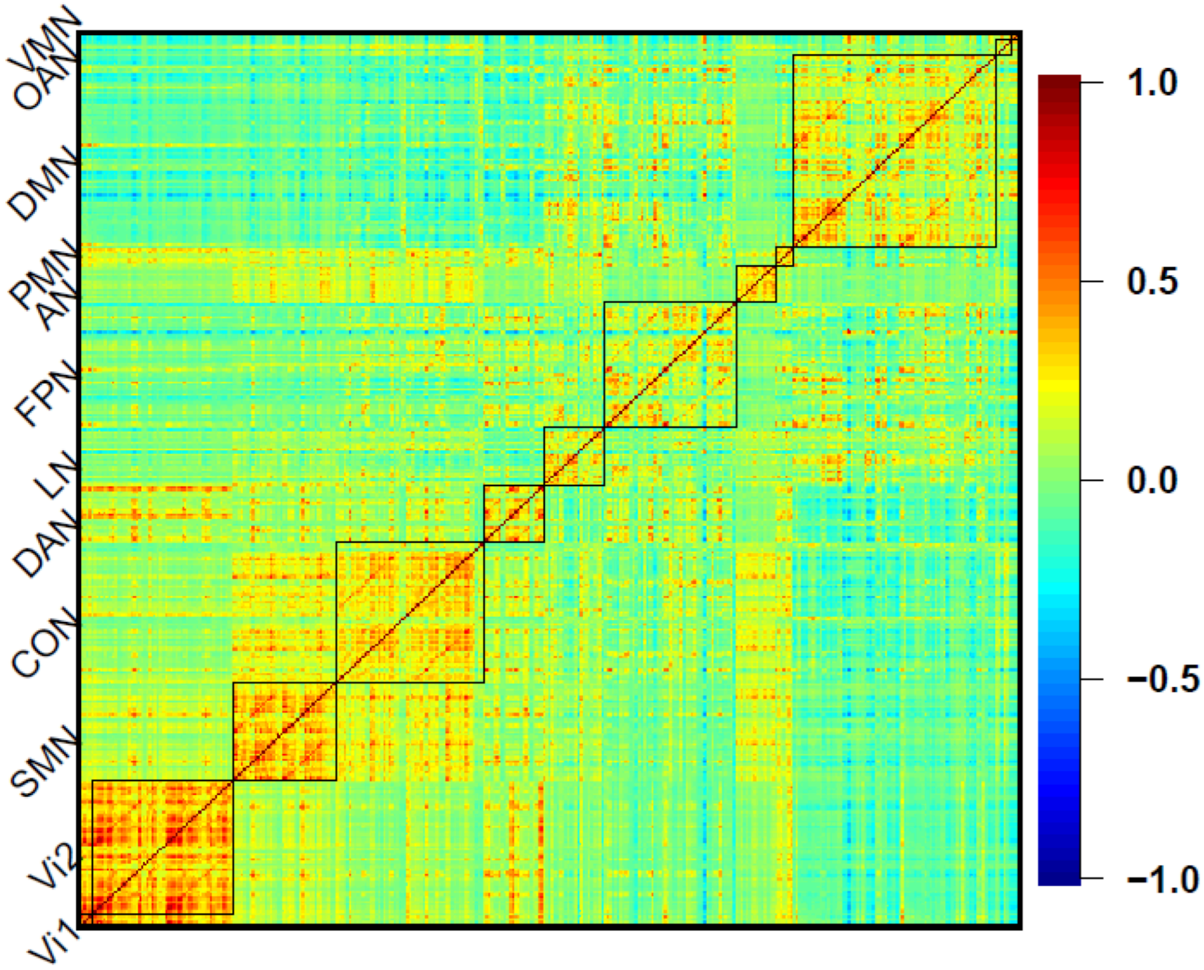


**eFigure 46. The functional connectivity map of the HCP-MMP atlas and Ji-12 network atlas across the population (36,630 subjects)**. The regions are organized to show region pairs within the same network together.

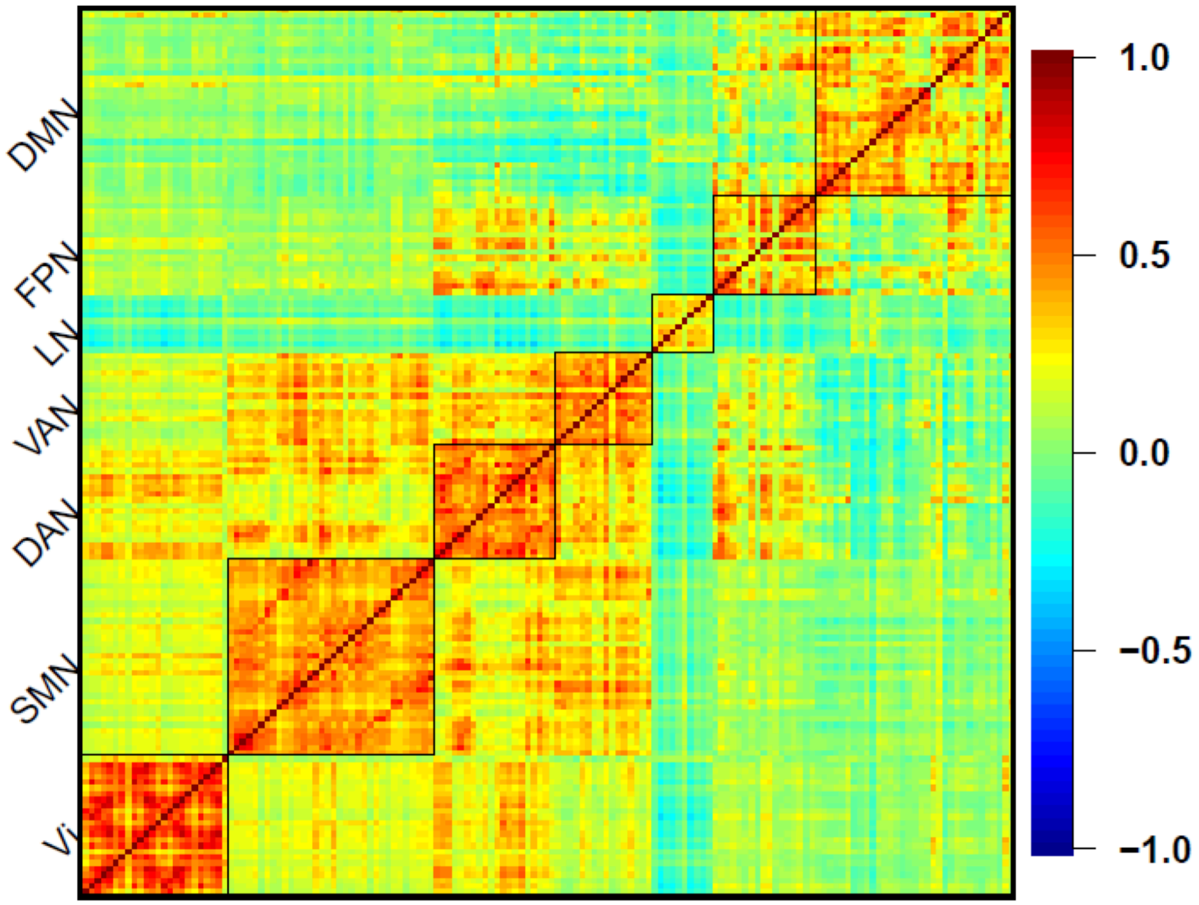


**eFigure 47. The functional connectivity map of the Schaefer-200 atlas and Yeo-7 network atlas across the population (36,630 subjects)**. The regions are organized to show region pairs within the same network together.

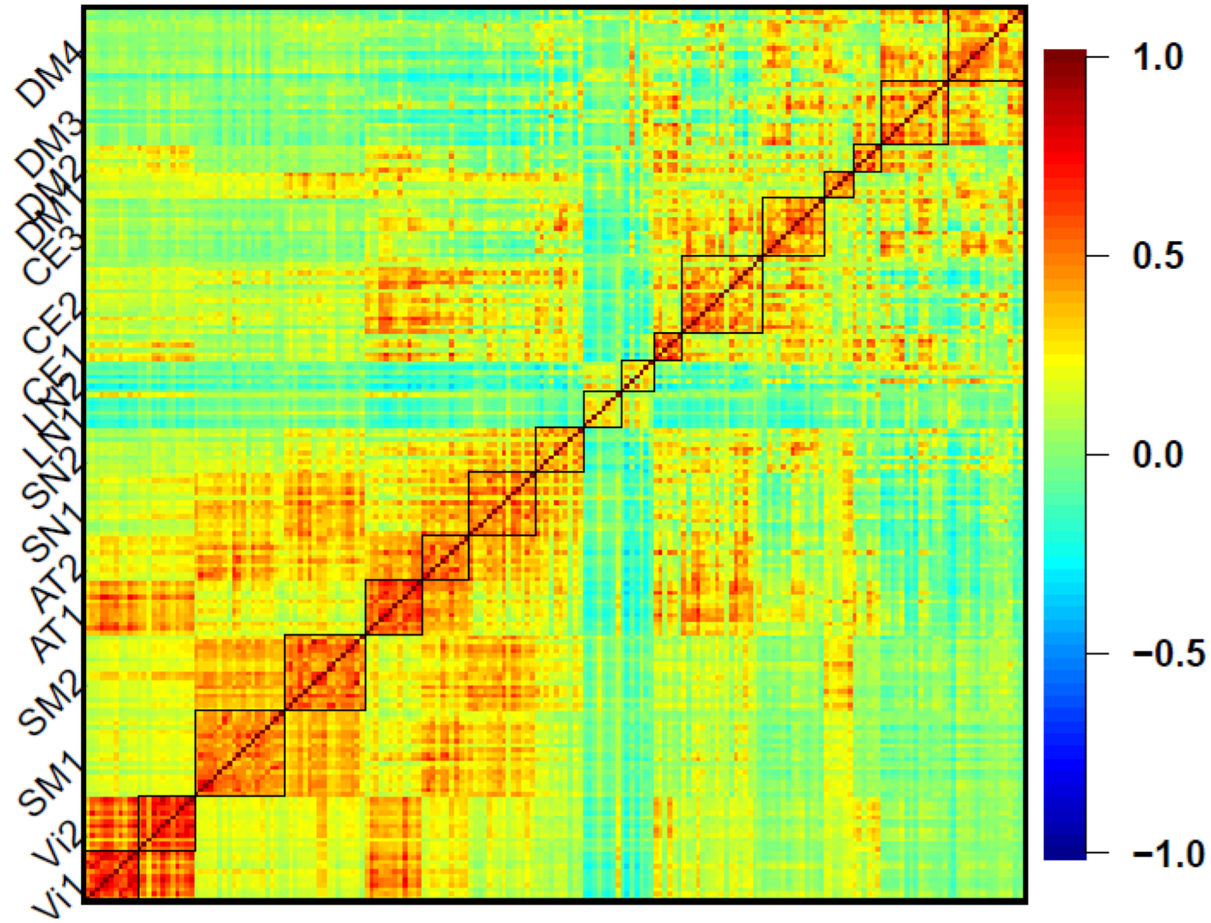


**eFigure 48. The functional connectivity map of the Schaefer-200 atlas and Yeo-17 network atlas across the population (36,630 subjects)**. The regions are organized to show region pairs within the same network together.

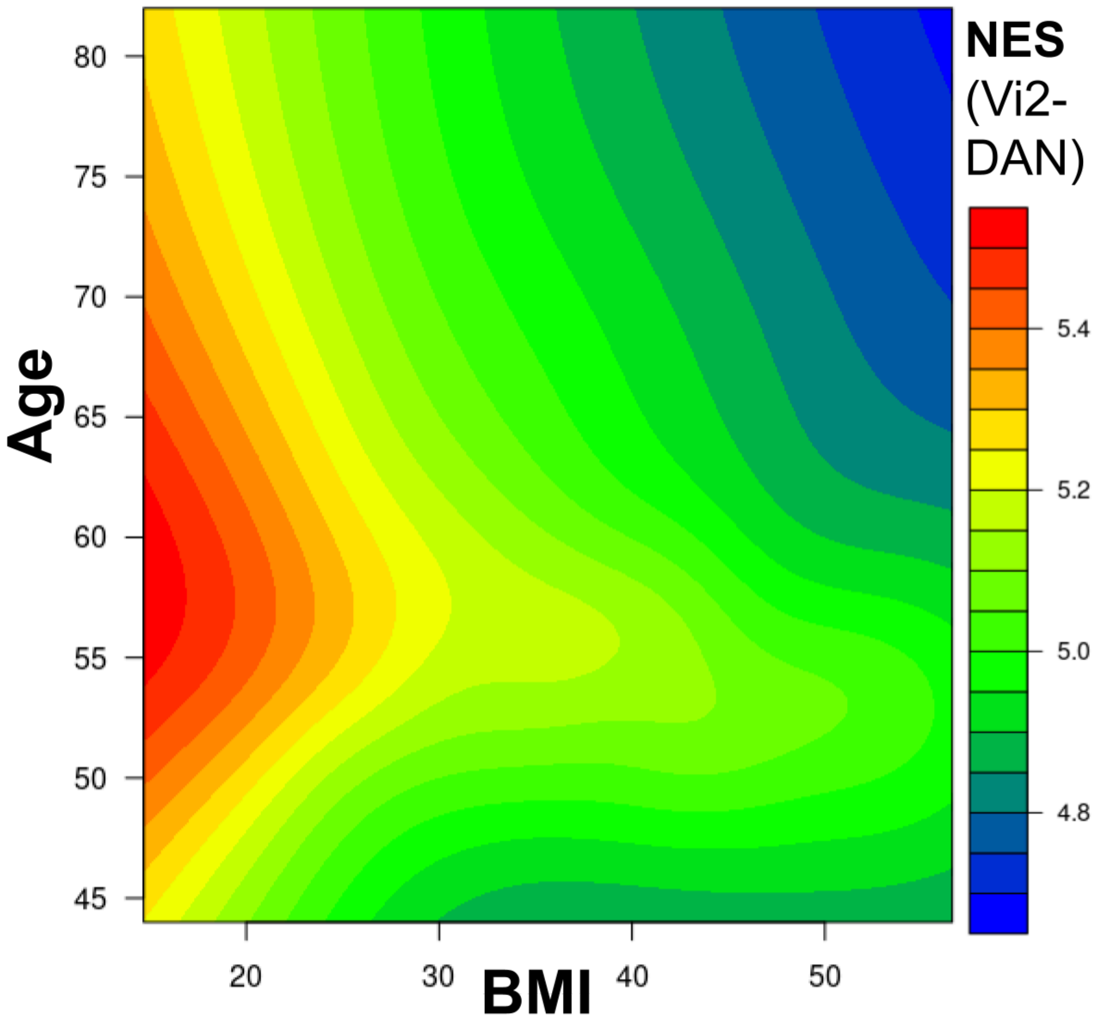


**eFigure 49. The contour plot of the network-level edge strength (NES) measures between visual2 and dorsal attention networks versus Age and BMI across the white British population (n = 33,824).** This may indicate a negative age-BMI interaction effect. The estimated bivariate function of age and BMI is fitted through cubic splines.

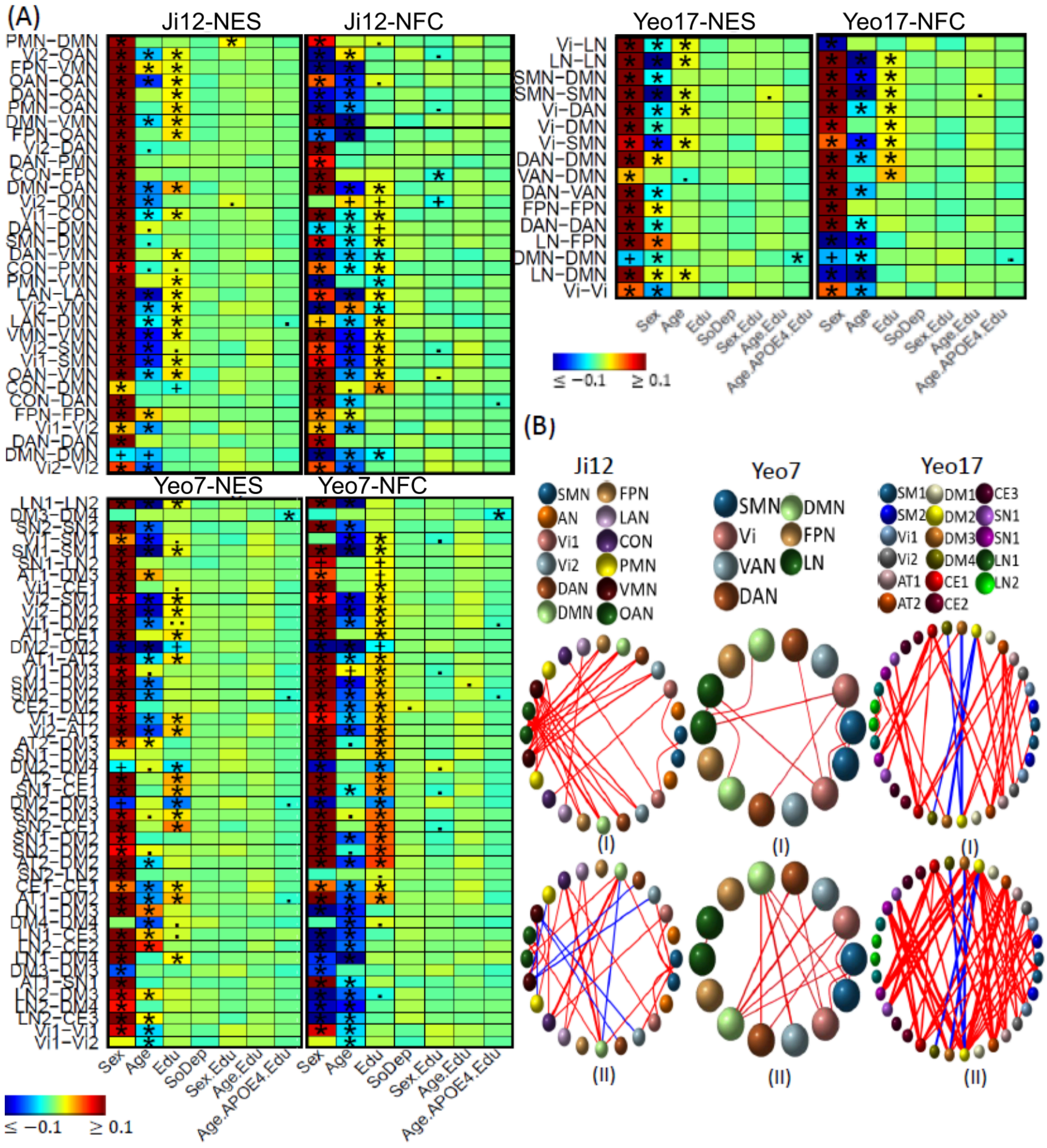

**eFigure 50. Selected association results of education with brain functional network connectivity measures.** (A) Heatmaps showing association results from the white British population using Ji-12, Yeo-7, and Yeo-17, for network edge strength (NES; I) and network functional connectivity (NFC; II). Significant results validated in the non-British population are marked with (*); significant but unvalidated results are indicated by (+). Non-significant results but with p-values <1e-4 and <1e-3 are denoted by (.) and (..), respectively. (B) Circular plots showing NES (I) and NFC (II) associations from the three atlases. Colored spheres represent different networks with positive (red) and negative (blue) connections.
